# Genome-wide association study of adolescent-onset depression

**DOI:** 10.1101/2025.09.26.25335972

**Authors:** Poppy Z Grimes, Brittany L Mitchell, Katherine N Thompson, Qingkun Deng, Xueyi Shen, Jareth C Wolfe, Jodi T Thomas, Robyn E Wootton, Daniel E Adkins, Saranya Arirangan, Elham Assary, Chris Chatzinakos, Charlotte A Dennison, Swathi Hassan Gangaraju, Andreas Jangmo, Yeongmi Jeong, Siim Kurvits, Qingqin S Li, Ehsan Motazedi, Joonas Naamanka, Thuy-Dung Nguyen, Ilja M Nolte, Vanessa K Ota, Joëlle A Pasman, Mina Shahisavandi, Amy Shakeshaft, John R Shorter, Chloe Slaney, Martin Tesli, Carol A Wang, Uxue Zubizarreta-Arruti, PGC MDD Working Group, GLAD+, NIHR BioResource, Silvia Alemany, Ole A Andreassen, Helga Ask, Sintia I Belangero, Rosa Bosch, Gerome Breen, Rodrigo A Bressan, Alfonso Buil, Enda M Byrne, Miquel Casas, William E Copeland, Thalia C Eley, Laurie J Hannigan, Catharina A Hartman, Alexandra Havdahl, Ian B Hickie, Golam M Khandaker, Kelli Lehto, Hermine Maes, Nicholas G Martin, Alexander Neumann, Albertine J Oldehinkel, Pedro M Pan, Hong Pan, Craig E Pennell, Roseann E Peterson, Alina Rodriguez, Giovanni A Salum, Tanja GM Vrijkotte, Robbee Wedow, Andrew JO Whitehouse, Anita Thapar, Henrik Larsson, Christel M Middeldorp, Andrew McIntosh, Mark J Adams, Yi Lu, Heather C Whalley, Alex SF Kwong

**Author notes:** **Corresponding authors**: Poppy Grimes and Alex Kwong. Joint last authors (Heather C Whalley, Alex SF Kwong).

## Abstract

Adolescent depression is a heritable psychiatric condition with rising global prevalence and severe long-term outcomes, yet its biological underpinnings remain poorly understood. We conducted the first genome-wide association study of adolescent-onset depression, comprising 102,428 cases (diagnosis or clinical symptom thresholds) and 286,911 controls, including diverse ancestries. Cross-ancestry meta-analysis identified 52 independent variants across 17 loci; European-only analysis found 61 variants at 29 loci, with a SNP-based heritability of 9.8%. Comparative analyses revealed two genes unique to adolescent-onset versus lifetime depression, enriched in neuronal subtypes, and two genes as potential drug repurposing targets. Polygenic scores were associated with adolescent-onset depression across ancestries, persistent depression trajectories, more severe outcomes, as well as reduced cortical volume, surface area and white matter integrity. Genetic correlation and Mendelian randomisation analyses support shared genetic liability and causal links with early puberty and modifiable health and behavioural risk factors. These findings uncover novel genetic loci and refine biological pathways underlying adolescent-onset depression, revealing age-specific mechanisms and early intervention opportunities.

## Introduction

Adolescence is a period marked by rapid biological and psychosocial changes that heighten vulnerability to mood disorders, particularly depression^1–3^. Adolescent depression is a leading cause of illness and disability in young people^4^, and nearly triples the risk of recurrence in adulthood^5^. Depression that emerges during this period is linked to long-term psychological, social, and functional impairments^6,7^, with more severe outcomes compared to later-onset depression^8^. Alarmingly, evidence suggests that prevalence is rising^9–11^, emphasising the urgent need to better understand adolescent depression – its developmental causes, underlying biology, and long-term consequences – to ultimately inform more effective intervention and prevention strategies.

Twin studies estimate that genetic factors explain up to 40% of the variance in depression risk, with stronger contributions at younger ages^12–14^, where genetic risk is compounded by greater exposure to social stressors^9^. Early-onset depression exhibits higher pedigree-based (55.1% vs. 43.85)^15^ and SNP-based (11.2-13.0% vs. 4.3-6.2%)^13,16^ heritability than later-onset forms, suggesting it may represent a biologically distinct subtype.

Further, given that adolescence is a sensitive period for both pubertal^17^ and neural development^18^, depression onset during this time likely involves unique pathophysiological pathways. These age-specific mechanisms may be obscured in genetic studies of lifetime depression due to confounding by illness chronicity or long-term consequences. Although the genetics of age-of-onset and early-onset depression have been studied^16,19,20^, no genome-wide association study (GWAS) has specifically examined adolescent-onset depression. Prior genetic research in young people has largely focused on depressive symptoms or internalising traits in European-only population samples, often with limited power and inconsistent phenotyping^21,22^, or on major depressive disorder onset in young adults^16^. These studies have not identified genome-wide significant loci for adolescent-onset depression, highlighting the need for age-specific genomic research to improve prediction and treatment development.

Here, we present the first GWAS meta-analysis of adolescent-onset depression, including ancestrally diverse populations which are typically underrepresented in genomic research (African, American-admixed, East Asian, European and South Asian)^23^. We leveraged data from prospective, retrospective, and clinical cohorts, defining cases based on one of the following criteria: (i) self-reported depressive symptoms meeting clinical thresholds between ages 10-19, (ii) self-reported diagnosis of major depressive disorder (MDD) between ages 10-19, or (iii) specialist-diagnosed MDD recorded in national health registers before age 25, used as a proxy for early-onset depression^16^.

To gain biological insight, we conducted gene-based association and mapping analyses to identify genes unique to adolescent-onset depression relative to lifetime major depression^24^, explored treatment targets via drug-gene interactions, and assessed human tissue- and neuronal cell-type-specific expression. To examine shared genetic aetiology and potential causal links, we evaluated genetic correlations and bidirectional Mendelian randomisation with related psychiatric, behavioural, and physical health traits. We tested the trait-association of polygenic scores across multiple ancestries and longitudinal adolescent psychiatric phenotypes. Lastly, we examined how polygenic scores relate to structural brain features, clinical outcomes, and pubertal timing in prospective longitudinal cohorts.

### Cross-ancestry and European GWAS of adolescent-onset depression

Our cross-ancestry analysis of adolescent-onset depression included 102,428 cases and 286,911 controls, drawn from 25 cohorts across 12 countries (**Supplementary Tables 1-2**); 3.8% of individuals were from non-European ancestries. Genome-wide association estimates of autosomes were meta-analysed, identifying 52 independent genome-wide significant SNPs located at 17 genomic risk loci (linkage disequilibrium [LD] r^2^ < 0.1, genomic inflation factor [λ_GC_] = 1.105) (**Figure 1**, **Supplementary Table 3**).

**Figure 1.**
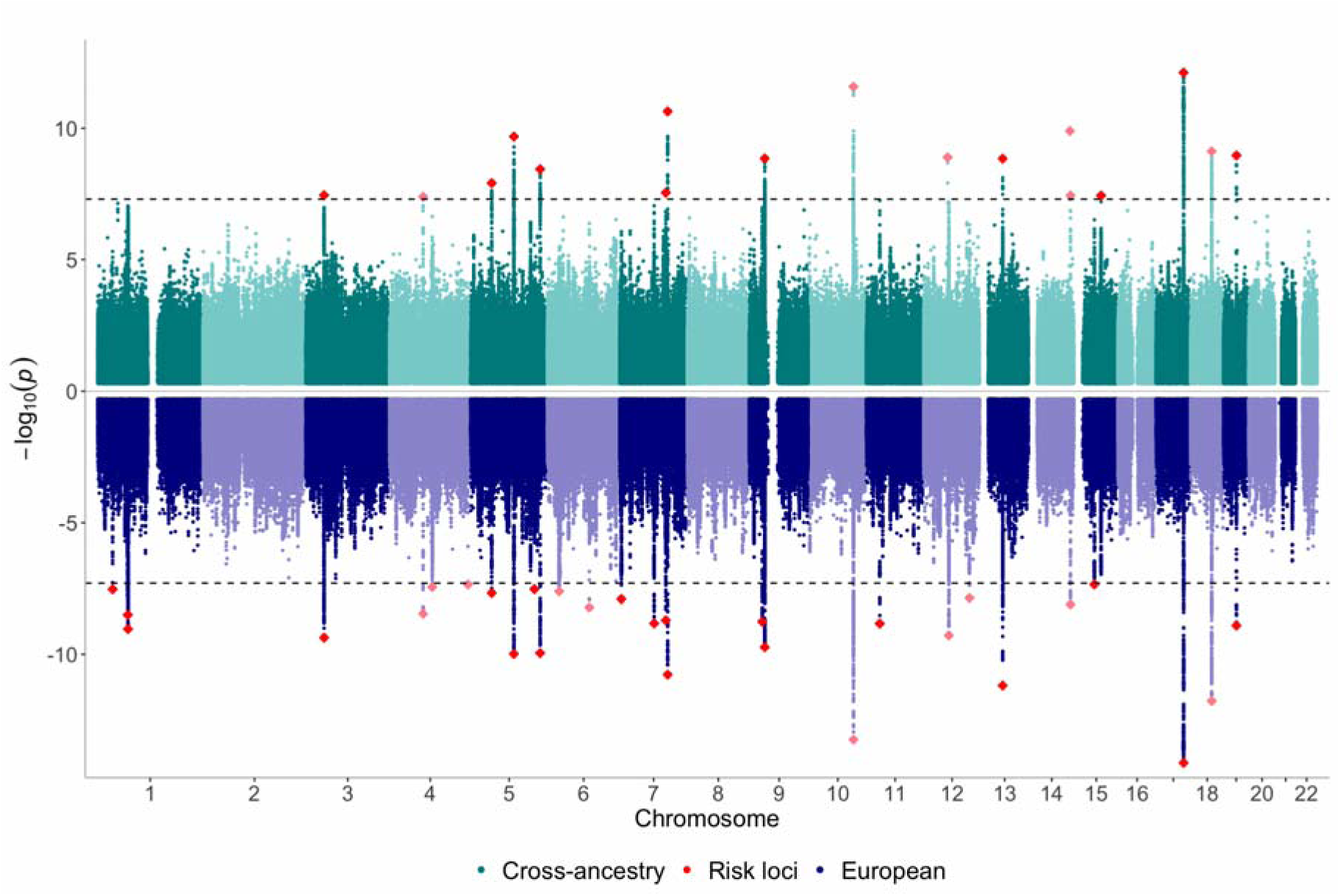
Miami plot of the cross-ancestry (top panel) and European (bottom panel) GWAS meta-analysis of adolescent-onset depression. The autosomal SNP association strength in the meta-analyses is indicated by -log_10_(p) value on the y-axis. Genome-wide significant loci are depicted as red diamonds.

As the largest single ancestry group, our European-only sample included 98,160 cases and 276,550 controls. Genome-wide association identified 61 independent lead variants at 29 genomic risk loci (**Figure 1**, **Supplementary Table 4**). In European ancestry, liability-scaled SNP heritability (SNP-*h^2^_liability_*) estimated with SBayesS was 9.8% (SE=0.3%) assuming a population prevalence of 15% and ranged from 7.0% to 10.7% when assuming prevalences between 5–20%. λ_GC_ as estimated with LD score regression (LDSC) was 1.26 with 81% of inflation likely due to true polygenicity (**Supplementary Table 5**). X chromosome analysis of European ancestry cohorts did not reveal any genome-wide significant loci (**Supplementary Figure 1)**.

Next, we stratified the European ancestry genome-wide association analyses by phenotype ascertainment type: (i) prospective self-reported symptoms meeting clinical thresholds between ages 10-19, (ii) retrospective self-reported clinical symptom threshold or diagnosis between ages 10-19, and (iii) specialist-diagnosed treatment before age 25. The stratified GWASs identified one genome-wide significant locus in the retrospective sample and one in the specialist diagnosis sample (**Supplementary Figures 2-4**).

We also ran meta-analyses within each non-European ancestry (African, American-admixed, East Asian, South Asian). We identified a single genome-wide significant locus in African ancestry individuals, but none in the American-admixed, East Asian or South Asian samples, though some suggestive signals (**Supplementary Figures 5-8**). For downstream analyses, we used the European ancestry only meta-analysis as the largest single ancestry sample, ensuring compatibility with LD reference panels.

### Correlations and factor model of ascertainment types in adolescent-onset depression

All ascertainment types showed moderate to strong genetic correlations: prospective with register-based (*r_g_*=0.89 [0.10], P*_rg=0_*=5.00×10^−19^, P*_rg=1_*=2.48×10^−01^), prospective with retrospective (*r_g_*=0.58 [0.09], P*_rg=0_*=8.03×10^−11^, P*_rg=1_*=3.72×10^−06^), and retrospective with register-based (*r_g_*=0.77 [0.04], P*_rg=0_*=1.57×10^−92,^ P*_rg=1_*=4.91×10^−10^). We used genomic structural equation modelling^25^ to assess the relative contributions of the different ascertainment types to the genetic liability underlying adolescent-onset depression by fitting a one-factor model (**Figure 2**). We fixed the clinical diagnosis phenotype for register-based early-onset depression as the primary indicator (factor loading = 1, residual variance = 0.01). All ascertainment types showed strong positive loadings on the common factor (clinical diagnosis = 1.0 [reference], prospective = 0.83 ± 0.06, retrospective = 0.75 ± 0.04). Model fit showed that the model captured the data structure well ( ^2^=2.53, *p*=0.111, CFI=0.997, SRMR=0.038), indicating that a single factor reflecting the variance in the clinical diagnosis phenotype adequately explained the shared variance across all phenotypes.

**Figure 2.**
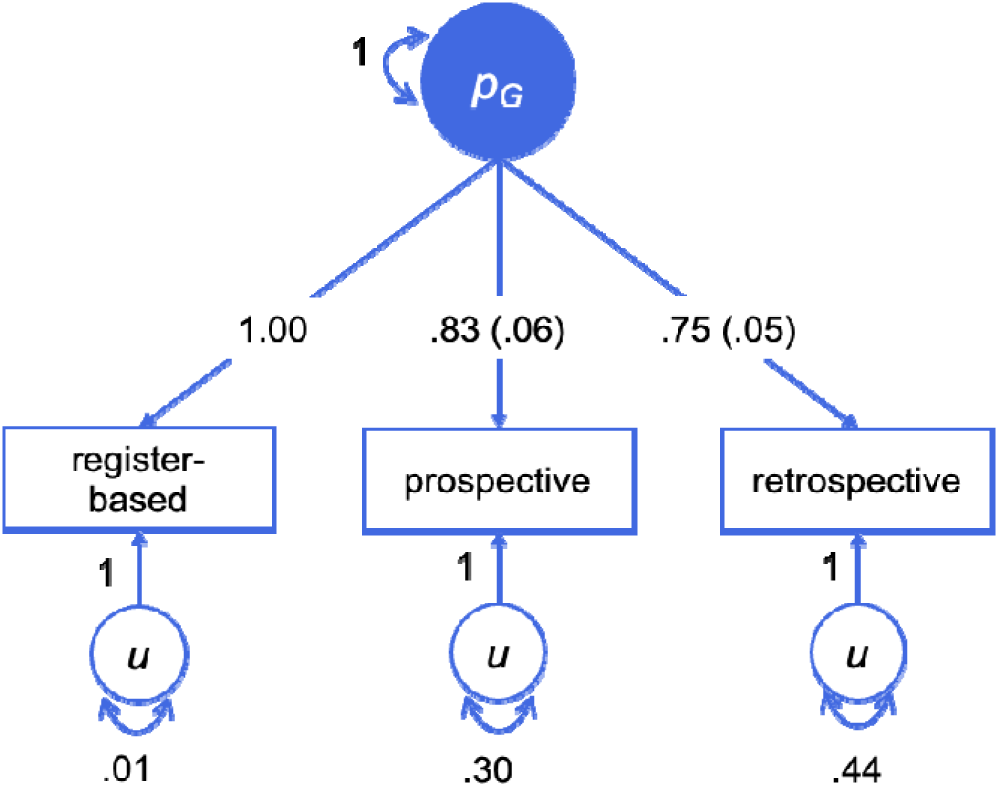
GenomicSEM path diagram showing genetic loadings of adolescent-onset depression phenotype ascertainment on a latent factor. p_G_ = genetic common factor, register-based = register-based early-onset diagnosis, prospective = prospectively self-reported, retrospective = retrospectively self-reported. Numbers represent standardised loadings with standard errors in brackets. The clinical phenotype loading was fixed to 1 as the reference. Self-directed arrows indicate variance of p_G_ or the residual variances of each phenotype (u).

### Gene mapping and gene-based association

To prioritise potentially causal genes, we mapped genome-wide significant SNPs to genes using both positional mapping and expression quantitative trait loci (eQTL) information (P*_FDR_*<0.05) from human brain tissue, via the SNP2GENE function in FUMA v1.6.6^26^. SNPs located within the 29 genome-wide significant loci were positionally mapped to 84 protein-coding genes (**Supplementary Table 6**). eQTL mapping further implicated 19 unique genes, of which 15 overlapped with the positionally mapped genes (**Supplementary Table 7**).

We next conducted MAGMA gene-based association testing, aggregating SNP-level association across each protein-coding gene to identify broader gene-level signals. This analysis identified 23 genes that were significantly associated with adolescent-onset depression after Bonferroni correction (P_bon_<2.547×10^−6^; 0.05/19,631 protein coding genes) (**Table 1, Supplementary Figure 9, Supplementary Table 8**). Overlap between genes identified with the three methods is in **Supplementary Table 9**.

**Table 1.** Genes significantly associated with adolescent-onset depression based on MAGMA gene-based analysis. Genomic coordinates refer to hg19. Z-scores and p-values reflect the strength of association across all SNPs assigned to each gene. Shaded rows highlight genes that were not identified with positional mapping.

| Gene | Chromosome | Start | Stop | N<br>SNPs | Z | P |
| --- | --- | --- | --- | --- | --- | --- |
| SORCS3 | 10 | 106400859 | 107024993 | 1820 | 7.5708 | 1.86E-14 |
| BPTF | 17 | 65821640 | 65980494 | 280 | 6.8334 | 4.15E-12 |
| DCC | 18 | 49866542 | 51057784 | 4590 | 6.4764 | 4.70E-11 |
| KPNA2 | 17 | 66031635 | 66042958 | 36 | 6.1085 | 5.03E-10 |
| ZCCHC7 | 9 | 37120536 | 37358146 | 302 | 5.8924 | 1.90E-09 |
| OLFM4 | 13 | 53602894 | 53626192 | 80 | 5.8374 | 2.65E-09 |
| C17orf58 | 17 | 65987217 | 65989765 | 2 | 5.7893 | 3.53E-09 |
| PAX6 | 11 | 31806340 | 31839509 | 44 | 5.5679 | 1.29E-08 |
| LSAMP | 3 | 115521235 | 117716095 | 6578 | 5.4539 | 2.46E-08 |
| BCL11B | 14 | 99635624 | 99737861 | 279 | 5.4367 | 2.71E-08 |
| SDK1 | 7 | 3341080 | 4308632 | 3806 | 5.324 | 5.08E-08 |
| TENM2 | 5 | 166711804 | 167691162 | 2335 | 5.1744 | 1.14E-07 |
| PCLO | 7 | 82383329 | 82792246 | 1197 | 5.1642 | 1.21E-07 |
| MMAB | 12 | 109991542 | 110011679 | 92 | 4.9096 | 4.56E-07 |
| BANK1 | 4 | 102332443 | 102995969 | 1893 | 4.8697 | 5.59E-07 |
| SEMA6D | 15 | 47476298 | 48066420 | 1770 | 4.8279 | 6.90E-07 |
| OR2B2 | 6 | 27878963 | 27880174 | 4 | 4.8145 | 7.38E-07 |
| MAP7D1 | 1 | 36621180 | 36646450 | 29 | 4.7681 | 9.30E-07 |
| GABBR1 | 6 | 29523406 | 29601753 | 284 | 4.6403 | 1.74E-06 |
| ZSCAN12 | 6 | 28346732 | 28367511 | 69 | 4.6396 | 1.75E-06 |
| FBXO10 | 9 | 37510889 | 37588871 | 225 | 4.6112 | 2.00E-06 |
| PDE4B | 1 | 66258197 | 66840259 | 1448 | 4.5989 | 2.12E-06 |
| RP11-613M10.8 | 9 | 37512544 | 37592466 | 234 | 4.5965 | 2.15E-06 |

To identify shared biological pathways, we conducted gene set enrichment analysis of 17,023 pathways (curated gene sets: 6,494, GO terms: 10,529) from Molecular Signatures Database MsigDB v2023.1Hs in MAGMA. No gene-sets met the P_bon_<0.05 significance threshold, however, the strongest signal was for the GOBP_FOREBRAIN_NEURON_FATE_COMMITMENT gene set, which relates to the developmental commitment of cells to become neurons in the forebrain. The top ten gene sets are reported in **Supplementary Table 10**.

### Treatment targets

Two of the MAGMA-identified genes were linked to six targetable biological entities according to the DrugBank 6.0 database^27^. These biological entities were linked to 11 medicines, and five of them were under the ‘Nervous system’ ATC classification, where medications for psychiatric conditions are categorized. Specifically, *GABBR1* is targeted by medicines for psychiatric conditions, such as clozapine (for schizophrenia) and progabide (for depression and anxiety). *PDE4B* is linked with five targetable proteins targeted by eight medicines, some of which are used for cardiovascular conditions (e.g., amrinone). A list of linked medicines can be found in **Supplementary Table 11**.

### Tissue-specific and cell-type specific expression

Using MAGMA gene-property analysis, we regressed gene-level association statistics on expression profiles across 30 general tissue types (GTEx v8) and identified genes significantly expressed in brain and pituitary gland tissue that were associated with adolescent-onset depression (at P_bon_<1.67×10^−3^) (**Supplementary Figure 11, Supplementary Table 12**). Across 53 specific human tissue types (GTEx v8), genes were significantly expressed in the cerebellar hemisphere, cerebellum, frontal cortex and cortex (P_bon_<9.43×10^−4^) (**Supplementary Figure 11, Supplementary Table 13**).

Two genes, *BCL11B* and *MAP7D1*, identified through gene-based testing, did not overlap with any high-confidence genes reported for lifetime MD and were not in LD with those loci^24^. To further investigate their biological relevance to adolescent-onset depression, we examined their expression in broad cell types, and major cell clusters as identified by^28^ in adolescent brain tissue (**Figure 3, Supplementary Tables 14-15**).

**Figure 3.**
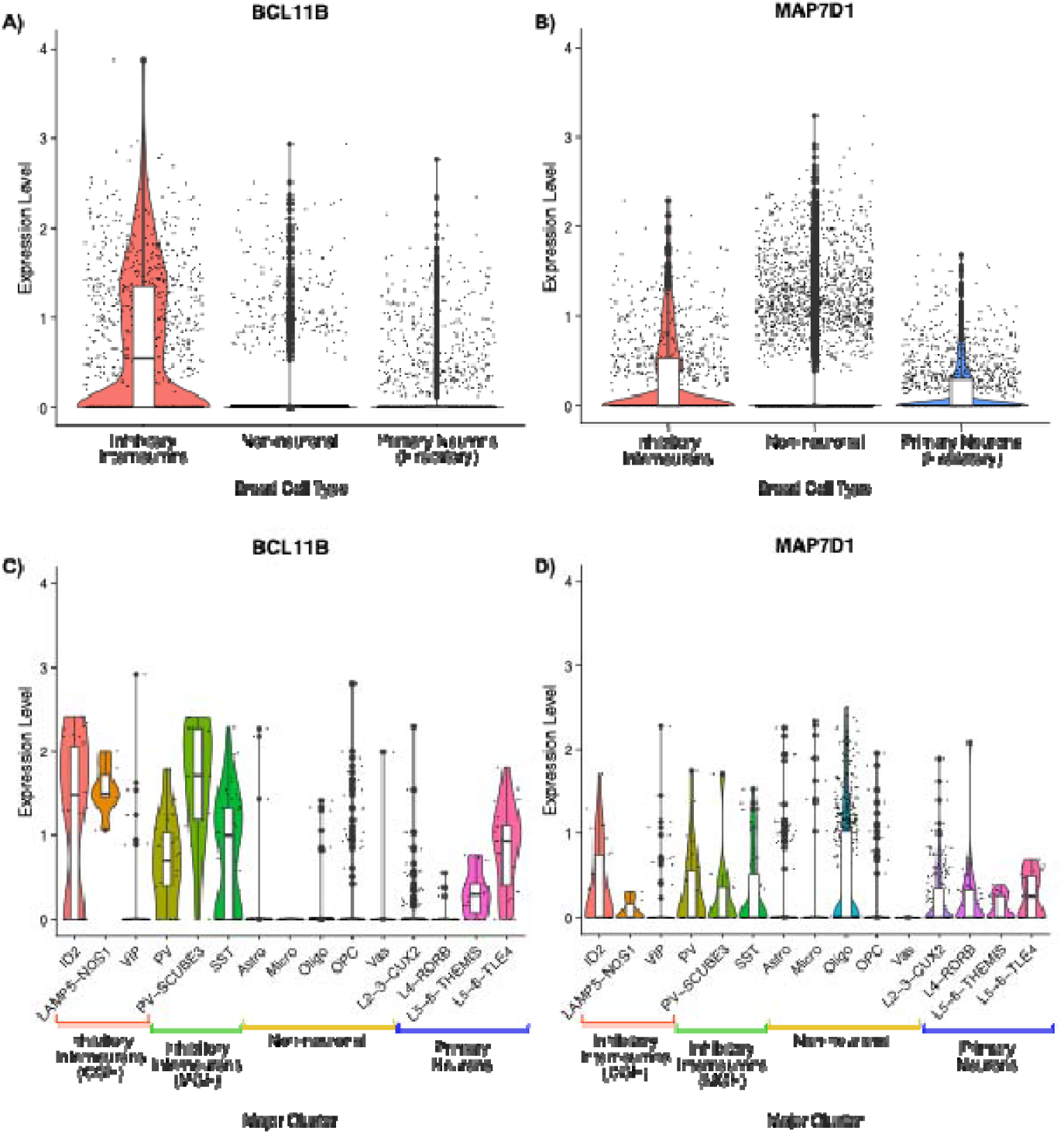
Expression patterns of BCL11B and MAP7D1 across neuronal subtypes in adolescent brain tissue. Violin plots show the distribution of expression levels for BCL11B (**a**) and MAP7D1 (**b**) across three broad cell classes: inhibitory neurons (red), non-neuronal cells (grey), and excitatory neurons (blue). Panels (**c**) and (**d**) depict expression of BCL11B and MAP7D1, respectively, across 15 specific brain cell subtypes (major clusters). The y-axis represents the log normalised expression level for each cell.

*BCL11B* showed enriched expression in inhibitory interneurons (INs) in adolescents, specifically in *ID2* and *LAMP5-NOS1* subtypes from the caudal ganglionic eminence, and in parvalbumin-expressing (*PV*), PV-*SCUBE3* and *SST* INs from the medial ganglionic eminence. *PV-SCUBE3* INs showed significantly higher expression of *BCL11B* than *PV* or *SST* INs (P=7.49×10^−5^ & 2.93×10^−6^ respectively). *BCL11B* was also expressed in deep-layer excitatory projections neurons (PNs) (L5/6-*THEMIS* and L5/6-*TLE4*), consistent with its role in neuronal development^29,30^.

*MAP7D1* was broadly expressed in INs and excitatory neurons, with significantly higher expression in INs than in PNs (P=2.96×10^−6^) or non-neuronal cells (P<2.2×10^−16^). *MAP7D1* was also enriched in oligodendrocytes (Oligo) compared to astrocytes (Astro), microglia (Micro), oligodendrocyte precursor cells (OPC) and vasculature associated cells (Vas) (P<2.2×10^−16^ for Astro, Micro, Oligo and OPC; and P=1.15×10^−13^ for Vas), though no major cell-subtype-specific differences were observed.

### Relationships with other traits

We investigated genetic correlations (*r_g_*) between our GWAS of adolescent-onset depression and a range of psychiatric and non-psychiatric health and behavioural traits using bivariate LD-score regression^31^ (**Figure 4 and Supplementary Tables 16-17)**. Strong positive correlations (0.6

**Figure 4.**
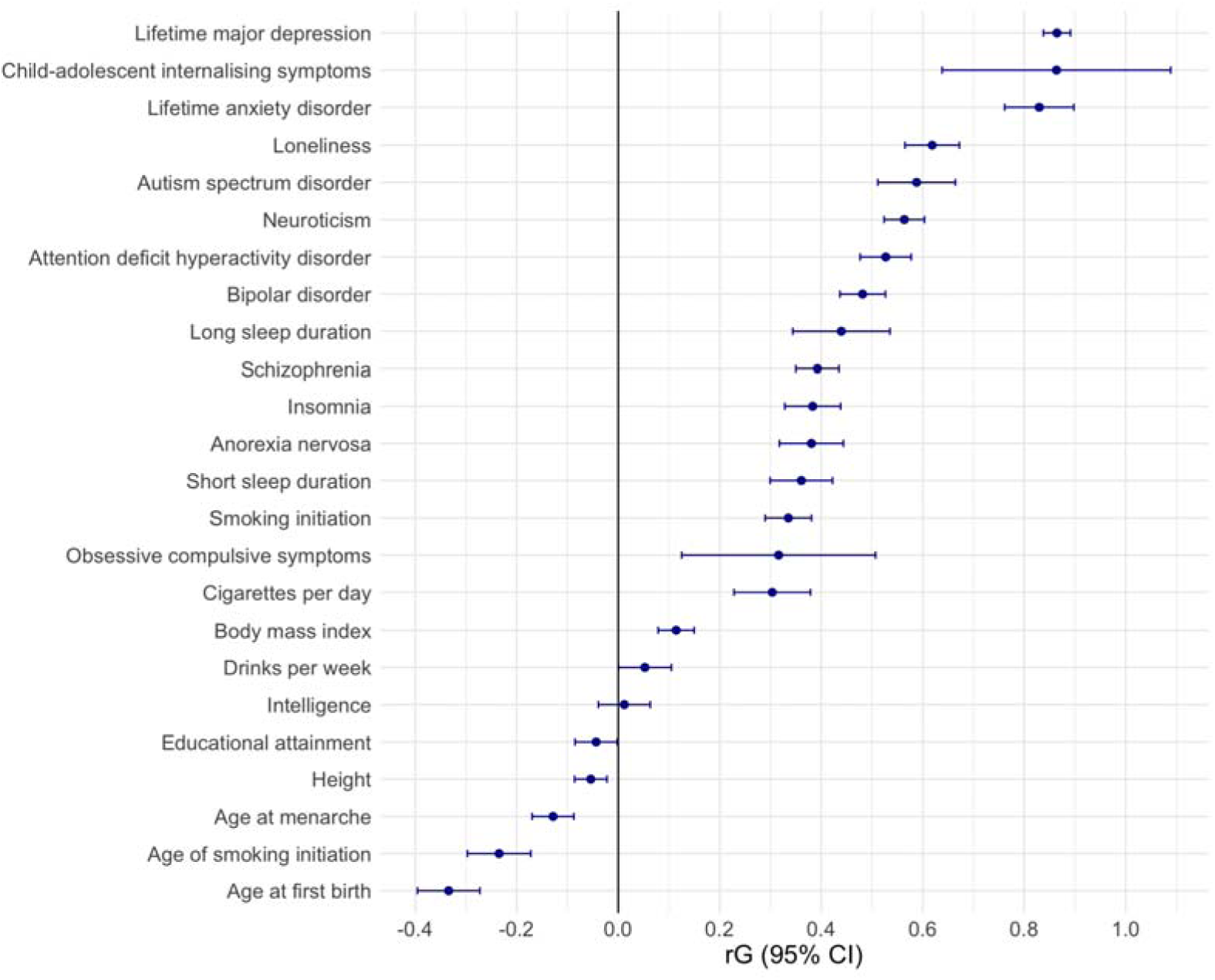
Genetic correlation results for the main meta-analysis with a range of psychiatric and non-psychiatric traits. Point estimates represent the genetic correlation value and error bars show 95% confidence intervals. All genetic correlations were significant after accounting for FDR, except the genetic correlation for intelligence.

<*r_g_*>1.0) were observed between adolescent-onset depression and multiple psychiatric traits including lifetime major depression and anxiety. Moderate positive correlations (0.4

<*r_g_*>0.59) were observed with autism, neuroticism, ADHD, and bipolar disorder, loneliness and long sleep duration.

We also tested genetic correlations between each trait and phenotype ascertainment types (**Supplementary Table 16, Supplementary Figure 12)**, and with lifetime MD^24^ for comparison with adolescent-onset depression (**Supplementary Table 16, Supplementary Figure 13)**.

We performed two-sample Mendelian randomization (MR) to test for potential bidirectional causal relationships between hypothesis-based traits and adolescent-onset depression (**Figure 5**). Analyses used the *TwoSampleMR* R package (v0.6.17)^32^, with inverse-variance-weighted (IVW) regression as the primary method and sensitivity analyses including the weighted median (WMedian), weighted mode (WMode), and MR Egger intercept test to assess horizontal pleiotropy. Associations were considered putatively causal if the IVW estimate survived FDR correction, the WMedian and WMode estimates were directionally consistent with the IVW estimate, and the MR Egger intercept was nonsignificant. To minimise bias from sample overlap, MR analyses were restricted to UK Biobank (UKB) traits, excluding UKB participants from the adolescent-onset depression GWAS.

**Figure 5.**
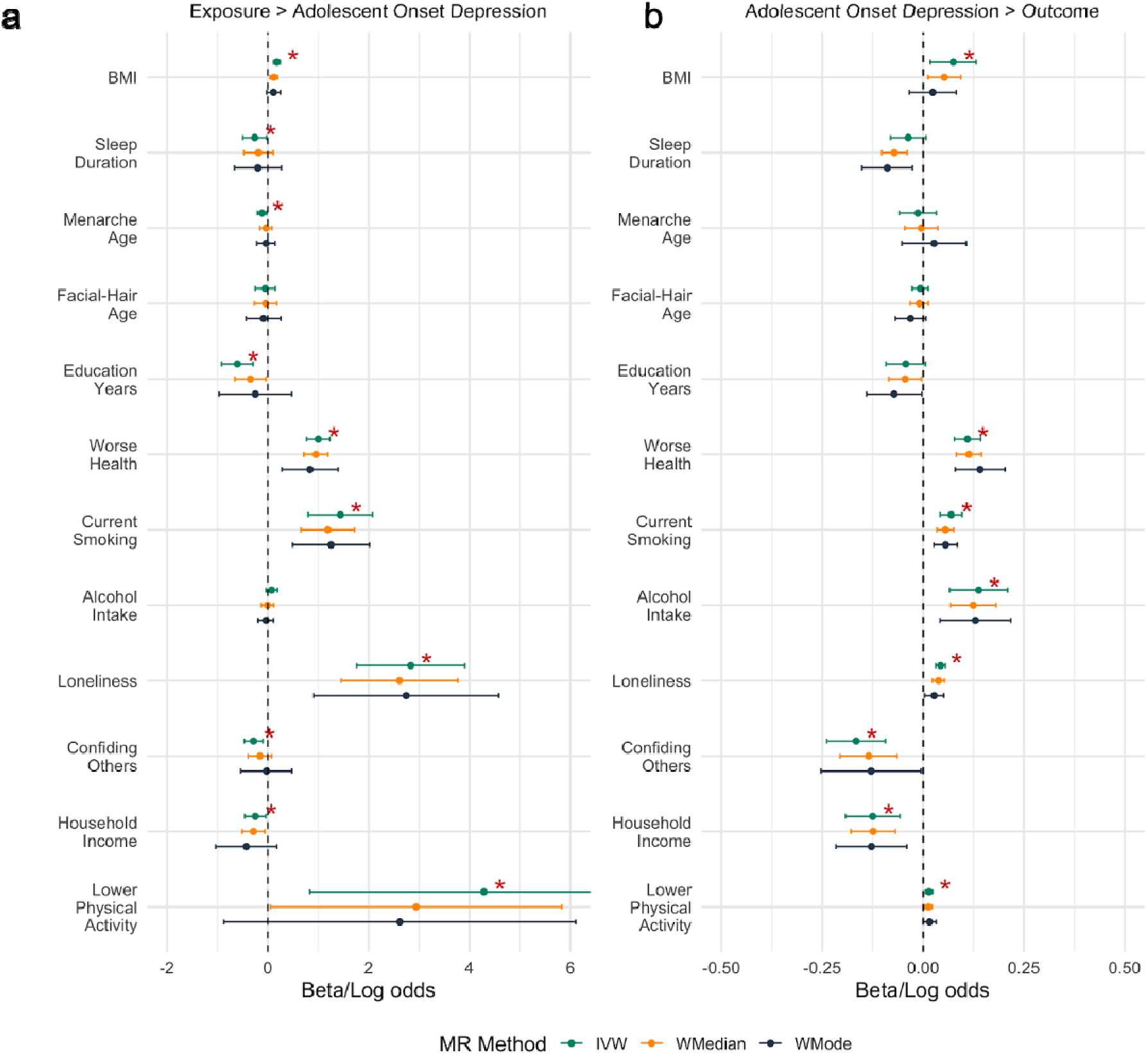
Bi-directional Mendelian randomization (MR) between traits and adolescent-onset depression on the beta/log odds scale. **a,** MR estimates for the effect of traits on adolescent-onset depression. For ‘No Physical Activity’, the upper 95% CI for the IVW extends to 7.742. **b,** MR estimates for the effect of adolescent-onset depression on traits. * indicates that the IVW method was significant at the FDR level, and the effect size was in the same direction for both the Weighted Median and the Weighted Mode methods. IVW: inverse-variance weighted method. WMedian: weighted median method. WMode: weighed mode method. MR Egger intercepts are given in Supplementary Table 18.

Our MR analyses provided putative evidence for causal effects of higher BMI, shorter sleep duration, earlier age of menarche, fewer years of education, poorer self-reported health, current smoking, and greater loneliness on adolescent-onset depression, with loneliness showing the strongest effect (beta_IVW_=2.835, [1.763, 3.909], P*_FDR_*=1.06×10^−06^). Additional putatively causal effects were observed for less confiding in others, lower household income, and lower physical activity, although these traits showed some evidence of horizontal pleiotropy (P≥0.054). (**Supplementary Table 18**).

In the other direction, we observed putative evidence for a causal effect of adolescent-onset depression on higher BMI, poorer self-reported health, current smoking, higher alcohol intake frequency, greater loneliness, lower confiding in others and lower household income, with confiding in others (beta_IVW_=-0.166, [-0.238, -0.093], P*_FDR_*=2.50×10^−05^) and higher alcohol intake frequency (beta_IVW_=0.138, [0.066, 0.210], P*_FDR_*=3.98×10^−04^) showing the strongest effect sizes. There was putative evidence for a causal effect of adolescent-onset depression on lower physical activity, however this also showed some evidence of horizontal pleiotropy (P=0.084). (**Supplementary Table 18**).

### Polygenic score prediction across psychiatric, developmental and brain phenotypes

#### Cross-ancestry out-of-sample PGS prediction

We used SBayesRC^33^ to generate polygenic scores (PGSs) for adolescent-onset depression based on leave-one-out versions of both the cross-ancestry and European-only meta-analyses, applying a fixed effects IVW standard error model. We chose the fixed effects model over the MR-MEGA cross-ancestry meta-analysis because it provides SNP effect sizes and standard errors, required for PGS prediction, although it is less powerful for cross-ancestry discovery^34^.

We tested PGS association with case-control status in 2,071 African, 2,523 American-admixed and 5,950 European ancestry individuals from the Adolescent Brain and Cognitive Development (ABCD-US) study, and 1,971 African, 986 American-admixed, and 5,727 European individuals from the National Longitudinal Study of Adolescent to Adult Health (Add Health). Fixed-effects meta-analysis was applied across both cohorts within each ancestry group (**Figure 6a, Supplementary Table 19**).

**Figure 6.**
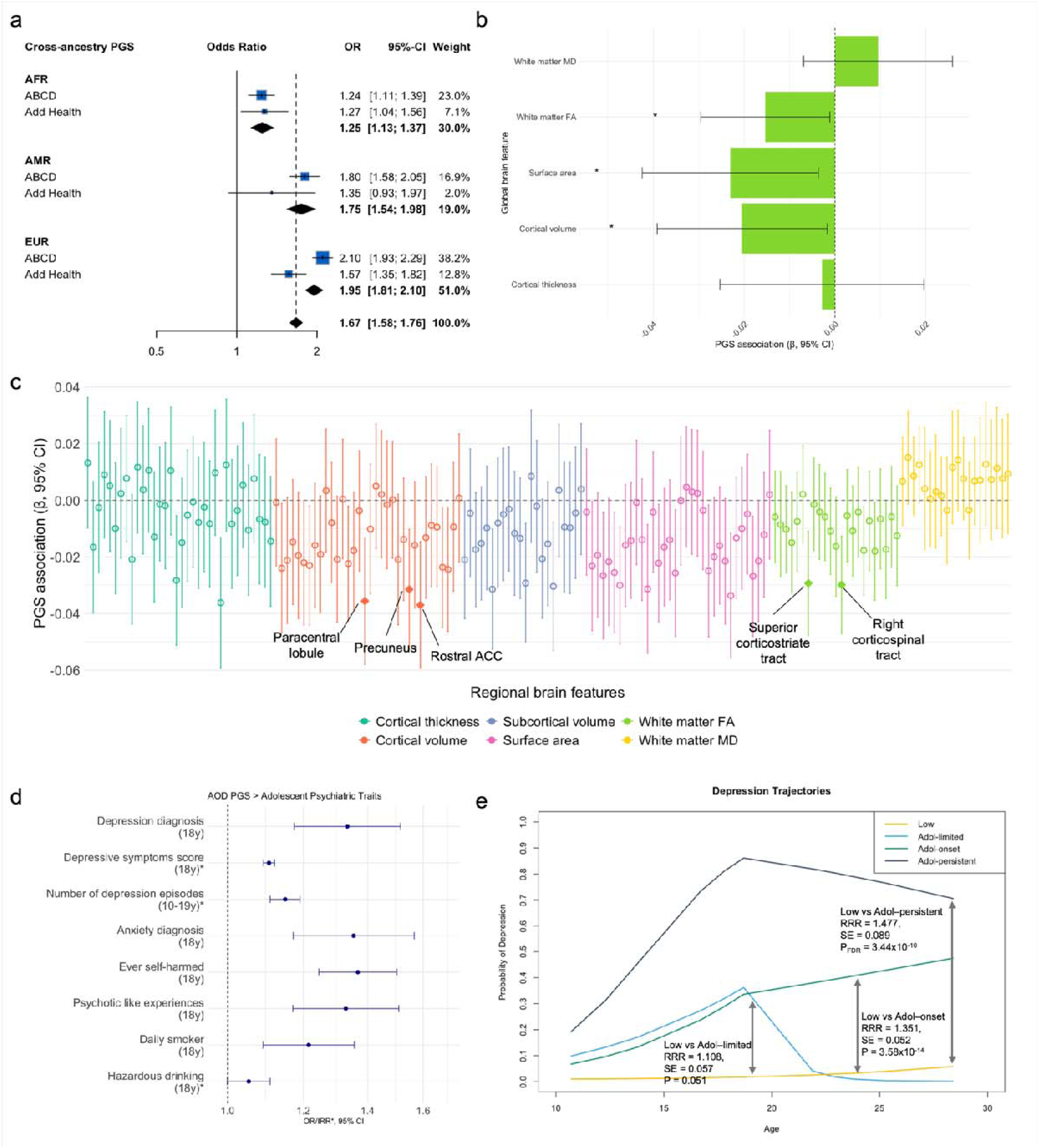
Polygenic score prediction across psychiatric, development and structural brain phenotypes. **a,** Meta-analysed effect (odds ratio) of the cross-ancestry PGS on adolescent-onset depression case-status in the ABCD-US and Add Health cohorts for African, American-admixed, and European ancestries (95% confidence interval). **b,** Cross-ancestry PGS association with structural brain features in the ABCD-US study. Bars plotted on the y-axis represent the standardised regression coefficients with 95% confidence interval error bars. **c**, Cross-ancestry PGS association with regional brain features in the ABCD-US study. The y-axis represents the within-modality FDR corrected (q < 0.05) standardised regression coefficients with 95% confidence intervals. Features are split by imaging modalities plotted along the x-axis. Solid dots represent features significantly associated with PGS after FDR correction. **d,** European PGS effects (odds ratio) on adolescent psychiatric traits in ALSPAC (95% confidence interval). **e,** PGS association with depression symptom trajectories in the ALSPAC cohort between the ages of 10 and 28 years (11 occasions). The reference category is the low trajectory for relative risk ratios (RRR).

Our PGSs derived from the cross-ancestry GWAS significantly predicted case-control status across all ancestry groups tested, though performed better in the European sample: European ancestry (OR=1.76 [95% CI: 1.57,1.97], P=1.05×10^−21^, ΔR²_liability_=0.8-1.9%), American-admixed ancestry (OR=1.64 [1.32, 2.04], P=9.54×10^−06^, ΔR²_liability_=0.4-1.1%), and African ancestry (OR=1.24 [1.06, 1.44], P=5.98×10^−03^, ΔR²_liability_=0.3%). We observed similar predictive performance using PGS derived from European ancestry only summary statistics (**Supplementary Table 19, Supplementary Figure 13**).

#### PGS association with structural brain phenotypes

To investigate how genetic risk for adolescent-onset depression relates to adolescent brain structure, we tested cross-ancestry PGS associations with 5 global and then 158 regional neuroimaging phenotypes in 7,490 participants aged 9-10 years from the ABCD-US study.

Globally, higher adolescent-onset depression PGS was associated with reduced whole brain cortical surface area (beta=-0.023 [95% CI: -0.04, -0.004], P=0.02), cortical volume (beta = - 0.020 [-0.04, -0.002], P=0.03), and global fractional anisotropy (FA) of white matter tracts (beta = -0.015 [-0.03, -0.001], P=0.03) (**Figure 6b**). Associations were marginally stronger with PGS derived from the European ancestry-only meta-analysis (**Supplementary Figure 14**).

In specific regions, higher adolescent-onset PGS was associated with reduced bilateral cortical volume in the paracentral lobule (beta=-0.04 [-0.06, -0.01], P_FDR_=0.03), precuneus (beta=-0.03 [-0.05, -0.01], P_FDR_=0.04), and rostral anterior cingulate cortex (ACC) (beta=- 0.04 [-0.06, -0.01], P_FDR_=0.03). It was associated with reduced FA in the bilateral superior corticostriate tract (beta=-0.03 [-0.05, -0.01], P*_FDR_* =0.02) and right hemisphere portion of the corticospinal tract (beta=-0.03 [-0.05, -0.01], P*_FDR_* =0.02) (**Figure 6c**). Four additional brain regions were significantly associated with the PGS based on the European ancestry-only meta-analysis: subcortical volume of the hippocampus and ventral diencephalon and reduced surface area of the fusiform gyrus and rostral ACC (**Supplementary Figure 15**). Associations between the cross-ancestry and European adolescent-onset PGSs with all 163 brain features are in **Supplementary Table 20**.

#### PGS association with adolescent psychiatric traits and puberty

We tested associations between the European ancestry-only adolescent-depression PGS with a range of prospective adolescent psychiatric phenotypes in the Avon Longitudinal Study of Parents and Children (ALSPAC) around age 18. Higher adolescent-onset depression PGS was associated with greater odds of a diagnosis of depression (OR=1.33 [95% CI 1.17, 1.51], P*_FDR_*=2.33×10^−05^, ΔR²_liability_=4.59%) and anxiety (OR=1.35 [1.17, 1.57], P*_FDR_*=7.68×10^−05^, ΔR²_liability_=6.26%), alongside more severe psychiatric phenotypes including ever having self-harmed before age 18 years (OR=1.37 [1.25, 1.50], P*_FDR_*=2.04×10^−10^, ΔR²_liability_=3.28%), psychotic like experiences (OR=1.33 [1.17, 1.51], P*_FDR_*=2.40×10^−05^, ΔR²_liability_=4.20%), and a greater number of depressive episodes between age 10 and 19 years (IRR=1.14 [1.11, 1.19], P*_FDR_*=1.61×10^−13^, ΔR^2^=0.47%) **(Figure 6d, Supplementary Table 21)**.

We modelled latent class trajectories of depressive symptoms between the ages of 10 and 28 years. Adolescent-onset depression PGS was associated with a greater likelihood of adolescent-onset (RRR=1.35 [1.25, 1.46], P*_FDR_*=3.58×10^−14^) and adolescent-persistent trajectories (RRR=1.48 [1.31, 1.66], P*_FDR_*=3.44×10^−10^) compared to the stable low trajectory. Higher PGS also distinguished between adolescent-limited and adolescent-onset trajectories (RRR=1.21 [1.09, 1.36], P*_FDR_*=5.60×10^−04^) (**Figure 6e, Supplementary Table 21**).

Finally, a higher PGS for adolescent-onset depression was associated with earlier puberty onset, as evidenced by an association with an earlier age of menarche in females (beta=-0.018 [-0.027, -0.008], P=.001, R^2^=0.5%) and an earlier age at peak height velocity in males (beta=-0.020 [-0.036, -0.004], P=.010, R^2^=0.6%).

## Discussion

We present the first large-scale GWAS of adolescent-onset depression, identifying 52 novel SNPs in our cross-ancestry analysis and 61 in our European ancestry-only analysis. Two genes (*BCL11B* and *MAP7D1*) identified in the European-only analysis appear specific to adolescent-onset, independent of lifetime major depression. Our results provide new and robust evidence for common variant contributions to adolescent-onset depression, addressing limitations of previous underpowered childhood-adolescent studies^21^. By combining multiple ascertainment methods and incorporating multiple ancestries, we increased discovery power and enabled downstream analyses and biological interpretability.

SNP-based heritability in the European ancestry group was 9.8%, higher than the 8.5% reported for lifetime depression^24^ supporting stronger genetic influences in adolescent-onset cases^13,35,36^. Cross-ancestry analysis revealed loci absent from European ancestry-only analyses, highlighting its value, though fewer loci overall were detected, likely due to heterogeneity and smaller sample sizes per ancestry. Importantly, <5% of participants were of non-European ancestry, emphasising the need for ongoing data collection efforts to expand data from across the ancestry spectrum.

A latent factor model showed that prospective, retrospective, and clinical diagnosis phenotypes all contributed to shared genetic liability for adolescent-onset depression, highlighting the value of integrating multiple ascertainment methods. Despite a strong latent factor loading (0.75), the retrospective phenotype correlated only moderately with the prospective phenotype (*r*_g_=0.58) and showed weaker, less consistent associations with external traits (**Supplementary Figure 12**). By contrast, prospective and clinical phenotypes displayed more robust genetic correlations with psychiatric and behavioural traits, particularly ADHD and loneliness, suggesting better capture of core neurodevelopmental and social impairments. Retrospective depression correlated more strongly with schizophrenia, bipolar disorder, and educational attainment, potentially reflecting recall bias, older reporting age, or inclusion of more severe or atypical cases.

Two genes – *BCL11B* and *MAP7D1* – emerged as unique to adolescent-onset depression, which could act on age-sensitive biological pathways such as cortical maturation and myelination. *BCL11B*, a transcription factor implicated in cortical development and immune regulation^37^, with links to neurodevelopmental conditions^30^, showed enriched expression in adolescent brain tissue across inhibitory interneurons and deep-layer excitatory neurons, consistent with its role in cortical circuit formation. *MAP7D1*, though less well-characterised, is involved in microtubule organisation during neuronal development^38,39^, and was expressed across inhibitory and excitatory neurons, with notable enrichment in oligodendrocytes – suggesting possible relevance to myelination and white matter maturation.

Among genes shared between adolescent-onset and lifetime major depression, *SORCS3* was the top hit. *SORCS3* encodes a hippocampal receptor regulating synaptic plasticity and AMPA receptor trafficking^40,41^, is involved in neurotrophin signalling, and has been implicated in multiple neuropsychiatric and neurodevelopmental conditions^24,42^, suggesting it may be an early marker of depression. *BPTF* also showed strong association, with roles in T-cell function^43^ and neurodevelopmental conditions^44^, and genetic overlap with COVID-19 severity and neuropsychiatric traits^45^. These associations with *BPTF* and *BCL11B* point to potential shared immune-neurodevelopmental pathways, consistent with evidence that childhood inflammation increases later depression risk^46,47^. We also identified two genes, *GABBR1* and *PDE4B*, which encode proteins targeted by existing compounds (e.g. clozapine for schizophrenia^48^ and theobromine in cocoa and tea linked to improved cognitive performance^49^), pointing to opportunities for treatment repurposing and broader relevance to neuropsychiatric pathways.

Our PGSs explained up to 2% of variance (ΔR²_liability_) in the European ancestry group – a substantial improvement over previous scores for adolescent internalising symptoms (0.03%)^21^ but lower than for lifetime MD (5.8%)^24^. While cross-ancestry prediction remains limited by allele frequency and LD differences^50^, we observed moderate transferability to American-admixed ancestry (up to ΔR²_liability_=1%) and African ancestry individuals (up to ΔR²_liability_=0.3%). Given low variance explained and unequal performance, clinical use remains premature and may risk exacerbating disparities^50,51^. Future work should prioritise using ancestry-matched GWASs for PGS training^52,53^ and integrate environmental risk factors as an important source of variance in depression risk^54,55^.

Genetic liability for adolescent-onset depression was associated with reduced cortical volume, surface area, and white matter integrity in 9–10-year-olds, suggesting early neurodevelopmental differences. Affected regions are relevant for emotion, cognition, self-referential processing, and motor function. These results support the hypothesis that genetic risk manifests in brain structure prior to symptom onset^56^ and extend previous neuroimaging studies^57,58^.

Our adolescent-onset depression PGS was associated with other psychiatric and behavioural traits in adolescence, including anxiety, self-harm, smoking, and psychotic-like experiences. Together, with genetic correlations and Mendelian randomisation methods, these findings suggest that shared genetic aetiology and potential causal pathways are likely to underpin the complex relationship between adolescent-onset depression and many related psychological, social and behavioural traits^9,59^. Importantly, our findings highlight opportunities for targeted, potentially modifiable interventions – such as improving sleep, education and social support at the population level, and reducing loneliness and increasing physical activity (the strongest MR signals) through educational settings. Targeting adolescent depression itself could also mitigate downstream outcomes, including poorer health, substance use, and poorer economic position, maximising both individual and societal benefit. Future work should extend these MR analyses to additional datasets and traits, such as neurodevelopmental conditions like ASD and ADHD, which share genetic overlap and may be affected by horizontal pleiotropy^60^. Findings should also be triangulated using complementary observational and other causal approaches.

Genetic risk is also more likely to underpin the course and persistence of depression in early development. Higher PGS was associated with more depressive episodes and greater likelihood of persistent depression trajectories, supporting the role of genetic risk in shaping depression severity and course across adolescence and young adulthood^35,60–62^. Our adolescent-onset PGS distinguished between adolescent-limited and adolescent-persistent symptom trajectories, suggesting higher genetic risk could differentiate between individuals likely to recover and those at risk of chronic depression post adolescence.

Genetic risk was also associated with earlier pubertal timing in both sexes, consistent with genetic correlations and MR evidence suggesting a causal effect of early puberty on adolescent-onset depression. We also identified gene expression in pituitary tissue, which secretes hormones regulating pubertal onset, and was absent in recent GWASs of lifetime MD^24,36^, suggesting adolescent-onset specific effects. These findings extend prior observational and causal work implicating puberty in adolescent depression onset^63,64^, providing the first evidence from adolescent-specific genetic markers that may influence neurodevelopment and hormonal change.

Together, our findings refine the genetic architecture of adolescent-onset depression, wherein genetic risk intersects with pubertal timing, psychosocial risk factors such as loneliness, brain maturation, and immune processes. By revealing partially distinct genetic components from lifetime major depression and integrating analyses from genes to brain to developmental biomarkers, our study advances understanding of adolescent depression’s biological basis. With rates of adolescent and young adult mental health in decline at present, these findings offer important insights to help identify target points for early treatment and preventative intervention.

## Online Methods

### GWAS meta-analysis overview

Our study aimed to identify genetic variants associated with adolescent-onset depression through a large-scale genome-wide association (GWAS) meta-analysis. The final cross-ancestry meta-analysis included data from 24 cohorts comprising 102,428 cases and 286,911 controls (**Supplementary Table 1**). Most individuals in this sample (96.2%) were of European ancestry. We performed two main meta-analyses: 1) cross-ancestry meta-analysis with all cohorts and 2) European ancestry only meta-analysis. In the European ancestry group, we took a stepwise approach based on combining phenotype ascertainment types. First, we meta-analysed prospective and retrospective cohorts independently, and then both samples together with a large clinical sample. We also performed ancestry-stratified meta-analyses for each non-European ancestry group. For all cohorts in the analyses, cohort descriptions, phenotype information, and ethics statements are in the **Supplementary Note.** Genotyping information is outlined below and in **Supplementary Table 2**.

### European ancestry samples and phenotype

#### Prospective adolescent sample

The prospectively ascertained data consisted of individuals from 19 samples from 15 cohorts (with some cohorts providing multiple samples from different batches or waves), many of which were recruited from the EAGLE consortium^65^. 20,432 prospective cases and 57,349 controls were assessed for depression onset between 10-19 years. Cases were individuals who met self-reported clinical symptom threshold cut-off for depression, and controls as those who did not. We chose self-reported symptoms rather than maternal-reported as this ascertainment type demonstrated higher heritability in a previous study^21^. Instruments included the Brief Problem Monitoring (BPM) scale^66^, the Centre for Epidemiological Studies Depression scale (CESD)^67^, the Child and Adolescent Psychiatric Assessment (CAPA)^68^, the Short Mood and Feelings Questionnaire (SMFQ)^69^, the Strength and Difficulties Questionnaire (SDQ)^70^, the Somatic and Psychological Health Report (SPHERE)^71^, and the Youth Self Report (YSR)^72^ (Further information in **Supplementary Table 1**).

#### Retrospective adult sample

The retrospective data consisted of 31,902 cases and 120,060 controls from 8 samples from 7 cohorts. Cases were individuals who retrospectively reported a depression episode between 10-19 years (inclusive), and controls were those who never reported an onset of depression in their lifetime. Retrospective instruments included the Composite International Diagnostic Interview (CIDI)^73^, the Mini International Neuropsychiatric Interview (MINI)^74^ and the Structured Clinical Interview for DSM Disorders (SCID)^75^ (Further information in **Supplementary Table 1**).

#### Early-onset clinical diagnosis sample

This sample, described in detail previously^16^, was from a recent GWAS meta-analysis of 46,708 cases and 106,824 controls. The study assigned cases and controls based on clinical diagnosis of early-onset depression who made contact for treatment up to age 25 years in a meta-analysis of cohorts from five Nordic countries: Denmark, Estonia, Norway, Sweden, and Finland. The early-onset age threshold of 25 was chosen based on prior meta-analytic findings indicating that the 25th percentile of age at onset (AAO) for depression falls around age 21, and on average, the onset predates first diagnosis by 5 years^2^. Therefore, individuals were classified as early-onset MDD cases if they had their first psychiatric treatment contact for MDD ≤ age 25 (approximate to AAO p25 at age 20-21)^16^. We used a leave-one-out sample, removing the Norwegian MoBa cohort to prevent sample overlap as it was already included as a prospective cohort, which resulted in 45,826 cases and 99,141 controls.

### Non-European ancestry samples and phenotype definition

Non-European genetic data were available for 4 samples of American-Admixed ancestry (2,136 cases and 3,701 controls), 4 samples of African ancestry (1,475 cases and 3,607 controls), 2 samples of East Asian ancestry (340 cases, 586 controls), 2 samples of South Asian ancestry (251 cases and 1481 controls) and 2 samples of admixed ancestry (66 cases and 986 controls) from a total of 8 cohorts (**Supplementary Table 1**). All cohorts had prospective reports of adolescent-onset depression, except for one which was retrospectively reported (UK Biobank). Instruments for depression case-control assignment included BPM, CAPA, CDI-2, CESD-5, CIDI, SDQ, SMFQ and YSR.

### Cohort-level GWAS – quality control, imputation and association analyses

A standard operating procedure was distributed to analysts for all participating cohorts advising the following quality control (QC) steps. Participants were excluded for low call rate, excess autosomal heterozygosity, duplicates, males with excessive X-chromosome homozygosity and chromosomal abnormalities. Related individuals were excluded unless the analysis was performed using software which can handle relatedness. Genotype QC was instructed to filter for minor allele frequency (MAF) < 0.01, minor allele count (MAC) < 20, a call rate of < 95%, Hardy-Weinberg Equilibrium (HWE) of p < 1e-06 and all other standard QC exclusions (e.g. evidence of poor clustering, strand ambiguous variants). Most cohorts followed the exact specifications with a handful performing different but nearing filtering thresholds. Full cohort-level quality control and imputation parameters are in **Supplementary Table 2**. Imputation panels varied across cohorts and included TOPMed, 1000 Genomes Phase III and the Haplotype Reference Consortium panels. Cohort-level GWAS were performed with REGENIE (15 samples), PLINK2 (8 samples) or SAIGE (2 samples) (**Supplementary Table 2**). Covariates included sex, year of birth, the first (4-10, depending on the cohort) principal components and any other relevant cohort-level covariates. The X chromosome was available in 10 cohorts and genotypes were coded 0 or 2 in males and 0, 1 and 2 in females.

### Single ancestry GWAS meta-analyses and identification of significant SNPs

All individual cohort summary statistics were aligned with the dbSNP reference panel (http://www.ncbi.nlm.nih.gov/SNP) to check that chromosome, base position and alleles match and were labelled consistently across cohorts. Variant chromosome, position and alleles were aligned with the Genome Reference Consortium Human Build 37 (GRCh37/hg19). Where necessary, liftover from other genome builds was performed, prioritising rsID matching and subsequently using CrossMap v0.6.1^76^ with NCBI dbSNP 155 patch 13.

We then used METAL^77^ software to perform a fixed effects inverse-variance weighted (IVW) standard error meta-analysis of the samples specifying the ‘SCHEME STDERR’ and ‘ANALYZE HETEROGENEITY’ commands.

For European ancestry, this was performed in two stages. First, we performed two independent meta-analyses of the prospective cohorts and retrospective cohorts. We then removed SNPs with MAF < 0.01 and SNPs that were present in < 50% of the contributing cohorts. In the second stage, we combined the prospective, retrospective and the register-based early-onset meta-analysed summary statistics into a single meta-analysis of all Europeans and removed SNPs with MAF < 0.005.

For non-European ancestry cohorts, we used the same model (fixed effects IVW standard error) in METAL to perform one meta-analysis within each of the following ancestries: African, American-admixed, East Asian, and South Asian. We then removed SNPs with MAF < 0.005.

For all sets of meta-analysed summary statistics, significant genomic risk loci and SNPs were identified with FUMA (https://fuma.ctglab.nl) using the conventional genome-wide significance threshold of P < 5×10^−8^. We used ancestry specific LD reference panels from 1000 Genomes provided in FUMA. Independent significant SNPs are defined as SNPs reaching the significance threshold and independent from each other (r^2^ = 0.6). This is equivalent to clumping GWAS tagged SNPs with the same P-value and r^2^. Independent significant SNPs were further clumped (r^2^ = 0.1) to define lead SNPs. Genomic risk loci – regions that represent all independent signals that are physically close – are identified by merging independent significant SNPs that are dependent on each other at r^2^ >= 0.1 and closer than 250kb distance with the top lead SNP representing each locus.

### Cross-ancestry GWAS meta-analyses

Though limited by the current availability of genetic data in non-European ancestries, we investigated whether association signals generalise and if we could capture variants beyond European ancestry. The cross-ancestry analysis was performed using the ancestry-aware MR-MEGA software^34^. MR-MEGA handles heterogeneity in allelic effects of differing genetic ancestries in an inverse-variance weighted random-effects meta-regression and has been used previously in one of the largest multi-ancestry studies of major depression in adults^78^. This software also allowed the inclusion of the 2 samples of admixed ancestry. We included the first 4 principal components for population stratification as standard practice with this software. Post-hoc QC involved the same filtering out of SNPs with MAF < 0.005. Significant SNPs and loci were identified with FUMA but using the 1000 Genomes ‘ALL’ reference panel.

### Estimation of liability scaled SNP-based heritabilities

In the European ancestry group, we used SBayesS (GCTB v2.5.2) to determine the SNP-based heritability (SNP-*h*^2^) of the summary statistics and converted results to the liability scale^79^ for a range of population prevalence estimates from 5-20%^9,80^. SBayesS uses shrunk sparse linkage disequilibrium (LD) matrices for >2.8 million UK Biobank variants. In non-European ancestry groups, we applied linkage disequilibrium score regression (LDSC)^31^, within each independent ancestry group. Ancestry-specific LD reference panels were used from the 1000 Genomes Project and heritability was converted to the liability scale. However, due to small sample sizes, non-European SNP-*h*^2^ were either negative or had large standard errors (crossing the null), limiting interpretability.

### Calculation of inflation due to polygenicity

Using LDSC, we further calculated the genomic inflation factor (λ_GC_) for each set of summary statistics and, if present, determined the proportion of inflation due to polygenicity vs. confounding. This was derived using the attenuation ratio: 1 – (LDSC intercept – 1 / λ_GC_ – 1) (**Supplementary Table 5**).

### GenomicSEM of phenotype ascertainment on a common factor

We used Genomic Structural Equation Modelling (GenomicSEM)^25^ to conduct multivariate modelling of the three ascertainment phenotypes (prospective, retrospective and clinical early-onset) in the European ancestry meta-analysis to see if they were reflective of a single underlying genetic factor. For each phenotype, we first munged summary statistics and then performed LDSC using the HapMap3 (Broad Institute) reference file for allele alignment. We then constructed a common factor model. In this model, a single overarching latent factor loads onto all three phenotypes and explains the shared covariance among them. We fixed the clinical early-onset phenotype to 1 as the reference and restricted its variance to 0.01 to prevent negative residual variances. From the model output we reported factor loadings, variances and model fit indices (chi-squared [ ^2^], *p*, comparative factor index [CFI], standardised root mean square residual [SRMR]).

### Functional annotation: genes, tissues and cells

To functionally interpret genome-wide significant loci, we performed gene-mapping, gene-based association testing and gene expression enrichment analyses using tools integrated in FUMA v1.6.6 for the European ancestry meta-analysis^26^. Loci were defined by independent lead SNPs and their corresponding LD blocks (r^2^ > 0.6, based on 1000 Genomes Phase 3 European reference panel). Functional mapping and annotation were conducted using the SNP2GENE pipeline in FUMA. We used two gene-mapping strategies. In positional mapping, SNPs were mapped to protein-coding genes if located within 10kb of the gene boundary (based on GRCh37/hg19 coordinates). For eQTL mapping, SNPs were mapped to genes if they were significantly associated with gene expression in human brain tissues (FDR < 0.05) based on data from GTEx v8.

We used MAGMA v1.10 (implemented in FUMA) to perform gene-based association testing, aggregating SNP-level p-values into gene-level statistics using the SNP-wise mean model. SNPs were assigned to 19,631 protein-coding genes based on NCBI 37.3 gene definitions. Bonferroni correction was applied to correct for multiple testing, with the significance threshold set at P < 2.547×10^−6^ (0.05/19,631 genes).

To identify shared biological pathways, we performed gene set enrichment using 17,023 curated gene sets and Gene Ontology (GO) terms from the Molecular Signatures Database MsigDB v2023.1Hs in MAGMA, and results were Bonferroni-corrected. To identify tissue-specific expression, we used MAGMA gene-property analysis for 30 general tissue types and 53 specific tissues from GTEx v8, correcting for multiple tests.

To examine cell-type enrichment, we identified genes uniquely associated with adolescent-onset depression by selecting those that reached significance in our MAGMA gene-based analysis but did not overlap with ‘high confidence’ genes identified in lifetime major depression^24^. Single nucleus RNA sequencing (snRNA-seq) data were obtained^28^ (GEO: GSE168408). Filtered feature HDF5 files for adolescent donors (aged ≥10 and <20 years), along with metadata, were imported, integrated and analysed using Seurat v5. Cells of poor quality and those labelled as developmental were excluded from downstream analyses. Statistical comparisons reported in **Supplementary Tables 14-15** were conducted using the Wilcoxon rank-sum test.

### Treatment target analysis

We focused on the genes identified by MAGMA and sought to identify medicines associated with these genes, leveraging datasets from the DrugBank 6.0^27^. We mapped genes to molecular entities and then used entities to identify medicines using the ‘bonds’ table. The data contains information for pairwise binding relationships between medicines and biological entities (mainly proteins, see details of the table in the URL: https://docs.drugbank.com/csv/#bonds). The binding relationships include: 1) targets of medicine actions, 2) enzymes that are inhibited, induced, or involved in medicine metabolism, and 3) & 4) carrier or transporter proteins that take part in the transportation of medicines across biological membranes.

Proteins encoded by the genes for adolescent-onset MD were listed as biological entities. Ensembl annotation was used to map genes and proteins in the human genome (’biomaRt’ R package version 2.64.0). Six entities were mapped to the targetable genome listed in the database. We identified all the medicines associated with the entities, which were annotated according to the Anatomical Therapeutic Chemical (ATC) classification using the ‘drugs’ table from DrugBank 6.0 (see details in the URL: https://docs.drugbank.com/csv/#drugs).

### LD genetic correlations

We estimated genetic correlations (*r_g_*) using LDSC^31^. Genetic correlations were calculated between each stratified ascertainment type for the European ancestry meta-analysis (prospective, retrospective, diagnosis), and between the meta-analysis and each ascertainment type for a range of other traits. We selected psychiatric and non-psychiatric traits based on reports of associations within the literature (**Supplementary Table 17**). We applied the false discovery rate (FDR) with *q* < 0.05 to correct for multiple testing.

### Mendelian randomization (MR)

We performed two-sample Mendelian randomization (MR) to examine potentially causal relationships between traits and adolescent-onset depression in the European ancestry meta-analysis^81^. Further details regarding the assumptions and criteria for MR are given here^82^. MR uses genetic variation as instruments to examine causal relationships, with two-sample MR using only GWAS summary statistics. We selected multiple biological and socially driven traits that have been shown to be either risk factors for or consequences of adolescent depression. We examined bi-directional relationships by estimating the effect of these traits on adolescent-onset depression and then of adolescent-onset depression on these traits. We used the *TwoSampleMR* v0.6.17 R package^32^ primarily relying on the inverse-variance-weighted (IVW) regression method. Associations were considered putatively causal if the IVW estimate passed FDR correction, both the weighted-median test (WMedian)^83^ and weighted-mode (WMode)^84^ estimates were concordant in direction with the IVW estimate and the MR Egger intercept was not significant from the null, suggesting minimal bias from directional horizontal pleiotropy^85^.

Due to potential bias from sample overlap in MR ^86^, we performed MR analyses using instruments from UK Biobank (UKB) only and removed UKB from our adolescent-onset depression GWAS. We selected traits that have previously been associated with adolescent depression in observational and MR analyses or postulated to be associated with adolescent depression^3,6,9,61,63,87^. We obtained UKB GWAS summary statistics for various traits from the MRC IEU OpenGWAS data repository^88^, these are given in **Supplementary Table 18**. When examining all traits as exposures, we restricted to independent variants using the ‘clump_data’ function with an *r*^2^ < 0.001, to avoid bias due to linkage disequilibrium. We then used the ‘harmonise_data’ function to ensure genetic variants were available in both the exposure and outcome data and that alleles were aligned.

### Polygenic score (PGS) analyses

#### PGS out-of-sample prediction

Polygenic scores (PGSs) were evaluated in two multi-ancestry internal validation cohorts: ABCD (N = 10,554) and Add Health (N = 9,121) (cohort details in **Supplementary Note**).

In ABCD, individuals were assigned to ancestry groups (European, African, American-admixed, East Asian) using the first four principal components (PCs) from 1000 Genomes reference panel superpopulations via a random forest classifier. Genotyped participants included 2,071 African, 2,523 American-admixed, and 5,950 European ancestry individuals; the East Asian ancestry group (N = 94) was excluded due to small sample size.

In Add Health, PCs were generated using EIGENSTRAT in an unrelated subset. The remaining participants and HapMap3 references were projected into this space. The sample included 1,971 African, 986 American-admixed, 437 East Asian, and 5,727 European ancestry individuals. The East Asian group was again excluded from PGS analyses (N = 437).

PGS were derived from both European ancestry-only and cross-ancestry meta-analyses. As MR-MEGA (used for cross-ancestry discovery) does not produce SNP effect sizes, we conducted an additional inverse-variance weighted meta-analysis in METAL to obtain summary statistics for PGS derivation (**Supplementary Figure 16**). Each PGS was computed using a leave-one-out approach to avoid overlap between discovery and target samples.

PGS construction used SBayesRC (GCTB v2.5.2), incorporating functional annotations and high-density SNPs ^33^. Ancestry-specific LD reference panels were used for European, African, and East Asian ancestries (UK Biobank), along with S-LDSC BaselineLD v2.2 annotations^89^. Due to lack of an American-admixed LD panel, the European panel was used as a proxy^90^. PGS were generated using SNP weights from SBayesRC and target genotype data via PLINK v1.9 (https://www.cog-genomics.org/plink/1.9/).

Finally, within each ancestry and cohort, logistic regression was used to assess PGS association with case-control status, adjusting for the first 4 genetic PCs and sex. Results were meta-analysed across cohorts using inverse-variance weighted fixed-effects models (*meta* R package). To calculate the incremental increase in variance explained by the PGS, we ran both a full model (PGS + PCs + sex) and a covariate only model (PCs + sex) using linear regression. We then calculated the ΔR² between these models and converted it to the liability scale (ΔR²_liability_)^79^ assuming a population prevalence of 15%.

### PGS association with structural brain phenotypes

The ACBD-US study collects imaging data capturing measures of brain development across childhood and adolescence (further cohort detail in the **Supplementary Note**). We tested for association between adolescent-onset depression PGS with 5 global and 158 regional structural brain phenotypes in ABCD-US study participants. Brain imaging data were acquired and processed by the ABCD-US team. Data was acquired using a 3-T Siemens Prisma, General Electric 750 or Philips scanner and data were harmonised between sites and scanners using a unified protocol. Quality control and data preprocessing steps have been described in detail previously^58,91^. Global features included cortical volume, thickness and surface area, and white matter tract fractional anisotropy (FA) and mean diffusivity (MD). Regional imaging features included: bilateral cortical thickness (N = 34), bilateral cortical surface area (N = 34), bilateral cortical volume (N = 34), bilateral subcortical volume (N = 12), unilateral subcortical features (N = 10), bilateral FA white matter tracts (N = 14), bilateral MD white matter tracts (N = 14), unilateral FA white matter tracts (N = 3), bilateral MD white matter tracts (N = 3).

We evaluated associations between polygenic risk for adolescent-onset depression and structural brain features using SBayesRC-derived PGS from the cross-ancestry meta-analysis, in a subset of 7,490 individuals with baseline imaging data (aged 9–10 years). Linear regression analyses were conducted using the *statsmodels* Python package (v0.14.4), with PGS as the predictor and brain phenotypes as outcomes, applying a two-sided significance threshold of *P* < .05.

For global and unilateral brain features, we used general linear models (GLMs) including covariates for biological sex, age, age², and imaging device ID. For bilateral features, we applied repeated-measures linear models with the same covariates and hemisphere included as a repeated measure. We also tested for PGS-by-hemisphere interaction effects; for structures showing significant interaction (FDR-corrected *q* < 0.05), lateralised GLMs were run separately for each hemisphere. All brain phenotypes were standardised (zero mean, unit variance), allowing effect sizes to be interpreted as standardised beta coefficients. Global measures were tested first, followed by regional analyses, with multiple comparisons controlled using FDR correction (*q* < 0.05).

### PGS association with adolescent psychiatric phenotypes in ALSPAC

We tested associations between the adolescent-onset depression PGS (derived from a leave-ALSPAC-out version of the European ancestry meta-analysis) with a range of adolescent psychiatric phenotypes from the Avon Longitudinal Study of Parents and Children (ALSPAC), a longitudinal and prospectively assessed cohort in the United Kingdom (cohort information in the **Supplementary Note**). The PGS in ALSPAC was created using SBayesRC as described above and was standardised to have a mean of 0 and a standard deviation of 1. We explored associations between the adolescent-onset depression PGS and the following phenotypes: 1) a diagnosis of depression measured through the Clinical Interview Schedule – Revised (CISR)^92^ assessed at around age 18, 2) self-reported depressive symptoms, assessed through the short mood and feelings questionnaire (SMFQ)^69^ around age 18, 3) the number of depression episodes, assessed by the number of times an individual surpassed the clinical cutoff of the SMFQ (>=11) between the ages of 10 and 19 years (max 6 occasions), 4) a diagnosis of anxiety measured through the CISR around age 18, 5) a history of self-reported self-harm up to the age of 18, 6) presence of suspected or confirmed psychosis like experiences assessed through the Psychotic Like-Symptoms Interview (PLIKS)^93^ assessed around age 18, 7) self-reported daily smoking assessed around the age of 18 and 8), and self-reported alcohol use measured by the Alcohol Use Disorders Identification Test (AUDIT) ^94^ around age 18.

Leveraging ALSPACs key feature of repeated assessments of depression, we also examined how the adolescent-onset PGS was associated with trajectories of depressive symptoms from childhood through early adulthood. We estimated trajectories of depressive symptoms using the clinical cutoff of the SMFQ (>=11) for up to 11 occasions between the ages of 10 and 28. We restricted our analytical sample to those who had at least 1 occasion of the SMFQ between the ages of 10 and 18 and between the ages of 19 and 28. We used latent class growth analysis (LCGA)^95^ to estimate trajectories using MPlus v8.11. To model the trajectories, we used a linear piecewise model accounting for two developmental phases: late childhood to mid adolescence and then mid adolescence to early adulthood, as per previous research in this cohort^87^. Missing data were handled using Full Information Maximum Likelihood. We estimated trajectories (or latent classes) comparing 1 through 6 class solutions and selected the best trajectories based upon several established criteria including entropy, likelihood ratio tests, class size and interpretation (**Supplementary Figure 17, Supplementary Table 21**). We used multinomial logistic regression to examine the association between the trajectories and the adolescent-onset depression PGS, adjusting for sex (coded as 0 for female, 1 for males) and the first 10 principal components for ancestry. Our main analysis used the low symptom-level trajectory as the reference category, but in sensitivity analysis, we changed the reference trajectory to allow comparisons between more at risk trajectories (**Supplementary Table 21**). Genotyping information and quality control for ALSPAC have been previously described^62^, with a summary in **Supplementary Table 2**.

### PGS association with timing of pubertal onset in ALSPAC

We tested association of adolescent-onset depression PGS (derived from the European ancestry meta-analysis and the SBayesRC methods as per above) with markers of pubertal timing in ALSPAC, stratified by sex. We used prospectively assessed markers of age of menarche as the pubertal marker in females and age at peak height velocity in males (i.e., the age where height is increasing at the fastest rate). Further details of how these phenotypes were derived have been reported previously^96^. We performed a linear regression adjusting for the first 10 principal components of ancestry.

Summary statistics will be made available on the PGC data-download page (https://pgc.unc.edu/for-researchers/download-result).

## Acknowledgements

Study specific acknowledgements and funding are in the **Supplementary Note**.

## Consortia and group authors

The members of the Major Depressive Disorder Working Group of the Psychiatric Genomics Consortium (PGC-MDD) are Mark J. Adams, Fabian Streit, Xiangrui Meng, Swapnil Awasthi, Brett N. Adey, Karmel W. Choi, V. Kartik Chundru, Jonathan R.I. Coleman, Bart Ferwerda, Jerome C. Foo, Zachary F. Gerring, Olga Giannakopoulou, Priya Gupta, Alisha S.M. Hall, Arvid Harder, David M. Howard, Christopher Hübel, Alex S.F. Kwong, Daniel F. Levey, Brittany L. Mitchell, Guiyan Ni, Vanessa K. Ota, Oliver Pain, Gita A. Pathak, Eva C. Schulte, Xueyi Shen, Jackson G. Thorp, Alicia Walker, Shuyang Yao, Jian Zeng, Johan Zvrskovec, Dag Aarsland, Ky’Era V. Actkins, Mazda Adli, Esben Agerbo, Mareike Aichholzer, Allison Aiello, Tracy M. Air, Thomas D. Als, Evelyn Andersson, Till F.M. Andlauer, Volker Arolt, Helga Ask, Julia Bäckman, Sunita Badola, Clive Ballard, Karina Banasik, Nicholas J. Bass, Aartjan T.F. Beekman, Sintia Belangero, Tim B. Bigdeli, Elisabeth B. Binder, Ottar Bjerkeset, Gyda Bjornsdottir, Sigrid Børte, Emma Bränn, Alice Braun, Thorsten Brodersen, Tanja M. Brückl, Søren Brunak, Mie T. Bruun, Margit Burmeister, Pichit Buspavanich, Jonas Bybjerg-Grauholm, Enda M. Byrne, Jianwen Cai, Archie Campbell, Megan L. Campbell, Adrian I. Campos, Enrique Castelao, Jorge Cervilla, Boris Chaumette, Chia-Yen Chen, Hsi-Chung Chen, Zhengming Chen, Sven Cichon, Lucía Colodro-Conde, Anne Corbett, Elizabeth C. Corfield, Baptiste Couvy-Duchesne, Nick Craddock, Udo Dannlowski, Gail Davies, E.J.C. de Geus, Ian J. Deary, Franziska Degenhardt, Abbas Dehghan, J. Raymond DePaulo, Michael Deuschle, Maria Didriksen, Khoa Manh Dinh, Nese Direk, Srdjan Djurovic, Anna R. Docherty, Katharina Domschke, Joseph Dowsett, Ole Kristian Drange, Erin C. Dunn, William Eaton, Gudmundur Einarsson, Thalia C. Eley, Samar S.M. Elsheikh, Jan Engelmann, Michael E. Benros, Christian Erikstrup, Valentina Escott-Price, Chiara Fabbri, Yu Fang, Sarah Finer, Josef Frank, Robert C. Free, Linda Gallo, He Gao, Michael Gill, Maria Gilles, Fernando S. Goes, Scott Douglas Gordon, Poppy Z. Grimes, Jakob Grove, Daniel F. Gudbjartsson, Blanca Gutierrez, Tim Hahn, Lynsey S. Hall, Thomas F. Hansen, Magnus Haraldsson, Catharina A. Hartman, Alexandra Havdahl, Caroline Hayward, Stefanie Heilmann-Heimbach, Stefan Herms, Ian B. Hickie, Henrik Hjalgrim, Jens Hjerling-Leffler, Per Hoffmann, Georg Homuth, Carsten Horn, Jouke-Jan Hottenga, David M. Hougaard, Iiris Hovatta, Qin Qin Huang, Donald Hucks, Floris Huider, Karen A. Hunt, Nicholas S. Ialongo, Marcus Ising, Erkki Isometsä, Rick Jansen, Yunxuan Jiang, Ian Jones, Lisa A. Jones, Lina Jonsson, Masahiro Kanai, Robert Karlsson, Siegfried Kasper, Kenneth S. Kendler, Ronald C. Kessler, Stefan Kloiber, James A. Knowles, Nastassja Koen, Julia Kraft, Henry R. Kranzler, Kristi Krebs, Theodora Kunovac Kallak, Zoltán Kutalik, Elisa Lahtela, Marilyn Lake, Margit Hørup Larsen, Eric J. Lenze, Melissa Lewins, Glyn Lewis, Liming Li, Bochao Danae Lin, Kuang Lin, Penelope A. Lind, Yu-Li Liu, Donald J. MacIntyre, Dean F. MacKinnon, Brion S. Maher, Wolfgang Maier, Victoria S. Marshe, Gabriela A. Martinez-Levy, Koichi Matsuda, Hamdi Mbarek, Peter McGuffin, Sarah E. Medland, Susanne Meinert, Christina Mikkelsen, Susan Mikkelsen, Yuri Milaneschi, Iona Y. Millwood,Esther Molina, Francis M. Mondimore, Preben Bo Mortensen, Benoit H. Mulsant, Joonas Naamanka, Jake M. Najman, Matthias Nauck, Igor Nenadić, Kasper R. Nielsen, Ilja M. Nolt, Merete Nordentoft, Markus M. Nöthen, Mette Nyegaard, Michael C. O’Donovan, Asmundur Oddsson, Adrielle M. Oliveira, Catherine M. Olsen, Hogni Oskarsson, Sisse Rye Ostrowski, Michael J. Owen, Richard Packer, Teemu Palviainen, Pedro M. Pan, Carlos N. Pato, Michele T. Pato, Nancy L. Pedersen, Ole Birger Pedersen, Wouter J. Peyrot, James B. Potash, Martin Preisig, Michael H. Preuss, Jorge A. Quiroz, Miguel E. Renteria, Charles F. Reynolds III, John P. Rice, Saori Sakaue, Marcos L. Santoro, Robert A. Schoevers, Andrew Schork, Thomas G. Schulze, Tabea S. Send, Jianxin Shi, Engilbert Sigurdsson, Kritika Singh, Grant C.B. Sinnamon, Lea Sirignano, Olav B. Smeland, Daniel J. Smith, Tamar Sofer, Erik Sørensen, Sundararajan Srinivasan, Hreinn Stefansson, Kari Stefansson, Peter Straub, Mei-Hsin Su, André Tadic, Henning Teismann, Alexander Teumer, Anita Thapar, Pippa A. Thomson, Lise Wegner Thørner, Apostolia Topaloudi, Shih-Jen Tsai, Ioanna Tzoulaki, George Uhl, André G. Uitterlinden, Henrik Ullum, Daniel Umbricht, Robert J. Ursano, Sandra Van der Auwera, Albert M. van Hemert, Abirami Veluchamy, Alexander Viktorin, Henry Völzke, G. Bragi Walters, Xiaotong Wang, Agaz Wani, Myrna M. Weissman, Jürgen Wellmann, David C. Whiteman, Derek Wildman, Gonneke Willemsen, Alexander T. Williams, Bendik S. Winsvold, Stephanie H. Witt, Ying Xiong, Lea Zillich, John-Anker Zwart, 23andMe Research Team, China Kadoorie Biobank, Collaborative Group, Estonian Biobank Research Team, Genes & Health Research Team, HUNT All-In Psychiatry, The BioBank Japan Project, VA Million Veteran Program, Ole A. Andreassen, Bernhard T. Baune, Klaus Berger, Dorret I. Boomsma, Anders D. Børglum, Gerome Breen, Na Cai, Hilary Coon, William E. Copeland, Byron Creese, Carlos S. Cruz-Fuentes, Darina Czamara, Lea K. Davis, Eske M. Derks, Enrico Domenici, Paul Elliott, Andreas J. Forstner, Micha Gawlik, Joel Gelernter, Hans J. Grabe, Steven P. Hamilton, Kristian Hveem, Catherine John, Jaakko Kaprio, Tilo Kircher, Marie-Odile Krebs, Po-Hsiu Kuo, Mikael Landén, Kelli Lehto, Douglas F. Levinson, Qingqin S. Li, Klaus Lieb, Ruth J.F. Loos, Yi Lu, Susanne Lucae, Jurjen J. Luykx, Hermine H.M. Maes, Patrik K. Magnusson, Hilary C. Martin, Nicholas G. Martin, Andrew McQuillin, Christel M. Middeldorp, Lili Milani, Ole Mors, Daniel J. Müller, Bertram Müller-Myhsok, Yukinori Okada, Albertine J. Oldehinkel, Sara A. Paciga, Colin N.A. Palmer, Peristera Paschou, Brenda W.J.H. Penninx, Roy H. Perlis, Roseann E. Peterson, Giorgio Pistis, Renato Polimanti, David J. Porteous, Danielle Posthuma, Jill A. Rabinowitz, Ted Reichborn-Kjennerud, Andreas Reif, Frances Rice, Roland Ricken, Marcella Rietschel, Margarita Rivera, Christian Rück, Giovanni A. Salum, Catherine Schaefer, Srijan Sen, Alessandro Serretti, Alkistis Skalkidou, Jordan W. Smoller, Dan J. Stein, Frederike Stein, Murray B. Stein, Patrick F. Sullivan, Martin Tesli, Thorgeir E. Thorgeirsson, Henning Tiemeier, Nicholas J. Timpson, Monica Uddin, Rudolf Uher, David A. van Heel, Karin J.H. Verweij, Robin G. Walters, Sylvia Wassertheil-Smoller, Jens R. Wendland, Thomas Werge, Aeilko H. Zwinderman, Karoline Kuchenbaecker, Naomi R. Wray, Stephan Ripke, Cathryn M. Lewis, and Andrew M. McIntosh.

## GLAD+ group author

Gerome Breen, Thalia C. Eley, Matthew Hotopf, Chérie Armour, Ian R. Jones, Andrew M. McIntosh, James T. R. Walters, Daniel J. Smith, Jonathan R. I. Coleman, Allan H. Young, Antony J. Cleare, Katrina A. S. Davis, Georgina Krebs, Katharine A. Rimes, David Veale, Roland Zahn, Molly R. Davies, Gursharan Kalsi, Shannon Bristow, Susannah C. B. Curzons, Henry C. Rogers, Katherine N. Thompson, Brett N. Adey, Ian Marsh, Dina Monssen, Monika McAtarsney-Kovacs, Charles J. Curtis, Jahnavi Arora, Saakshi Kakar, Laura Meldrum, Iona Smith, Le Roy Dowey, Victor Gault, Donald M. Lyall, Ruth K. Price, Keith G. Thomas, Zain Ahmad, Helena L. Davies, Christopher Hübel, Sang Hyuck Lee, Abigail R. ter Kuile, Danyang Li, Yuhao (Leo) Lin, Jared G. Maina, Jessica Mundy, Alish Palmos, Alicia J. Peel, Kirstin Purves, Christopher Rayner, Megan Skelton, Rujia Wang, Johan Zvrskovec.

## NIHR BioResource group author

John Bradley, Nathalie Kingston, Hannah Stark, Carola Kanz, Alexei Moulton, Nigel Ovington, Jacinta Lee, Debbie Clapham-Riley, Katie Mills.

## Author contributions

P.Z. Grimes and A.S.F. Kwong proposed and designed the study. P.Z. Grimes wrote the manuscript and carried out all meta-analyses and downstream analyses in the paper except for: the Mendelian randomization and ALSPAC PGS analyses carried out by A.S.F. Kwong, the Add Health PGC analysis carried out by K.N. Thompson, the brain phenotype association analysis carried out by Q. Deng, the drug bank analysis conducted by X. Shen and the cell expression analysis conducted by J. Wolfe. J.T. Thomas developed and provided the SNP reference file and liftover script. R. Wootton provided analytical support for the MR analysis. A.S.F Kwong, H.C. Whalley, B.L. Mitchell, Y. Lu, C. Middeldorp, M.J Adams, A. McIntosh, and H. Larsson helped to conceive the experimental design and supervised the project. All authors provided comments on the manuscript and approved the final version.

Analysts for the contributing cohorts were as follows: **ABCD** (US) = P.Z. Grimes, **ABCD** (Netherlands) = E. Motazedi, **Add Health** (US) = K.N. Thompson, Y. Jeong, S. Arirangan, D.E Adkins, **AGDS** (Australia) = P.Z. Grimes, J.T. Thomas, **ALSPAC** (UK) = P.Z. Grimes, **BHRC** (Brazil) = V.K. Ota, **BLTS** (Australia) = P.Z. Grimes, **CATSS** (Sweden) = T-D. Nguyen, **GEDI** (US) = R. Peterson and C. Chatzinakos, **GenR** (Netherlands) = M. Shahisavandi, **GenScot** (UK) = P.Z. Grimes and M.J Adams, **GLAD** (UK) = E. Assary, **GUSTO** (Singapore) = H. Pan, **INSchool** (Spain) = U. Zubizarreta-Arruti, **Janssen** (US) = Q. Li, **Lifelines** (Netherlands) = C. Slaney, **MUSP** (Australia) = S. Hassan Gangaraju, **MCS** (UK) = A. Shakeshaft and C. Dennison, **MoBa** (Norway) = R. Wootton, **Nordic early-onset** = J.R. Shorter, J.A. Pasman, S. Kurvits, A. Jangmo, J. Naamanka, **PGC cohorts** = P.Z. Grimes, **Raine Study** (Australia) = C. Wang, **TEDS** (UK) = E. Assary, **TRAILS** (Netherlands) = I. Nolte, **UKBiobank** = P.Z. Grimes and M.J. Adams.

Contribution of data and/or supervision of analysts was provided by the following people for each cohort: **ABCD** (US) = H.C. Whalley, **ABCD** (Netherlands) = T. Vrijkotte, Add Health (US) = R. Wedow, **AGDS** (Australia) = N. Martin, and I.Hickie, **ALSPAC** (UK) = A.S.F. Kwong, **BHRC** (Brazil) = P.M. Pan, S.I. Belangero, G.A. Salum, **BLTS** (Australia) = N. Martin, **CATSS** (Sweden) = Y. Lu, **GEDI** (US) = H. Maes, W. Copeland, and R. Peterson, **GenR** (Netherlands) = A. Neumann, **GenScot** (UK) = H.C. Whalley, **GLAD** (UK) = G. Breen and T.C. Eley, **GUSTO** (Singapore) = A. Rodriguez, H.Pan, **INSchool** (Spain) = S. Alemany, R. Bosch, M. Casas, **Janssen** (US) = Q. Li, **Lifelines** (Netherlands) = C. Hartman and G. Khandaker, **MUSP** (Australia) = E. Byrne, **MCS** (UK) = A. Thapar, **MoBa** (Norway) = A. Havdahl, H. Ask, L. Hannigan, **Nordic early-onset** = Y. Lu, A. Buil, K. Lehto, M. Tesli, **PGC cohorts** = A. McIntosh, **Raine Study** (Australia) = A.J.O. Whitehouse, C. Pennell, **TEDS** (UK) = T.C. Eley, **TRAILS** (Netherlands) = C. Hartman, A.J. Oldehinkel, **UKBiobank** = A. McIntosh.

## References

1. Sawyer, S. M., Azzopardi, P. S., Wickremarathne, D. & Patton, G. C. The age of adolescence. The Lancet Child & Adolescent Health 2, 223–228 (2018).

2. Solmi, M. et al. Age at onset of mental disorders worldwide: large-scale meta-analysis of 192 epidemiological studies. Mol Psychiatry 27, 281–295 (2022).

3. Thapar, A., Collishaw, S., Pine, D. S. & Thapar, A. K. Depression in adolescence. Lancet 379, 1056–1067 (2012).

4. World Health Organization. Mental health of adolescents. https://www.who.int/news-room/fact-sheets/detail/adolescent-mental-health (2024).

5. Johnson, D., Dupuis, G., Piche, J., Clayborne, Z. & Colman, I. Adult mental health outcomes of adolescent depression: A systematic review. Depression and Anxiety 35, 700–716 (2018).

6. Clayborne, Z. M., Varin, M. & Colman, I. Systematic Review and Meta-Analysis: Adolescent Depression and Long-Term Psychosocial Outcomes. Journal of the American Academy of Child & Adolescent Psychiatry 58, 72–79 (2019).

7. Copeland, W. E., Alaie, I., Jonsson, U. & Shanahan, L. Associations of Childhood and Adolescent Depression With Adult Psychiatric and Functional Outcomes. Journal of the American Academy of Child & Adolescent Psychiatry 60, 604–611 (2021).

8. Zisook, S. et al. Effect of age at onset on the course of major depressive disorder. Am J Psychiatry 164, 1539–1546 (2007).

9. Thapar, A., Eyre, O., Patel, V. & Brent, D. Depression in young people. Lancet 400, 617–631 (2022).

10. World Health Organization. World mental health today: Latest data. (2025).

11. Lu, W. Adolescent Depression: National Trends, Risk Factors, and Healthcare Disparities. Am J Health Behav 43, 181–194 (2019).

12. McIntosh, A. M., Sullivan, P. F. & Lewis, C. M. Uncovering the Genetic Architecture of Major Depression. Neuron 102, 91–103 (2019).

13. Nguyen, T.-D. et al. Genetic heterogeneity and subtypes of major depression. Mol Psychiatry 27, 1667–1675 (2022).

14. Polderman, T. J. C. et al. Meta-analysis of the heritability of human traits based on fifty years of twin studies. Nat Genet 47, 702–709 (2015).

15. Nguyen, T.-D. et al. Genetic Contribution to the Heterogeneity of Major Depressive Disorder: Evidence From a Sibling-Based Design Using Swedish National Registers. Am J Psychiatry 180, 714–722 (2023).

16. Shorter, J. et al. Genome-wide association study using Nordic biobank and longitudinal health registry reveals different genetic architecture between early- and late-onset depression. Nat Genet (In press).

17. Pfeifer, J. H. & Allen, N. B. Puberty Initiates Cascading Relationships Between Neurodevelopmental, Social, and Internalizing Processes Across Adolescence. Biological Psychiatry 89, 99–108 (2021).

18. Bethlehem, R. a. I., et al. Brain charts for the human lifespan. Nature 604, 525–533 (2022).

19. Harder, A. et al. Genetics of age-at-onset in major depression. Transl Psychiatry 12, 1–7 (2022).

20. Power, R. A. et al. Genome-wide Association for Major Depression Through Age at Onset Stratification: Major Depressive Disorder Working Group of the Psychiatric Genomics Consortium. Biol Psychiatry 81, 325–335 (2017).

21. Jami, E. S. et al. Genome-wide Association Meta-analysis of Childhood and Adolescent Internalizing Symptoms. Journal of the American Academy of Child & Adolescent Psychiatry 61, 934–945 (2022).

22. Sallis, H. et al. Genetics of depressive symptoms in adolescence. BMC Psychiatry 17, 321 (2017).

23. Peterson, R. E. et al. Genome-wide Association Studies in Ancestrally Diverse Populations: Opportunities, Methods, Pitfalls, and Recommendations. Cell 179, 589–603 (2019).

24. Adams, M. J. et al. Trans-ancestry genome-wide study of depression identifies 697 associations implicating cell types and pharmacotherapies. Cell 0, (2025).

25. Grotzinger, A. D. et al. Genomic structural equation modelling provides insights into the multivariate genetic architecture of complex traits. Nat Hum Behav 3, 513–525 (2019).

26. Watanabe, K., Taskesen, E., van Bochoven, A. & Posthuma, D. Functional mapping and annotation of genetic associations with FUMA. Nat Commun 8, 1826 (2017).

27. Knox, C. et al. DrugBank 6.0: the DrugBank Knowledgebase for 2024. Nucleic Acids Res 52, D1265–D1275 (2024).

28. Herring, C. A. et al. Human prefrontal cortex gene regulatory dynamics from gestation to adulthood at single-cell resolution. Cell 185, 4428–4447.e28 (2022).

29. Du, H. et al. Transcription factors Bcl11a and Bcl11b are required for the production and differentiation of cortical projection neurons. Cereb Cortex 32, 3611–3632 (2022).

30. Lennon, M. J., Jones, S. P., Lovelace, M. D., Guillemin, G. J. & Brew, B. J. Bcl11b—A Critical Neurodevelopmental Transcription Factor—Roles in Health and Disease. Front. Cell. Neurosci. 11, (2017).

31. Bulik-Sullivan, B. K. et al. LD Score regression distinguishes confounding from polygenicity in genome-wide association studies. Nat Genet 47, 291–295 (2015).

32. Hemani, G. et al. The MR-Base platform supports systematic causal inference across the human phenome. eLife 7, e34408 (2018).

33. Zheng, Z. et al. Leveraging functional genomic annotations and genome coverage to improve polygenic prediction of complex traits within and between ancestries. Nat Genet 56, 767–777 (2024).

34. Mägi, R. et al. Trans-ethnic meta-regression of genome-wide association studies accounting for ancestry increases power for discovery and improves fine-mapping resolution. Human Molecular Genetics 26, 3639–3650 (2017).

35. Mitchell, B. L. et al. The Australian Genetics of Depression Study: New Risk Loci and Dissecting Heterogeneity Between Subtypes. Biological Psychiatry 92, 227–235 (2022).

36. Wray, N. R. et al. Genome-wide association analyses identify 44 risk variants and refine the genetic architecture of major depression. Nat Genet 50, 668–681 (2018).

37. García-Aznar, J. M., Alonso Alvarez, S. & Bernal del Castillo, T. Pivotal role of BCL11B in the immune, hematopoietic and nervous systems: a review of the BCL11B-associated phenotypes from the genetic perspective. Genes Immun 25, 232–241 (2024).

38. Kikuchi, K. et al. Map7D2 and Map7D1 facilitate microtubule stabilization through distinct mechanisms in neuronal cells. Life Science Alliance 5, (2022).

39. Koizumi, H. et al. DCLK1 phosphorylates the microtubule-associated protein MAP7D1 to promote axon elongation in cortical neurons. Developmental Neurobiology 77, 493–510 (2017).

40. Breiderhoff, T. et al. Sortilin-Related Receptor SORCS3 Is a Postsynaptic Modulator of Synaptic Depression and Fear Extinction. PLOS ONE 8, e75006 (2013).

41. Christiansen, G. B. et al. The sorting receptor SorCS3 is a stronger regulator of glutamate receptor functions compared to GABAergic mechanisms in the hippocampus. Hippocampus 27, 235–248 (2017).

42. Kamran, M. et al. Independent Associated SNPs at SORCS3 and Its Protein Interactors for Multiple Brain-Related Disorders and Traits. Genes 14, 482 (2023).

43. Wu, B. et al. BPTF Is Essential for T Cell Homeostasis and Function. The Journal of Immunology 197, 4325–4333 (2016).

44. Stankiewicz, P. et al. Haploinsufficiency of the Chromatin Remodeler BPTF Causes Syndromic Developmental and Speech Delay, Postnatal Microcephaly, and Dysmorphic Features. The American Journal of Human Genetics 101, 503–515 (2017).

45. Monistrol-Mula, A. et al. Genetic analyses point to alterations in immune-related pathways underpinning the association between psychiatric disorders and COVID-19. Mol Psychiatry 30, 29–36 (2025).

46. Khandaker, G. M., Pearson, R. M., Zammit, S., Lewis, G. & Jones, P. B. Association of Serum Interleukin 6 and C-Reactive Protein in Childhood With Depression and Psychosis in Young Adult Life: A Population-Based Longitudinal Study. JAMA Psychiatry 71, 1121–1128 (2014).

47. Milaneschi, Y. et al. Association of inflammation with depression and anxiety: evidence for symptom-specificity and potential causality from UK Biobank and NESDA cohorts. Mol Psychiatry 26, 7393–7402 (2021).

48. Nair, P. C., McKinnon, R. A., Miners, J. O. & Bastiampillai, T. Binding of clozapine to the GABAB receptor: clinical and structural insights. Mol Psychiatry 25, 1910–1919 (2020).

49. Gao, L. et al. Association between Dietary Theobromine and Cognitive Function in a Representative American Population: A Cross-Sectional Study. The Journal of Prevention of Alzheimer’s Disease 9, 449–457 (2022).

50. Duncan, L. et al. Analysis of polygenic risk score usage and performance in diverse human populations. Nat Commun 10, 3328 (2019).

51. Martin, A. R. et al. Clinical use of current polygenic risk scores may exacerbate health disparities. Nat Genet 51, 584–591 (2019).

52. Kanjira, S. C. et al. Polygenic prediction of major depressive disorder and related traits in African ancestries UK Biobank participants. Mol Psychiatry 30, 151–157 (2025).

53. Okewole, A., et al. Depression Genetics in Africa (DepGenAfrica): protocol for the first large-scale case-control study of major depressive disorder among continental Africans. Preprint at 10.31219/osf.io/pf3u9_v1 (2025).

54. Lewis, C. M. & Vassos, E. Polygenic risk scores: from research tools to clinical instruments. Genome Med 12, 44 (2020).

55. Xu, E. Y., Grimes, P. Z., Kwong, A. S. F., Lawrie, S. M. & Whalley, H. C. Longitudinal Prediction of Adolescent Depression from Environmental and Polygenic Risk Scores. 2025.07.08.25331098 Preprint at 10.1101/2025.07.08.25331098 (2025).

56. Whittle, S. et al. Structural Brain Development and Depression Onset During Adolescence: A Prospective Longitudinal Study. AJP 171, 564–571 (2014).

57. Barch, D. M. et al. White matter alterations associated with lifetime and current depression in adolescents: Evidence for cingulum disruptions. Depression and Anxiety 39, 881–890 (2022).

58. Shen, X. et al. Brain structural associations with depression in a large early adolescent sample (the ABCD study®). eClinicalMedicine 42, (2021).

59. Thapar, A., Oginni, O., Dennison, C. A. & Rice, F. Youth depression: An overview of genetic findings and the challenge of heterogeneity. Journal of Affective Disorders 391, 120049 (2025).

60. Grimes, P. Z. et al. Genetic Architectures of Adolescent Depression Trajectories in 2 Longitudinal Population Cohorts. JAMA Psychiatry (2024) doi:10.1001/jamapsychiatry.2024.0983.

61. Kwong, A. S. F. et al. Genetic and Environmental Risk Factors Associated With Trajectories of Depression Symptoms From Adolescence to Young Adulthood. JAMA Network Open 2, e196587 (2019).

62. Kwong, A. S. F. et al. Polygenic risk for depression, anxiety and neuroticism are associated with the severity and rate of change in depressive symptoms across adolescence. Journal of Child Psychology and Psychiatry 62, 1462–1474 (2021).

63. Askelund, A. D. et al. Assessing causal links between age at menarche and adolescent mental health: a Mendelian randomisation study. BMC Medicine 22, 155 (2024).

64. Joinson, C., Heron, J., Lewis, G., Croudace, T. & Araya, R. Timing of menarche and depressive symptoms in adolescent girls from a UK cohort. Br J Psychiatry 198, 17–23, sup 1–2 (2011).

65. Middeldorp, C. M. et al. The Early Growth Genetics (EGG) and EArly Genetics and Lifecourse Epidemiology (EAGLE) consortia: design, results and future prospects. Eur J Epidemiol 34, 279–300 (2019).

66. Achenbach, T. M. Child Behavior Checklist. in Encyclopedia of Clinical Neuropsychology (eds. Kreutzer, J. S., DeLuca, J. & Caplan, B.) 546–552 (Springer, New York, NY, 2011). doi:10.1007/978-0-387-79948-3_1529.

67. Radloff, L. S. The CES-D Scale: A self-report depression scale for research in the general population. Applied Psychological Measurement 1, 385–401 (1977).

68. Angold, A. & Costello, E. J. The Child and Adolescent Psychiatric Assessment (CAPA). J Am Acad Child Adolesc Psychiatry 39, 39–48 (2000).

69. Angold, A., Costello, E. J., Messer, S. C. & Pickles, A. Development of a short questionnaire for use in epidemiological studies of depression in children and adolescents. International Journal of Methods in Psychiatric Research 5, 237–249 (1995).

70. Goodman, R. The Strengths and Difficulties Questionnaire: A research note. Child Psychology & Psychiatry & Allied Disciplines 38, 581–586 (1997).

71. Hickie, I. B. et al. Development of a simple screening tool for common mental disorders in general practice. Med J Aust 175, S10–7 (2001).

72. Achenbach, T. M. Manual for ASEBA School-Age Forms & Profiles. *University of Vermont, Research Center for Children*, Youth & Families (2001).

73. Wittchen, H. U. Reliability and validity studies of the WHO--Composite International Diagnostic Interview (CIDI): a critical review. J Psychiatr Res 28, 57–84 (1994).

74. Sheehan, D. V. et al. The Mini-International Neuropsychiatric Interview (M.I.N.I.): the development and validation of a structured diagnostic psychiatric interview for DSM-IV and ICD-10. J Clin Psychiatry 59 **Suppl 20**, 22–33;quiz 34-57 (1998).

75. First, M. B. et al. The Structured Clinical Interview for DSM-III-R Personality Disorders (SCID-II). Part II: Multi-Site Test-Retest Reliability Study. Journal of Personality Disorders 9, 92–104 (1995).

76. Zhao, H. et al. CrossMap: a versatile tool for coordinate conversion between genome assemblies. Bioinformatics 30, 1006–1007 (2014).

77. Willer, C. J., Li, Y. & Abecasis, G. R. METAL: fast and efficient meta-analysis of genomewide association scans. Bioinformatics 26, 2190–2191 (2010).

78. Meng, X. et al. Multi-ancestry genome-wide association study of major depression aids locus discovery, fine mapping, gene prioritization and causal inference. Nat Genet 1–12 (2024) doi:10.1038/s41588-023-01596-4.

79. Lee, S. H., Goddard, M. E., Wray, N. R. & Visscher, P. M. A Better Coefficient of Determination for Genetic Profile Analysis. Genetic Epidemiology 36, 214–224 (2012).

80. Shorey, S., Ng, E. D. & Wong, C. H. J. Global prevalence of depression and elevated depressive symptoms among adolescents: A systematic review and meta-analysis. Br J Clin Psychol 61, 287–305 (2022).

81. Sanderson, E. et al. Mendelian randomization. Nat Rev Methods Primers 2, 1–21 (2022).

82. Davies, N. M., Holmes, M. V. & Davey Smith, G. Reading Mendelian randomisation studies: a guide, glossary, and checklist for clinicians. BMJ 362, k601 (2018).

83. Bowden, J., Davey Smith, G., Haycock, P. C. & Burgess, S. Consistent Estimation in Mendelian Randomization with Some Invalid Instruments Using a Weighted Median Estimator. Genet Epidemiol 40, 304–314 (2016).

84. Hartwig, F. P., Davey Smith, G. & Bowden, J. Robust inference in summary data Mendelian randomization via the zero modal pleiotropy assumption. International Journal of Epidemiology 46, 1985–1998 (2017).

85. Bowden, J., Davey Smith, G. & Burgess, S. Mendelian randomization with invalid instruments: effect estimation and bias detection through Egger regression. Int J Epidemiol 44, 512–525 (2015).

86. Burgess, S., Davies, N. M. & Thompson, S. G. Bias due to participant overlap in two-sample Mendelian randomization. Genet Epidemiol 40, 597–608 (2016).

87. Weavers, B. et al. The antecedents and outcomes of persistent and remitting adolescent depressive symptom trajectories: a longitudinal, population-based English study. The Lancet Psychiatry 8, 1053–1061 (2021).

88. Elsworth, B. et al. The MRC IEU OpenGWAS data infrastructure. Preprint at 10.1101/2020.08.10.244293 (2020).

89. Gazal, S. et al. Linkage disequilibrium dependent architecture of human complex traits shows action of negative selection. Nat Genet 49, 1421–1427 (2017).

90. Bryc, K., Durand, E. Y., Macpherson, J. M., Reich, D. & Mountain, J. L. The genetic ancestry of African Americans, Latinos, and European Americans across the United States. Am J Hum Genet 96, 37–53 (2015).

91. Casey, B. J. et al. The Adolescent Brain Cognitive Development (ABCD) study: Imaging acquisition across 21 sites. Developmental Cognitive Neuroscience 32, 43–54 (2018).

92. Lewis, G., Pelosi, A. J., Araya, R. & Dunn, G. Measuring psychiatric disorder in the community: a standardized assessment for use by lay interviewers. Psychol Med 22, 465–486 (1992).

93. Zammit, S. et al. Investigating if psychosis-like symptoms (PLIKS) are associated with family history of schizophrenia or paternal age in the ALSPAC birth cohort. Schizophrenia Research 104, 279–286 (2008).

94. Saunders, J. B., Aasland, O. G., Babor, T. F., de la Fuente, J. R. & Grant, M. Development of the Alcohol Use Disorders Identification Test (AUDIT): WHO Collaborative Project on Early Detection of Persons with Harmful Alcohol Consumption--II. Addiction 88, 791–804 (1993).

95. Jung, T. & Wickrama, K. a. S. An Introduction to Latent Class Growth Analysis and Growth Mixture Modeling. Social and Personality Psychology Compass 2, 302–317 (2008).

96. Elhakeem, A. et al. Using linear and natural cubic splines, SITAR, and latent trajectory models to characterise nonlinear longitudinal growth trajectories in cohort studies. BMC Medical Research Methodology 22, 68 (2022).

