## Supplementary Note and Figures for "Genome-wide association study of adolescent-onset depression"

**Supplementary Note (page 2) and**

**Supplementary Figures (page 14)**

**Supplementary Note**

**Cohort Descriptions, Ethical Approval Statements and Acknowledgements**

**The National Longitudinal Study of Adolescent to Adult Health (ADDHEALTH)**

The National Longitudinal Study of Adolescent to Adult Health (Add Health, <https://addhealth.cpc.unc.edu/>) is a longitudinal study of a nationally representative sample of over 20,000 adolescents who were in grades 7-12 during the 1994-95 school year, and have been followed for five waves to date, most recently in 2016-18 (K. M. Harris et al., 2019). Over the years, Add Health has collected rich demographic, social, familial, behavioural, psychosocial, cognitive, and health survey data from participants and their parents; a vast array of contextual data from participants’ schools, neighbourhoods, and geographies of residence; and in-home physical and biological data from participants, including genetic markers, blood-based assays, anthropometric measures, and medications. Ancillary studies have added even more data over the years. Data from the project are available in various forms and have been analysed in thousands of publications in peer-reviewed journals.

The Add Health phenotypic data was applied for and approved under restricted-use application #27111802. The Add Health genetic data was applied for and approved under application #38868 using the National Institutes of Health (NIH) National Center for Biotechnology Information (NCBI)'s database of Genotypes and Phenotypes (dbGaP). Use of the Add Health data at Purdue University was approved by the Purdue Institutional Review Board (IRB) under application IRB-2023-1819.

This research uses data from Add Health, funded by grant P01 HD31921 (Harris) from the Eunice Kennedy Shriver National Institute of Child Health and Human Development (NICHD), with cooperative funding from 23 other federal agencies and foundations. Add Health is currently directed by Robert A. Hummer and funded by the National Institute on Aging cooperative agreements U01 AG071448 (Hummer) and U01AG071450 (Aiello and Hummer) at the University of North Carolina at Chapel Hill. Add Health was designed by J. Richard Udry, Peter S. Bearman, and Kathleen Mullan Harris at the University of North Carolina at Chapel Hill. All funders can be found here: <https://addhealth.cpc.unc.edu/about/>.

**Adolescent Brain and Cognitive Development Study (ABCD US)**

The Adolescent Brain and Cognitive Development (ABCD) study is a longitudinal North American cohort of 11,876 individuals recruited between the ages of 9 and 10 years at baseline starting in 2015 across 21 different North American study sites (Volkow et al., 2018). Self-reported race and ethnicity in the ABCD cohort were as follows: Asian, Black, Hispanic, White and Other (alternative survey option).

The study was approved by the National Institute of Mental Health Data Archive, United States (NIMH). Written consent was obtained from all participants. Data was accessed through the NDA database (<https://nda.nih.gov/abcd/>); Federal-Wide Assurance: FWA00018101).

Data used in the preparation of this article were obtained from the Adolescent Brain Cognitive Development (ABCD) Study (https://abcdstudy.org), held in the NIMH Data Archive (NDA). This is a multisite, longitudinal study designed to recruit more than 10,000 children age 9-10 and follow them over 10 years into early adulthood. The ABCD Study® is supported by the National Institutes of Health and additional federal partners under award numbers U01DA041048, U01DA050989, U01DA051016, U01DA041022, U01DA051018, U01DA051037, U01DA050987, U01DA041174, U01DA041106, U01DA041117, U01DA041028, U01DA041134, U01DA050988, U01DA051039, U01DA041156, U01DA041025, U01DA041120, U01DA051038, U01DA041148, U01DA041093, U01DA041089, U24DA041123, U24DA041147. A full list of supporters is available at https://abcdstudy.org/federal-partners.html. A listing of participating sites and a complete listing of the study investigators can be found at https://abcdstudy.org/consortium_members/. ABCD consortium investigators designed and implemented the study and/or provided data but did not necessarily participate in the analysis or writing of this report. This manuscript reflects the views of the authors and may not reflect the opinions or views of the NIH or ABCD consortium investigators. The ABCD data repository grows and changes over time. The ABCD data used in this report came from <https://dx.doi.org/10.15154/8873-zj65>. DOIs can be found at <https://nda.nih.gov/abcd/>. Genetic data: GWAS summary statistics were downloaded from the PGC (<https://pgc.unc.edu/for-researchers/download-results/>) and iPSYCH (<https://ipsych.dk/en/research/downloads>) websites.

**Amsterdam Born Children and their Development (ABCD NL)**

The Amsterdam Born Children and their Development cohort is a multi-ethnic birth study, which follows the health, growth and development of approximately 8,000 children born in Amsterdam between January 2003 and March 2004 (van Eijsden et al., 2011). The study is conducted by Amsterdam University Medical Center in collaboration with the Public Health Service of Amsterdam and has been implemented in cooperation with hospitals and midwives, Youth Health Care (YHC) centres, primary schools and the University of Tilburg. The genetic data for this cohort were derived from the ABCD-GE study (ABCD-Genetic Enrichment), which was a substudy with 1192 ethnic Dutch children. During Phase 4 and 5 of the ABCD study information of the participants was collected at 10-13 years of age and 15-17 years, respectively. You can find more information on the [ABCD study website](https://www.amc.nl/web/abcd-studie-2/abcd-studie/achtergrondinformatie-abcd-studie.htm).

Approval of the study was obtained from the Central Committee on Research Involving Human Subjects in The Netherlands, the medical ethics review committees of the participating hospitals and the Registration Committee of the Municipality of Amsterdam (van Eijsden et al., 2011). All ABCD participants gave written informed consent for data collection of the phenotypes. Regarding the DNA collection and analysis, an opt-out procedure was used (METC approval 2002_039#B2013531) (Zafarmand et al., 2020).

The authors thank the participating mothers, fathers, their children, and all others who contributed to the ABCD-study: obstetric care providers, primary schools, students, and youth healthcare centres in Amsterdam, The Netherlands.

**Avon Longitudinal Study of Parents and Children (ALSPAC)**

The Avon Longitudinal Study of Parents and Children (ALSPAC) is a UK-based cohort of 15,645 children born between 1991 and 1992 (Boyd et al., 2013; Fraser et al., 2013; Northstone et al., 2019). Self-reported race and ethnicity in the ALSPAC cohort was overwhelmingly European White. Pregnant women resident in Avon, UK with expected dates of delivery between 1st April 1991 and 31st December 1992 were invited to take part in the study. The initial number of pregnancies enrolled was 14,541 with 13,988 children who were alive at 1 year of age. The total sample size for analyses using any data collected after the age of seven is therefore 15,454 pregnancies, resulting in 15,658 foetuses. Of these 14,901 children were alive at 1 year of age.

Ethical approval for the study was obtained from the ALSPAC Ethics and Law Committee and the Local Research Ethics Committees. Informed consent for the use of all data collected was obtained from participants following the recommendations of the ALSPAC Ethics and Law Committee at the time. Participants can contact the study team at any time to retrospectively withdraw consent for their data to be used. Study participation is voluntary and during all data collection sweeps, information was provided on the intended use of data. Consent for biological samples has been collected in accordance with the Human Tissue Act (2004).

Please note that the study website contains details of all the data that is available through a fully searchable data dictionary and variable search tool: <https://www.bristol.ac.uk/alspac/researchers/our-data/>. Permission to use the ALSPAC data is obtained through a proposal system managed by the ALSPAC executive. Part of this project used data collected by RedCap (Harris et al., 2009).

We are extremely grateful to all the families who took part in this study, the midwives for their help in recruiting them, and the whole ALSPAC team, which includes interviewers, computer and laboratory technicians, clerical workers, research scientists, volunteers, managers, receptionists and nurses.

The UK Medical Research Council and Wellcome (Grant ref: MR/Z505924/1) and the University of Bristol provide core support for ALSPAC. This publication is the work of the authors PZG and ASFKand will serve as guarantors for the contents of this paper. Genomewide genotyping data was generated by Sample Logistics and Genotyping Facilities at Wellcome Sanger Institute and LabCorp (Laboratory Corporation of America) using support from 23andMe.

**Australian Genetics of Depression Study (AGDS)**

Details about the Australian Genetics of Depression Study (AGDS) are published elsewhere (Byrne et al., 2020). Briefly, over 22,000 participants (approximately 17,000 genotyped) were recruited via Australian government prescription records or through a media campaign. Participants completed online questionnaires, including a core module assessing Major Depressive Disorder (MDD) diagnosis. Here, we used the Composite International Diagnostic Interview short form (CIDI-SF) diagnostic questionnaire (Kessler et al., 1998) to assess MDD DSM-5 criteria. MDD cases were defined as participants in AGDS who met DSM-5 criteria for MDD at some point within their lifetime.

We used the QSkin Sun and Health Study (QSkin) as a control cohort; a population-based cohort from Queensland, Australia that was invited to participate via a random draw from the electoral roll (Olsen et al., 2012). Participants completed a lifestyle questionnaire, including a checklist about previous diagnosis, experience or treatment for a range of conditions. MDD controls were defined as QSkin participants who did not report having been diagnosed, experienced or treated for depression nor experienced postnatal or antenatal depression. All protocols and questionnaires for both the AGDS and QSkin cohorts were approved by the QIMR Berghofer Medical Research Institute Human Research Ethics Committee (P1039, P2034, P2118).

**Brazilian High Risk Cohort Study (BHRC)**

The Brazilian High Risk Study for Mental Conditions (BHRCS), conducted at the Universidade Federal do Rio Grande do Sul, Universidade Federal de São Paulo, and Faculdade de Medicina da Universidade de São Paulo, used a two-stage design. First, we assessed childhood symptoms and family history of psychiatric disorders in a screening interview, collecting information from 9,937 index children at 57 schools in the cities of São Paulo and Porto Alegre, as well as from 45,394 family members. In the second stage, a random subsample (intended to be representative of the community, n = 957) and a high-risk subsample (children at increased risk for mental disorders, based on family risk and childhood symptoms, n = 1,554) were selected for further evaluation.

We evaluated those 2,512 subjects using an extensive protocol, involving one 2-hour home evaluation with the parents, two 1-hour evaluations of the child by a psychologist, and two 1-hour evaluations of the child by a speech pathologist. During those evaluations, saliva samples for genetic studies were collected from the subjects and their biological parents. Additionally, 750 children were invited to take part in a neuroimaging study and to provide blood samples for the assessment of peripheral blood biomarkers. These children have collectively been designated the enriched imaging cohort. We have a 3-year follow-up with 80% retention and a 6-year follow-up with 75% retention. For a full description of the cohort, see (10.1002/mpr.1459).

The study was approved by the ethics committee of the Hospital de Clínicas de Porto Alegre, University of São Paulo and Universidade Federal de São Paulo.

This study was supported by the National Institute of Developmental Psychiatry for Children and Adolescent (INPD) with grants from: Fundação de Amparo à Pesquisa do Estado de São Paulo (FAPESP; grant numbers: 2008/57896-8; 2014/50917-0; 2013/08531-5; 2020/06172-1; 2021/12901-9; 2021/05332-8; 2023/00437-1; 2023/05560-6); Conselho Nacional de Desenvolvimento Científico e Tecnológico (CNPq; grant numbers 573974/2008-0 and 465550/2014-2); European Research Council (ERC: 337673 and 101057390), UK Medical Research Council (MRC: MR/R022763/1); Ministério da Saúde (Decit/SECTICS/MS Grant number: 888379/2019 - Portaria Nº 1.949, 04/08/2020); Banco Industrial do Brasil S/A; CISM grant; Coordenação de Aperfeiçoamento de Pessoal de Nível Superior (CAPES): code #001. Collaboration between the BHRC and other cohorts has been funded by the National Institutes of Health (NIMH grant number: R01MH120482-01). Involvement of NIMH Intramural investigators has been funded by NIMH-Intramural Research Program Project MH 002782.

**Brisbane Longitudinal Twin Study (BLTS)**

The Brisbane Longitudinal Twin Study (BLTS) began in 1992 and recruited twins and their non-twin siblings from primary and secondary schools in the greater Brisbane area via media appeals and by word of mouth (Gillespie et al., 2013). The study was led by researchers from the Queensland Institute of Medical Research (QIMR). Twins were ascertained along with family members as part of a study examining the development of melanocytic naevi at ages 12 and 14, and of cognition at age 16.

Between 2009 and 2016, the Nineteen Up Study (19Up: Mapping neurobiological changes across mental health stages) assessed a range of mental health and behavioural problems and associated risk factors as part of the BLTS (Couvy-Duchesne et al., 2018). This was funded by the NIH NIDA to explore the genetic and environmental pathways to cannabis use, abuse, and dependence on 1,000 BLTS twins and their siblings. These data were supplemented with an additional 1,100 twins, with funding from the Australian National Health and Medical Research Council (NHMRC), along with seed funding from an NHMRC Australia grant.

In BLTS, depression cases were ascertained using the psychological distress subscale with a cut off of >= 2 from the Somatic and Psychological HEalth REport (SPHERE) (Hickie et al., 2001) (also used to assess mental health on most of the BLTS) (Mitchell et al., 2019).

Written, informed consent was obtained from a parent or guardian, and ethics approval was obtained from the Human Research Ethics Committee at the QIMR.

**Child and Adolescent Twin Study in Sweden (CATSS)**

The Child and Adolescent Twin Study in Sweden (CATSS) is an ongoing longitudinal study investigating the impact of genetic and environmental factors on health and behavior in children and young adults (Anckarsäter et al., 2011; Zagai et al., 2019). Launched in 1992, it includes all twins born in Sweden since July 1, 1992. Since 2004, parents of twins have been interviewed about their children's health and social environment at ages 9 and 12 (CATSS-9/12). At ages 15 (CATSS-15) and 18 (CATSS-18), twins and parents also complete questionnaires assessing health, personality, and psychosocial development. This GWAS study uses data from CATSS-15 and CATSS-18.

Subjects are protected by the informed consent process, in which they are informed of what is being collected and repeatedly given the option to withdraw their consent and discontinue their participation. The study has ethical approval from the Karolinska Institute Ethical Review Board: CATSS-15 Dnr: 2009/1599-32/5, CATSS-15/DOGSS Dnr: 03-672 and 2010/1356/31/1, and CATSS-18 Dnr: 2010/1410/31/1.

The use of data for this GWAS analysis is approved by the executive board of the Swedish Twin Register and is covered under the general ethical permit (PI: Yi Lu) for conducting MDD research (Dnr 2023-03073-01) by the Swedish Ethical Review Authority.

This work was supported by the US NIMH R01 MH123724, the European Union’s Horizon 2020 research and innovation program under grant agreement No 964874, the Swedish Research Council (Vetenskapsrådet, awards 2021-02615), and the European Research Council (grant agreement ID: 101042183). We thank the Swedish Twin Registry for providing access to the data used in this analysis.

**Gene-Environment Development Initiative (GEDI): Great Smoky Mountains Study (GSMS) & Virginia Twin-Family Study of Adolescent Behavioural Development (VTSABD)**

This cohort included samples from the Great Smoky Mountains Study (GSMS), the Virginia Twin Study of Adolescent Behavioral Development (VTSABD), and the Caring for Children in the Community Study. Additional details about these studies and the samples included in this analysis are available in (Costello et al., 2013).

Participants and parents were interviewed with the depressive disorders module of the Child and Adolescent Psychiatric Assessment interview. Responses from this interview were then used to generate DSM depressive disorder diagnoses from the symptoms assessed by computer algorithms. If either parent or child reported a symptom as present in the past three months, it was counted as present. A three-month “primary period” was selected because longer recall periods are associated with forgetting and recall bias.

The consents and protocols for GSMS were approved by the Duke Medical Institutional Review Board. Ethical approval for the VTSABD was provided by the institutional review board of Virginia Commonwealth University, and appropriate informed consent was obtained from all persons participating in this research.

HHM Maes was supported by NIH R01 DA054313. RE Peterson and C Chatzinakos were supported by NIH R01MH125938 and by The Brain & Behavior Research Foundation NARSAD grant 28632 P&S Fund (RE Peterson). The content is solely the responsibility of the authors and does not necessarily represent the official views of the National Institutes of Health.

**Generation R (GenR)**

Generation R is a prospective population-based study from fetal life until young adulthood, to understand the early environmental and genetic causes of normal and abnormal growth from fetal life until young adulthood. The nature of this study is multiethnic. The Genome data was gathered at two time points with different arrays and independent participants and are referred to here as GenerationR3 and GenerationR4 (Ghatan et al., 2024; Medina-Gomez et al., 2015). Website: <https://generationr.nl/> The generation and management of GWAS genotype data for the Generation R Study was done at the Human Genomics Facility, HuGe-F, housed within the Laboratory for Population Genomics of the Department of Internal Medicine at Erasmus MC. Genetic Laboratory of the Department of Internal Medicine, Erasmus MC, The Netherlands. We thank Zahra Alawi, Marijn Verkerk, Dr. Katerina Trajanoska, Costanza Vallerga, Samuel Gathan, Dr. Carolina Medina-Gomez, Dr. Linda Broer and Jard de Vries for their help in creating, managing and QC the GWAS database.

The Generation R Study is conducted by Erasmus MC in close collaboration with the Erasmus University Rotterdam, Faculty of Social Sciences, the Municipal Health Service Rotterdam area, the Rotterdam Homecare Foundation, Rotterdam and the Stichting Trombosedienst & Artsenlaboratorium Rijnmond (STAR-MDC), Rotterdam. The general design of the Generation R Study is made possible by financial support from the Erasmus MC, University Medical Center, Rotterdam, the Netherlands, the Organization for Health Research and Development (ZonMw) and the Ministry of Health, Welfare and Sport. We gratefully acknowledge the contribution of children and their parents, general practitioners, hospitals, midwives and pharmacies in Rotterdam.

The general design, all research aims and the specific measurements in the Generation R Study have been approved by the Medical Ethical Committee of Erasmus MC, University Medical Center Rotterdam. New measurements are only introduced into the study after approval of the Medical Ethical Committee. Participants need to give written informed consent for each phase of the study (fetal, preschool, childhood and adolescence period). From the age of 12 years onwards, children must sign their own consent form, in accordance with Dutch Law. At the start of each phase, children and their parents receive written and oral information about the study. Even with consent, when the child or the parents are not willing to participate actively, specific measurements are skipped or no measurements at all are performed.

This work was supported by the Erasmus MC Sophia Foundation (“Stiching Vrienden van het Sophia,” Grant WAR24-30, AN, MS), the European Research Council (TEMPO; grant agreement No 101039672; AN) and the European Union’s HorizonEurope Research and Innovation Programme (FAMILY, grant agreement No 101057529;AN).

**Generation Scotland (GenScot)**

Generation Scotland is a cohort study of 7,000 families recruited from the general population of Scotland (Smith et al., 2013). All clinical participants were screened for a history of emotional and psychiatric disorders using the structured clinical interview for DSM-IV disorders (SCID) (Fernandez-Pujals et al., 2015; Smith et al., 2013); 21.7% screened positive and were invited to continue the interview that focused on mood disorders; of these, 88% completed the interview (19.0% of participants). Adolescent-onset depression cases were defined as participants in Generation Scotland who met DSM-5 criteria for MDD between the ages of 10-19 years using the SCID-5. MDD controls were defined as participants who did not meet DSM-5 criteria for MDD at any point within their lifetime. We removed participants overlapping with UK Biobank.

Tayside Committee on Medical Research Ethics A (reference: 05/S1401/89), Research Tissue Bank by the East of Scotland Research Ethics Service (reference: 20/ES/0021)

Generation Scotland received core support from the Chief Scientist Office of the Scottish Government Health Directorates (CZD/16/6) and the Scottish Funding Council (HR03006) and is currently supported by the Wellcome Trust (216767/Z/19/Z). Genotyping of the Generation Scotland samples was carried out by the Genetics Core Laboratory at the Edinburgh Clinical Research Facility, University of Edinburgh, Scotland and was funded by the Medical Research Council UK and the Wellcome Trust (Wellcome Trust Strategic Award “STratifying Resilience and Depression Longitudinally” (STRADL) Reference 104036/Z/14/Z).

**Genetic Links to Anxiety and Depression (GLAD)**

The Genetic Links to Anxiety and Depression (GLAD) Study (www.gladstudy.org.uk) recruits individuals with depression or anxiety into the NIHR Mental Health BioResource (Davies et al., 2019). Participants invited to join the study (via media campaigns) provide demographic, environmental and genetic data, and consent for medical record linkage and recontact. Currently, the GLAD study has over 66,000 consented participants, with more than 50,000 having completed the online survey, and over 36,000 saliva samples collected. The study sample is severe, highly comorbid, with chronic psychopathology. During the COVID-19 pandemic, the GLAD Study research team recontacted GLAD participants and healthy volunteers from other NIHR BioResource studies, with the aim of estimating the impact of the pandemic on mental and neurological health (COVID-19 Psychiatric and Neurology Genetics Study (COPING) study). This resulted in recruiting over 20,000 participants with common mental disorders and over 11,000 healthy volunteers, two-thirds of whom have been genotyped. All participants provided informed consent for the GLAD and/or COPING studies and the NIHR BioResource (<https://bioresource.nihr.ac.uk/>).

The GLAD Study was approved by the London https://gladstudy.org.uk/ - Fulham Research Ethics Committee on 21st August 2018 (REC reference: 18/LO/1218) following a full review by the committee. The NIHR BioResource has been approved as a Research Tissue Bank by the East of England - Cambridge Central Committee (REC reference: 17/EE/0025). Prior to submission for ethical approval, this research was reviewed by a team with experience of mental health problems and their carers who have been specially trained to advise on research proposals and documentation through the Feasibility and Acceptability Support Team for Researchers (FAST-R): a free, confidential service in England provided by the NIHR Maudsley Biomedical Research Centre via King's College London and South London and Maudsley NHS Foundation Trust.

We thank NIHR BioResource volunteers for their participation, and gratefully acknowledge NIHR BioResource centres, NHS Trusts and staff for their contribution. We thank the National Institute for Health and Care Research, NHS Blood and Transplant, and Health Data Research UK as part of the Digital Innovation Hub Programme. The views expressed are those of the author(s) and not necessarily those of the NHS, the NIHR or the Department of Health and Social Care.

We gratefully acknowledge the participation of the NIHR BioResource Centre Maudsley, Biomedical Research Centre at South London and Maudsley NHS Foundation Trust and King’s College London volunteers and thank the staff for their help with volunteer recruitment. We thank the NIHR Biomedical Research Centre at South London and the Maudsley NHS Foundation Trust and King’s College London for funding. This study represents independent research supported by the NIHR Biomedical Research Centre BioResource at South London and Maudsley NHS Foundation Trust and King's College London. We gratefully acknowledge capital equipment funding from the Maudsley Charity (Grant Ref. 980) and Guy’s and St Thomas’s Charity (Grant Ref. STR130505).

**Growing Up in Singapore Towards healthy Outcomes (GUSTO)**

GUSTO (Growing Up in Singapore Towards healthy Outcomes) is a prospective mother-offspring cohort study conducted in Singapore between June 2009 and September 2010 at KK Women’s and Children’s Hospital (KKH) and National University Hospital (NUH) (Soh et al., 2014). The study recruited 1247 pregnant women with singleton pregnancies who self-identified as Chinese, Malay, or Indian at 11-14 weeks of gestation. Inclusion criteria included the age range of 18-50, the intention to reside in Singapore within 5 years after the recruitment, the intention to deliver in KKH and NUH, and a willingness to contribute antenatal and cord blood. Women receiving chemotherapy, psychotropic drugs or those with type I diabetes mellitus were excluded.

This study was approved by the National Health Care Group Domain Specific Review Board (reference D/2009/021 & B/2014/00411) and the SingHealth Centralized Institutional Review Board (reference 2018/2767 & 2019/2406). This study has been performed in accordance with the relevant guidelines and written information consent was obtained from all participants upon recruitment.

GUSTO study is supported by the National Research Foundation (NRF) under the Open Fund-Large Collaborative Grant (OF-LCG; MOH-000504) administered by the Singapore Ministry of Health’s National Medical Research Council (NMRC) and the Agency for Science, Technology and Research (A*STAR). In RIE2025, GUSTO is supported by funding from the NRF’s Human Health and Potential (HHP) Domain, under the Human Potential Programme. Gusto thanks all study group members.

We acknowledge the GUSTO study group members: Airu Chia, Andrea Cremaschi, Anna Magdalena Fogel, Anne Eng Neo Goh, Anne Rifkin-Graboi, Anqi Qiu, Arijit Biswas, Bee Wah Lee, Birit Froukje Philipp Broekman, Candida Vaz, Chai Kiat Chng, Chan Shi Yu, Choon Looi Bong, Daniel Yam Thiam Goh, Dawn Xin Ping Koh, Dennis Wang, Desiree Y. Phua, E Shyong Tai, Elaine Kwang Hsia Tham, Elaine Phaik Ling Quah, Elizabeth Huiwen Tham, Evelyn Chung Ning Law, Evelyn Keet Wai Lau, Evelyn Xiu Ling Loo, Fabian Kok Peng Yap, Falk Müller-Riemenschneider, Franzolini Beatrice, George Seow Heong Yeo, Gerard Chung Siew Keong, Hannah Ee Juen Yong, Helen Yu Chen, Hong Pan, Huang Jian, Huang Pei, Hugo P S van Bever, Hui Min Tan, Iliana Magiati, Inez Bik Yun Wong, Ives Lim Yubin, Ivy Yee-Man Lau, Jacqueline Chin Siew Roong, Jadegoud Yaligar, Jerry Kok Yen Chan, Jia Xu, Johan Gunnar Eriksson, Jonathan Tze Liang Choo, Jonathan Y. Bernard, Jonathan Yinhao Huang, Joshua J. Gooley, Jun Shi Lai, Karen Mei Ling Tan, Keith M. Godfrey, Keri McCrickerd, Kok Hian Tan, Kothandaraman Narasimhan, Krishnamoorthy Naiduvaje, Kuan Jin Lee, Li Chen, Lieng Hsi Ling, Lin Lin Su, Ling-Wei Chen, Lourdes Mary Daniel, Lynette Pei-Chi Shek, Maria De Iorio, Marielle V. Fortier, Mary Foong-Fong Chong, Mary Wlodek, Mei Chien Chua, Melvin Khee-Shing Leow, Michael J. Meaney, Michelle Zhi Ling Kee, Min Gong, Mya Thway Tint, Navin Michael, Neerja Karnani, Ngee Lek, Noor Hidayatul Aini Bte Suaini, Oon Hoe Teoh, Peter David Gluckman, Priti Mishra, Queenie Ling Jun Li, Sambasivam Sendhil Velan, Seang Mei Saw, See Ling Loy, Seng Bin Ang, Shang Chee Chong, Shiao-Yng Chan, Shirong Cai, Shu-E Soh, Stephen Chin-Ying Hsu, Suresh Anand Sadananthan, Swee Chye Quek, Tan Ai Peng, Varsha Gupta, Victor Samuel Rajadurai, Wee Meng Han, Wei Wei Pang, Yap Seng Chong, Yin Bun Cheung, Yiong Huak Chan, Yung Seng Lee, Zhang Han.

**Impact of Neurodevelopmental Disorders and School Performance (INSchool) (Waves 1-3)**

The Impact of Neurodevelopmental disorders and School performance: genes and environment (INSchool) is part of ongoing research aimed to identify children and adolescents’ mental health problems in a school setting. Schools from different counties in Catalonia were contacted and invited to participate after explaining the study to the school staff. Participants include children and adolescents aged 5 to 18 years old (Bosch et al., 2022).

The project was authorised by the Ministry of Health and the Ministry of Education (Generalitat de Catalunya, Spain), with the approval from the Ethics Committee of the Vall d’Hebron Hospital Universitari, in Barcelona. Families were informed and written consent was obtained. Participants who were at least aged 11 also gave permission.

This work was funded by the Agència de Gestió d’Ajuts Universitaris i de Recerca (AGAUR, 2017SGR-1461, 2021SGR-00840), the Instituto de Salud Carlos III (PI19/01224, PI20/00041, PI22/00464 and PI23/00404, PI23/00026, PI24/00195), the Network Center for Biomedical Research (CIBER); the European Regional Development Fund (ERDF); the ECNP Network ‘ADHD across the Lifespan’; “La Marató de TV3” (202228-30 and 202228-31), Fundació ‘la Caixa’, Diputació de Barcelona, “Pla Estratègic de Recerca i Innovació en Salut” (PERISSLT006/17/285); “Fundació Privada d'Investigació Sant Pau” (FISP); and Ministry of Health of Generalitat de Catalunya. SA acknowledges her Miguel Servet contract (CP22/00026) awarded by the Instituto de Salud Carlos III and co-funded by the European Union Found: Fondo Social Europeo Plus, FSE +. The genotyping service was carried out at the Genotyping Unit-CEGEN in the Spanish National Cancer Research Centre (CNIO), supported by Instituto de Salud Carlos III (ISCIII), Ministerio de Ciencia e Innovación. CEGEN is part of the initiative IMPaCTGENóMICA (IMP/00009) cofunded by ISCIII and the European Regional Development Fund (ERDF).

The authors are grateful to families, students, and staff of the public primary schools (i.e., Joan Maragall, Maria Borés, Marquès de la Pobla, Martinet, Pins del Vallès, Puiggraciós, Sant Jordi, Ramon Llull, Rivo Rubeo, Tagamanent and Teresa Berguedà), public secondary schools (i.e., Angeleta Ferrer i Sensat, Antoni Pous i Argila, Cal Gravat, Duc de Montblanc, Institut del Ter, Jaume Callís, Lacetània, Lluís de Peguera, Molí de la Vila, Montsuar, Pius Font i Quer, Vallbona d’Anoia, and Vil·la Romana), and private schools (i.e., Airina, L'Ave Maria, Casals – Gràcia, Episcopal Lleida, La Farga, FEDAC Manresa, FEDAC Vic, Garbí Pere Vergés Esplugues, Institució Igualada, Joviat, Oms i de Prat, Pies Mataró, Pureza de María, Regina Carmeli, Sagrats Cors Centelles, La Salle Manlleu, La Salle Manresa, Sant Miquel dels Sants, Thau Barcelona and Vedruna Escorial Vic) who kindly contribute in this research.

**Janssen**

MDD cases must meet criteria of the Diagnostic and Statistical Manual of Mental Diseases, 4th edition (DSM-IV), for Major Depressive Disorder with history of resistance to therapy with antidepressant medication. Genetic sampling was optional in this study (NCT00044681). Control data were obtained from the Alzheimer’s Disease Neuroimaging Initiative (ADNI) database (adni.loni.usc.edu).

All patients who provided genetic samples gave written informed consent to the genetic testing. The adolescent-onset depression GWAS data were collected in a multi-site clinical trial sponsored by Janssen (J&J) in 2002. Each site received IRB/Ethical Committee approval for conducting the clinical study (NCT00044681) prior to participant screening, and all participants gave written informed consent.
There was no single central IRB.

We are grateful to the study volunteers for participating in the research studies and to the clinicians and support staff for enabling patient recruitment and blood sample collection. Informed consent was obtained from all participants or their parents or guardians. We thank the staff in the former Neuroscience Biomarkers of Janssen Research & Development for laboratory and operational (including but not limited to sample banking, processing, plating, and clinical data/sample de-identification) support, and the staff at Illumina for genotyping Janssen DNA samples.

Control data were obtained from the Alzheimer’s Disease Neuroimaging Initiative (ADNI) database (adni.loni.usc.edu). As such, the investigators within the ADNI contributed to the design and implementation of ADNI and/or provided data but did not participate in analysis or writing of this report. A complete listing of ADNI investigators can be found at:

<http://adni.loni.usc.edu/wp-content/uploads/how_to_apply/ADNI_Acknowledgement_List.pdf>

**Lifelines (GSA and Affymetrix)**

Lifelines is a multi-disciplinary prospective population-based cohort study examining in a unique three-generation design the health and health-related behaviours of 167,729 persons living in the north of the Netherlands (Sijtsma et al., 2022). It employs a broad range of investigative procedures in assessing the biomedical, socio-demographic, behavioural, physical and psychological factors which contribute to the health and disease of the general population, with a special focus on multi-morbidity and complex genetics.

Participants were recruited between 2006-2013 via their GP (49%), participating family members (38%), and self-registration on the Lifelines website (13%) (Scholtens et al., 2015). Exclusion criteria for GP recruitment were: insufficient knowledge of Dutch language, severe psychiatric or physical illness, limited life expectancy (<5 years) (Scholtens et al., 2015). Baseline data included approximately: 15,000 children (0-17 years), 140,000 adults (18-65 years), 12,000 elderly individuals (65+ years). Following baseline, participants were invited to attend study visits every 5 years (2nd follow-up visit finished end of 2023. In between assessments, participants completed follow-up questionnaires every 1.5-2.5 years (Scholtens et al., 2015).

The general Lifelines protocol has been approved by the UMCG Medical ethical committee under number 2007/152.

The Lifelines initiative has been made possible by subsidy from the Dutch Ministry of Health, Welfare and Sport, the Dutch Ministry of Economic Affairs, the University Medical Center Groningen (UMCG), Groningen University and the Provinces in the North of the Netherlands (Drenthe, Friesland, Groningen). The generation and management of GWAS genotype data for the Lifelines Cohort Study is supported by the UMCG Genetics Lifelines Initiative (UGLI). UGLI is partly supported by a Spinoza Grant from NWO, awarded to Cisca Wijmenga. The authors wish to acknowledge the services of the Lifelines Cohort Study, the contributing research centres delivering data to Lifelines, and all the study participants.

**Mater-University of Queensland Study of Pregnancy (MUSP)**

MUSP study is of 7223 women recruited early in pregnancy over the period 1981-1983 and the live singleton children to whom they subsequently gave birth (Najman et al., 2015). There was follow-up of the mothers at 5, 14, 21 and 27 years after recruitment. Children were also followed up in the mothers’ questionnaire and independently at 21 and 30 years of age. The CBCL and YSR were administered to children at 5, 14 and 21 years of age. The CIDI was administered to mothers at 27 years after recruitment and the children at 21 and 30 years after recruitment. The cohort comprises both mothers (to 27 years after the birth) and children (to 30 years of age).

This research was approved by the Human Research Ethics Committee of the University of Queensland, in accordance with the ethical standards of the institution.

This study was funded by grants received from the National Health and Medical Research Council (NHMRC) and Australian Research Council (ARC). Thanks also to Shelby Marrington (Project Manager) and Greg Shuttlewood (Data Manager) who have supervised the day-to-day management of the study. We also extend our thanks to the mothers and children who have continued to participate in the study. The authors thank the MUSP study participants and study team. The authors thank the National Health and Medical Research Council (NHMRC).

**Millenium Cohort Study (MCS)**

The Millennium Cohort Study (MCS) follows the lives of young people born across England, Scotland, Wales and Northern Ireland in 2000-02 (Joshi & Fitzsimons, 2016). The study began with an original sample of 18,818 cohort members. Cohort members were genotyped at age 14. Link to study: <https://cls.ucl.ac.uk/cls-studies/millennium-cohort-study/>

MCS received ethical approval from the National Health Service Research Ethics Committee at each sweep of data collection. Informed consent was provided by parents/caregivers, and children agreed to participate. From age 16, cohort members provided verbal informed consent to participate in the study. Further details of the ethical approval and consent process can be found here: <https://cls.ucl.ac.uk/wp-content/uploads/2017/07/MCS-Ethical-Approval-and-Consent-2019.pdf>.

The authors are grateful to the Centre for Longitudinal Studies (CLS), UCL Social Research Institute, for the use of MCS data and to the UK Data Service for making them available. However, neither CLS nor the UK Data Service bear any responsibility for the analysis or interpretation of these data.

**Norwegian Mother, Father and Child Cohort Survey (MoBa)**

The Norwegian Mother, Father and Child Cohort Study (MoBa) is a population-based pregnancy cohort study conducted by the Norwegian Institute of Public Health (Magnus et al., 2016). Participants were recruited from all over Norway from 1999-2008. The women consented to participation in 41% of the pregnancies. The cohort includes approximately 114.500 children, 95.200 mothers and 75.200 fathers. The current study is based on version 12 of the quality-assured data files released for research in Jan 2019.

The establishment of MoBa and initial data collection was based on a license from the Norwegian Data Protection Agency and approval from The Regional Committees for Medical and Health Research Ethics. The MoBa cohort is currently regulated by the Norwegian Health Registry Act. The current study was approved by The Regional Committees for Medical and Health Research Ethics (2016/1702).

The Norwegian Mother, Father and Child Cohort Study is supported by the Norwegian Ministry of Health and Care Services and the Ministry of Education and Research. We are grateful to all the participating families in Norway who take part in this on-going cohort study. This work was performed on the TSD (Tjeneste for Sensitive Data) facilities, owned by the University of Oslo, operated and developed by the TSD service group at the University of Oslo, IT-Department (USIT;). The analyses were performed on resources provided by Sigma2 - the National Infrastructure for High Performance Computing and Data Storage in Norway. We thank the Norwegian Institute of Public Health (NIPH) for generating high-quality genomic data. This research is part of the HARVEST collaboration, supported by the Research Council of Norway (#229624). We also thank the NORMENT Centre for providing genotype data, funded by the Research Council of Norway (#223273), South East Norway Health Authorities and Stiftelsen Kristian Gerhard Jebsen. We further thank the Center for Diabetes Research, the University of Bergen for providing genotype data and performing quality control and imputation of the data funded by the ERC AdG project SELECTionPREDISPOSED, Stiftelsen Kristian Gerhard Jebsen, Trond Mohn Foundation, the Research Council of Norway, the Novo Nordisk Foundation, the University of Bergen, and the Western Norway Health Authorities.

This work was supported by Helse Sør-Øst (#2020022; 2020024) and the Research Council of Norway (#336085).

**Psychiatric Genomics Consortium (PGC)**

Data from the PGC drawn from 23 cohorts in the Wave 1 and Wave 2 datasets of the Major Depressive Disorder Working Group (Major Depressive Disorder Working Group of the Psychiatric GWAS Consortium et al., 2013; Wray et al., 2018). Symptoms were assessed by trained interviewers using structured diagnostic instruments and DSM checklists. The analysis used data from 2 cohorts from the PGC MDD datasets that had age of onset data on cases. Data was drawn from PsyCoLaus (col3) and NESDA/NTR (nes1). The PGC has received major funding from the US National Institute of Mental Health (MH124873).

**Raine Study (Raine)**

The Raine Study is a prospective pregnancy cohort of 2900 mothers recruited between 1989–1991 (<https://www.rainestudy.org.au/>) (Newnham et al., 1993). Recruitment took place at Western Australia’s major perinatal centre, King Edward Memorial Hospital, and nearby private practices. Women who had sufficient English language skills, an expectation to deliver at King Edward Memorial Hospital, and an intention to reside in Western Australia to allow for future follow-up of their child were eligible for the study. The primary carers (Gen1) completed questionnaires regarding their respective study child, and the children (Gen2) had physical examinations at ages 1, 2, 3, 5, 8, 10, 14, 17, 18, 20, 22, 27 and 28.

Ethics approval for the original pregnancy cohort and subsequent follow-ups were granted by the Human Research Ethics Committee of King Edward Memorial Hospital, Princess Margaret Hospital, the University of Western Australia, and the Health Department of Western Australia. Parents, guardians and young adult participants provided written informed consent either before enrolment or at data collection at each follow-up.

The Raine Study was supported by the National Health and Medical Research Council of Australia [grant numbers 572613, 403981, 1059711], the Canadian Institutes of Health Research [grant number MOP-82893]. The authors would like to acknowledge the Raine Study participants and their families for their ongoing participants in the study and the Raine Study team for cohort co-ordination and data collection. The authors also thank the NHMRC and the Raine Study Medical Research Foundation for their long-term contribution to funding to the study over the last 30 years. The core management of the Raine Study is funded by The University of Western Australia, Curtin University, Telethon Kids Institute, Women and Infants Research Foundation, Edith Cowan University, Murdoch University, and The University of Notre Dame Australia. This work was supported by resources provided by the Pawsey Supercomputing Centre with funding from the Australian Government and Government of Western Australia.

**Tracking Adolescents’ Individual Lives Survey (TRAILS)**

TRAILS (Tracking Adolescents' Individual Lives Survey) is an ongoing, multidisciplinary study on the psychological, social and physical development of adolescents and young adults (Hartman et al., 2022; Oldehinkel et al., 2015). TRAILS consists of a population cohort (N = 2230) and a clinical cohort (N = 543), both of which were followed from about age 11 years onwards. Eight waves of assessment have been performed since and participants are now in their thirties.

Ethical Approval Ethical approval for TRAILS was obtained for each wave of data collection from the Dutch national ethics committee Central Committee on Research Involving Human Subjects (CCMO) or the local medical ethical review board (METc UMC Groningen).

Informed written consent was obtained from both adolescents (all waves) and their parents (first three waves) prior to each assessment wave.

TRAILS has been financially supported by various grants from the Netherlands Organization for Scientific Research NWO (Medical Research Council program grant GB-MW 940-38-011; ZonMW Brainpower grant 100-001-004; ZonMw Risk Behavior and Dependence grant 60-60600-97-118; ZonMw Culture and Health grant 261-98-710; Social Sciences Council medium-sized investment grants GB-MaGW 480-01-006 and GB-MaGW 480-07-001; Social Sciences Council project grants GB-MaGW 452-04-314 and GB-MaGW 452-06-004; ZonMw Longitudinal Cohort Research on Early Detection and Treatment in Mental Health Care grant 636340002; NWO large-sized investment grant 175.010.2003.005; NWO Longitudinal Survey and Panel Funding 481-08-013 and 481-11-001; NWO Vici 016.130.002, 453-16-007/2735, and Vi.C.191.021; NWO Gravitation 024.001.003), the Dutch Ministry of Justice (WODC), the European Science Foundation (EuroSTRESS project FP-006), the European Research Council (ERC-2017-STG-757364 and ERC-CoG-2015-681466), Biobanking and Biomolecular Resources Research Infrastructure BBMRI-NL (CP 32), the Gratama foundation, the Jan Dekker foundation, the participating universities, and Accare Centre for Child and Adolescent Psychiatry.

**Twins Early Development Study (TEDS)**

The Twins Early Development Study (TEDS) is a longitudinal twin study that recruited over 13,000 twin-pairs born between 1994 and 1996 in England and Wales through national birth records (Lockhart et al., 2023). More than 10,000 of these families are still engaged in the study. TEDS was and still is a representative sample of the population in England and Wales. Rich cognitive and emotional/behavioural data have been collected from the twins from infancy to emerging adulthood, with data collection at first contact and at ages 2, 3, 4, 7, 8, 9, 10, 12, 14, 16, 18, 21 and 26. Genotyped DNA data are available for 10,346 individuals (who are unrelated except for 3320 dizygotic co-twins). TEDS data have contributed to over 400 scientific papers involving more than 140 researchers in 50 research institutions.

Ethical approval for TEDS is provided by the Ethics Committee (reference: PNM/09/10–104). Written informed consent was obtained prior to each wave of data collection from parents and from twins themselves from age 16 onwards.

We gratefully acknowledge the ongoing contribution of the participants in the Twins Early Development Study (TEDS) and their families. TEDS is supported by the UK Medical Research Council (MR/V012878/1 and previously MR/M021475/1).

**UK Biobank (UKB)**

The UK Biobank (UKB) is a deeply phenotyped population health cohort recruited from general practitioners in the United Kingdom (Bycroft et al., 2018; Sudlow et al., 2015). The UKB cohort includes ~500,000 people from the UK aged 40-69 at recruitment, which took place across 22 assessment centres in the UK between 2006-2010. Genotype data is available for 488,377 participants, covering 805,426 variants. Website: <https://www.ukbiobank.ac.uk/>

Lifetime depression symptoms were assessed during online recontact and taken from the CIDI portion of the Mental Health Questionnaire (Davis et al., 2019) (UKB-MHQ, N=157,366). For the CIDI, low mood and anhedonia were used as gating symptoms, where participants had to endorse at least one to be asked about the other symptoms. The symptom was present if the question was endorsed as "Yes", absent if answered as "No", and missing otherwise.

UK Biobank received ethical approval from the Research Ethics Committee (reference 11/NW/0382). All participants gave informed consent for the collection of phenotypic, genetic and biomarker data. UK Biobank is supported by the [Wellcome Trust](http://www.wellcome.ac.uk/), [Medical Research Council](https://www.ukri.org/councils/mrc/), [Department of Health](http://www.dh.gov.uk/en/index.htm), [Scottish Government](https://www.gov.scot/), and the Northwest Regional Development Agency.

**Supplementary Figures 1-17**

**Supplementary Figure 1. European meta-analysis with chromosome X**

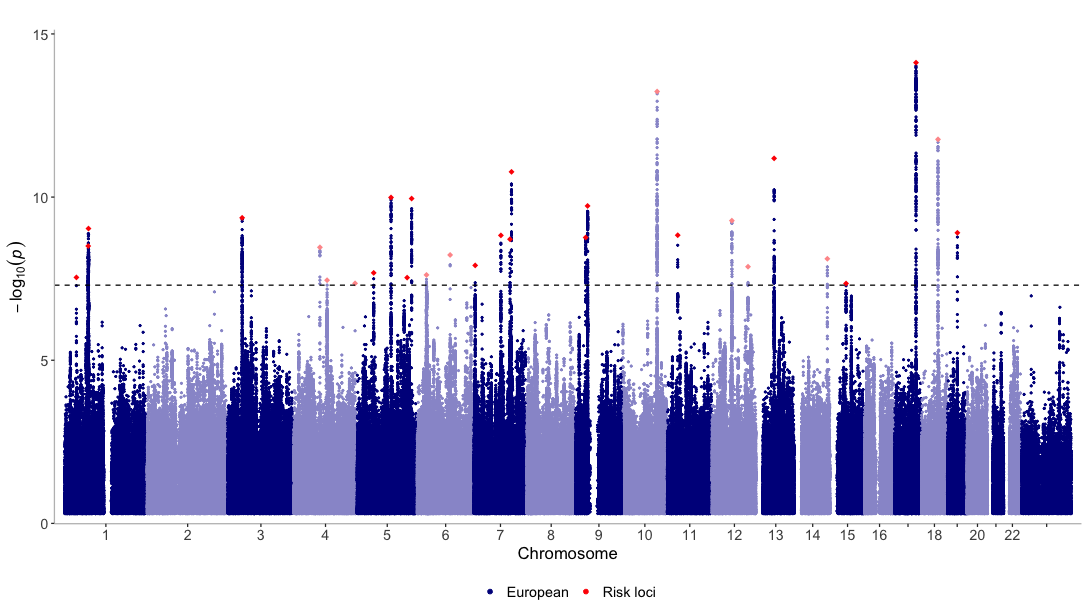

**Supplementary Figure 2. European prospective cohorts GWAS**

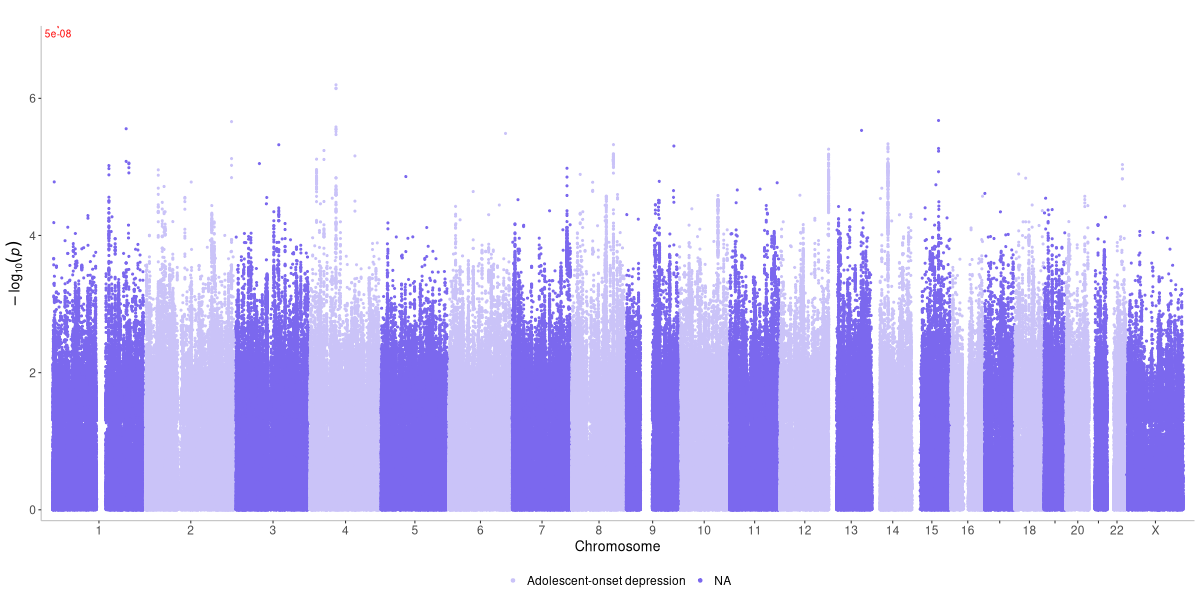

The meta-analysis of prospective samples included 20,432 cases and 57,349 controls and identified no genome-wide significant SNPs (SNP-*h^2^_liability_* = 10.9% (SE 1.0%), λ_GC_ = 1.052, polygenicity inflation = 78%). The meta-analysis resulted in 6002937 SNPs.

**Supplementary Figure 3. European retrospective cohorts GWAS**

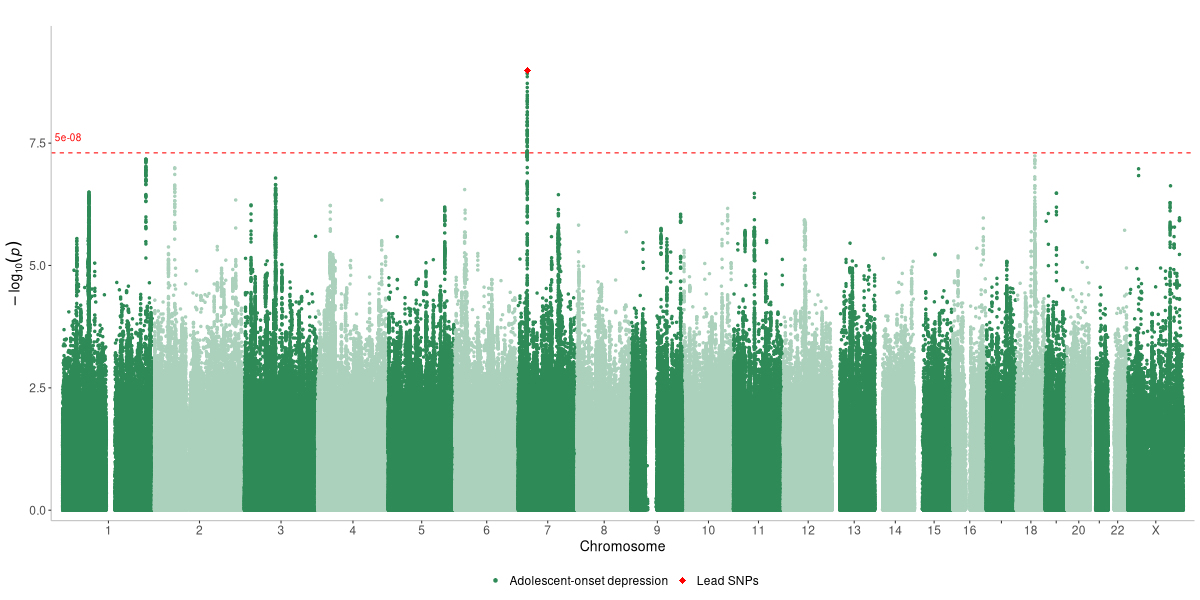

The retrospective sample included 31,902 cases and 120,060 controls and identified 1 genome-wide significant loci (rs2529089) on chromosome 7 (SNP-*h^2^_liability_* = 13.3% (SE 0.5%), λ_GC_ = 1.162, polygenicity inflation = 89%). The meta-analysis resulted in 7994741 SNPs

**Supplementary Figure 4. Nordic clinical early-onset GWAS**

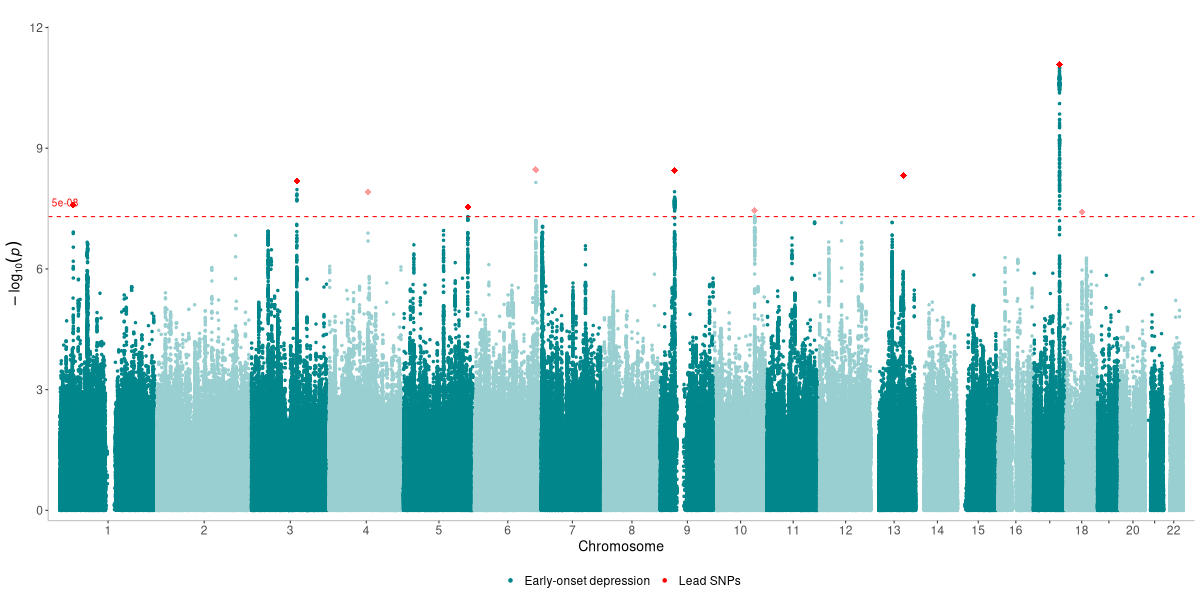

GWAS results for the full clinical early onset sample are reported in detail in Shorter et al (in press). We used a version of these summary statistics that omitted the MoBa cohort to prevent overlap. This sample included 45,826 cases and 99,141 controls (SNP-*h^2^_liability_* = 11.2% (SE = 0.7%), λ_GC_ = 1.181, polygenicity inflation = 71%).

**Supplementary Figure 5. African ancestry GWAS**
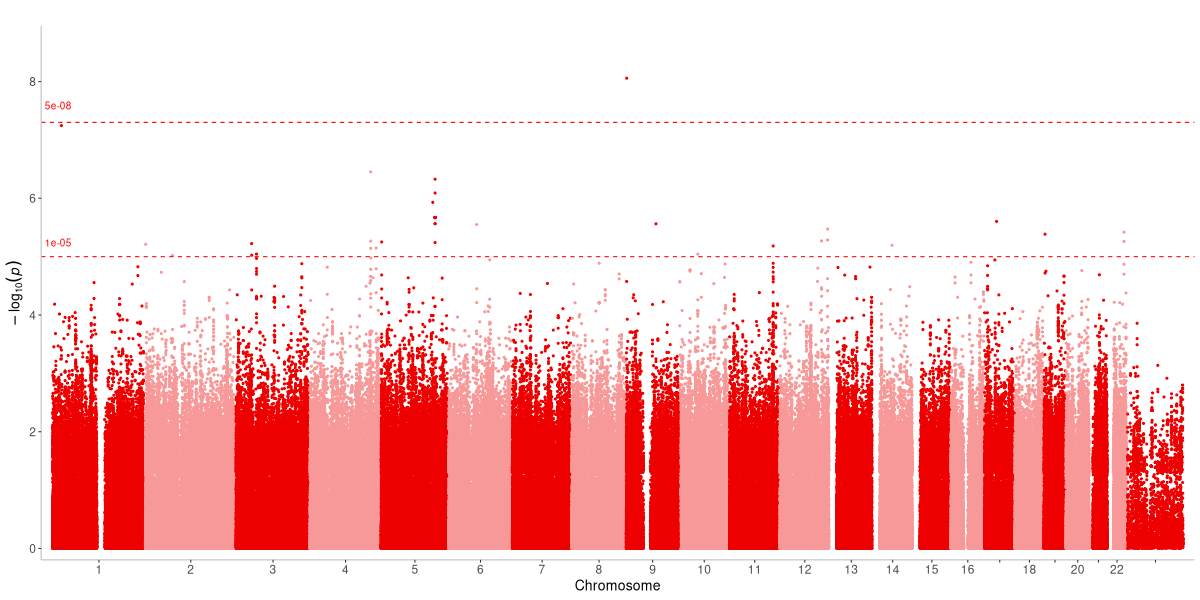

Independent African ancestry analyses included 1475 cases, and 3607 controls and Africans revealed 1 genome-wide significant loci (rs140713200) on chromosome 9 and 1 mapped gene (DMRT1, λ_GC_ = 0.999). The meta-analysis resulted in 4265384 SNPs.

**Supplementary Figure 6. American-admixed ancestry GWAS**

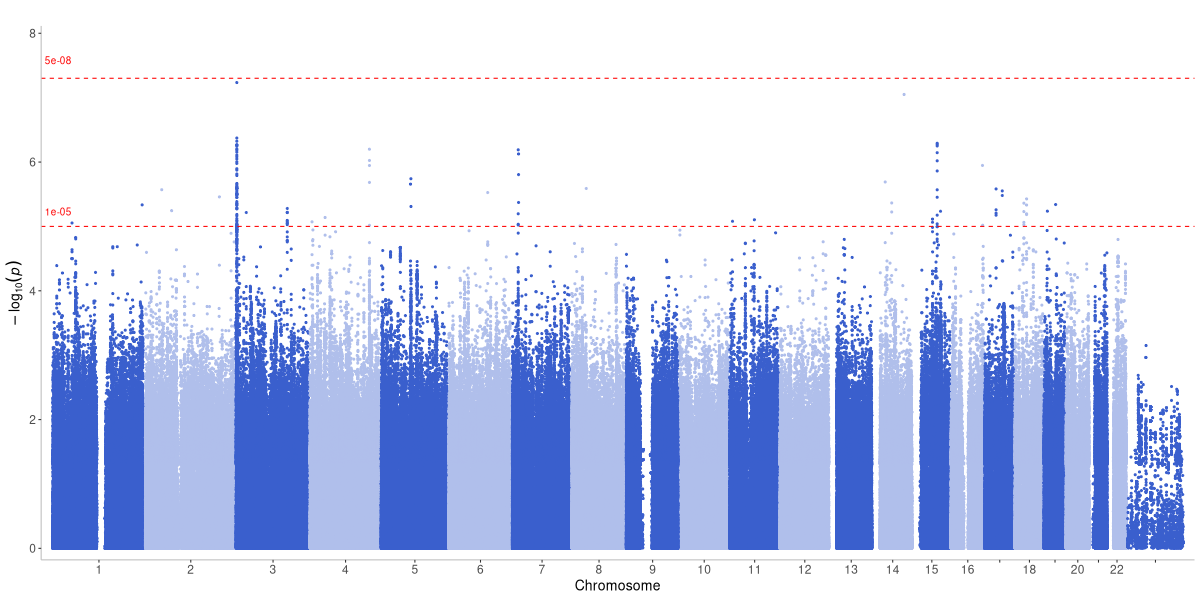

Independent American-admixed ancestry analyses included 2136 cases and 3701 controls and revealed no genome-wide significant loci (λ_GC_ = 1.048). The meta-analysis resulted in 7441952 SNPs.

**Supplementary Figure 7. South Asian ancestry GWAS**
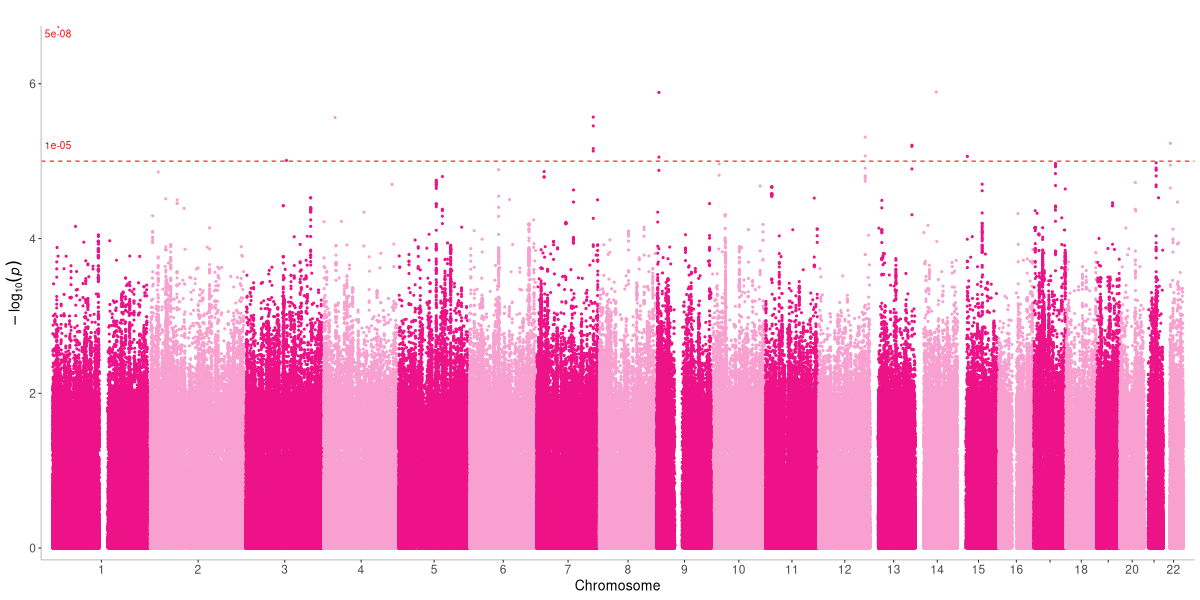

Independent South Asian ancestry analyses included 251 cases and 1481 controls and revealed no genome-wide significant loci (λ_GC_ = 0.918). The meta-analysis resulted in 8465647 SNPs.

**Supplementary Figure 8. East Asian ancestry GWAS**
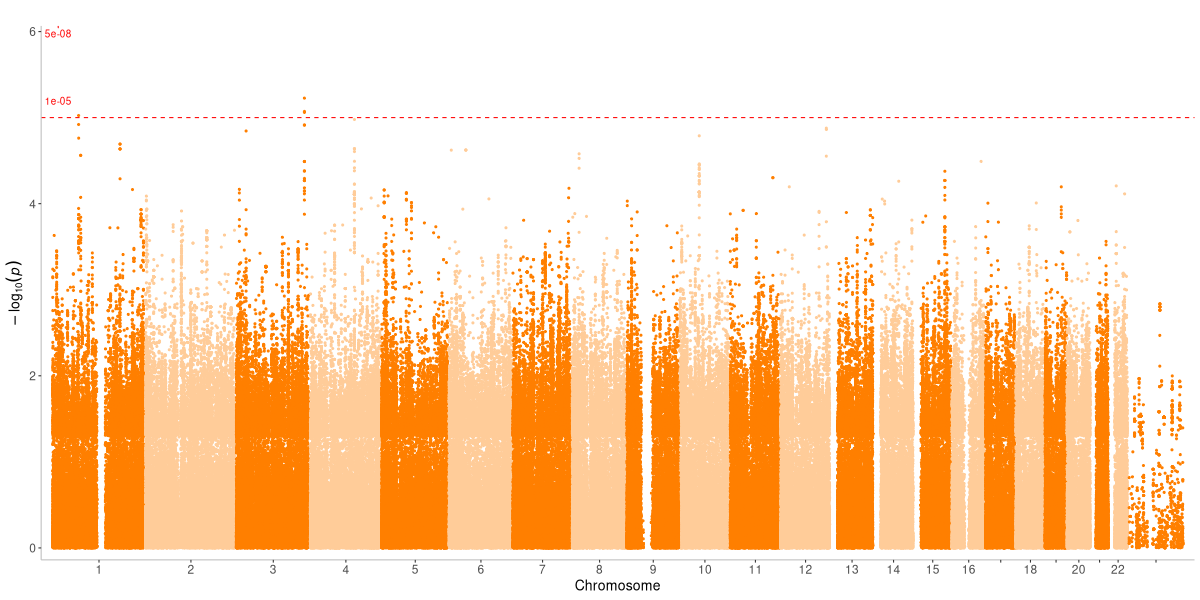

Independent East Asian ancestry analyses included 340 cases and 586 controls and revealed no genome-wide significant loci (λ_GC_ = 0.948). The meta-analysis resulted in 3137719 SNPs.

**Supplementary Figure 9. European GWAS meta-analysis MAGMA gene-based analysis Manhattan plot. 23 significant genes are labelled.**

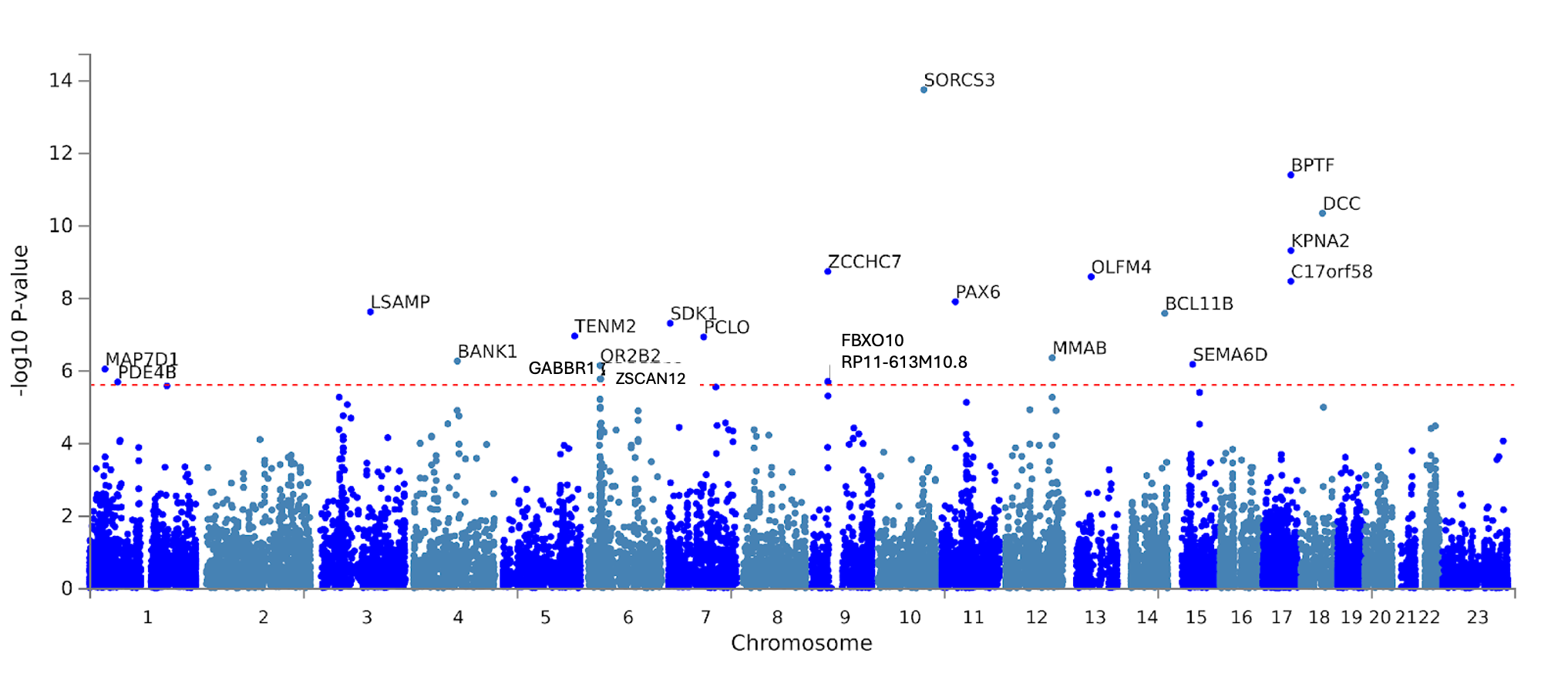

**Supplementary Figure 10. GTEx v8 30 general tissue type enrichment for the European meta-analysis**

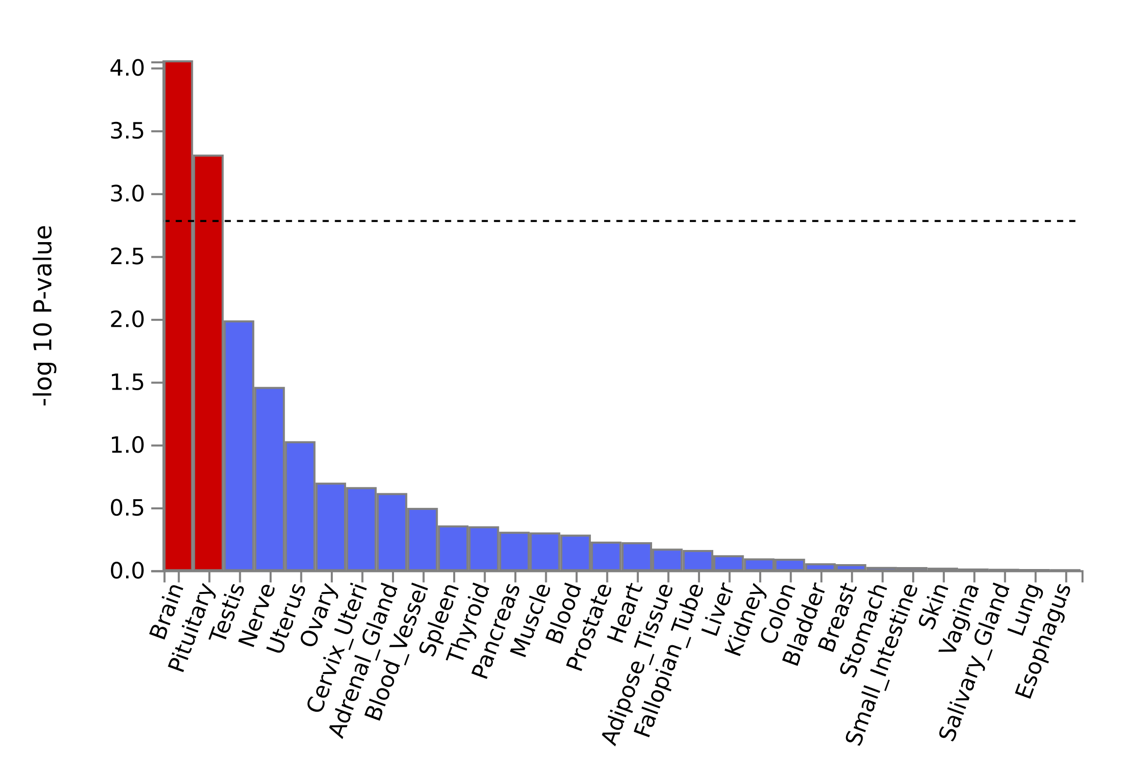

**Supplementary Figure 11. GTEx v8 53 specific tissue type enrichment for the European meta-analysis**

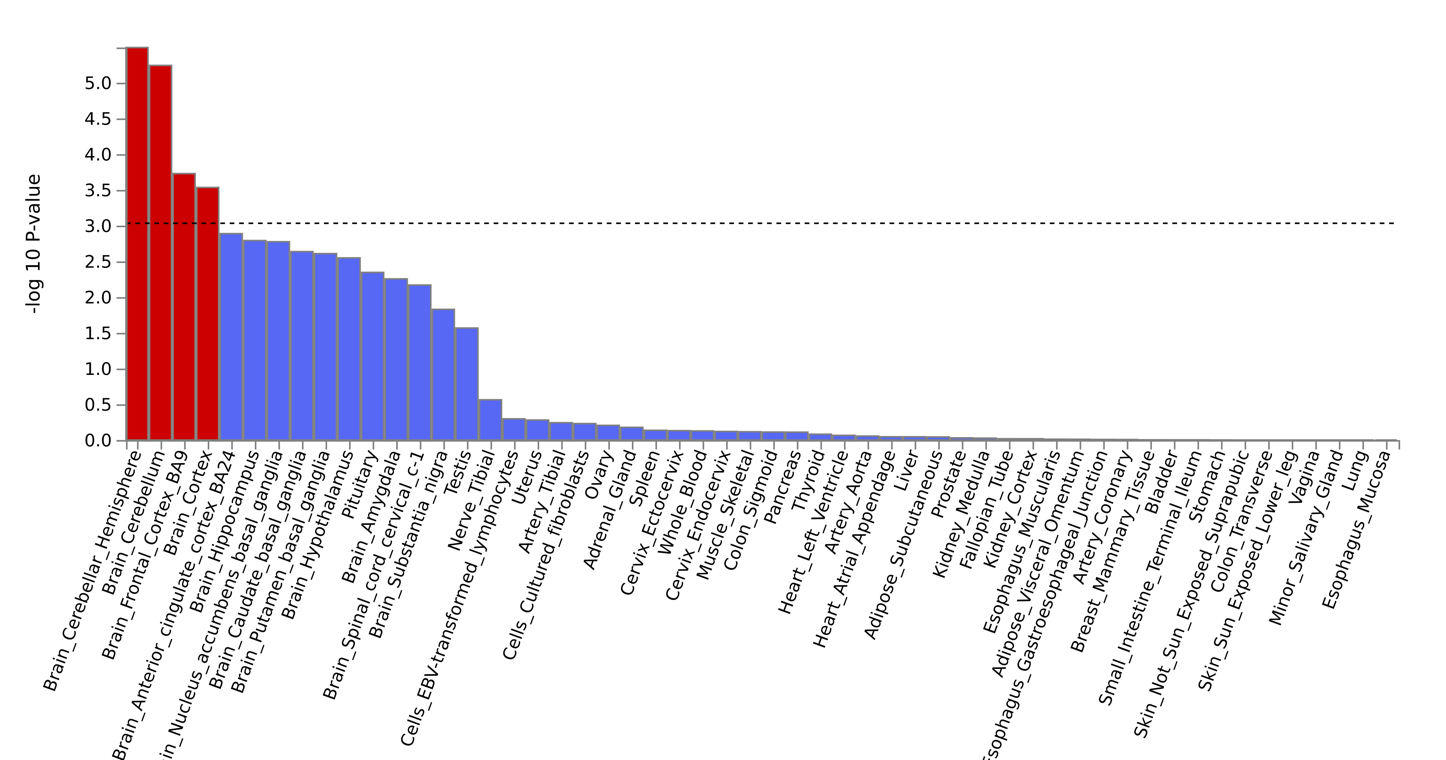

**Supplementary Figure 12. Genetic correlations between traits and adolescent-onset depression by ascertainment phenotype**

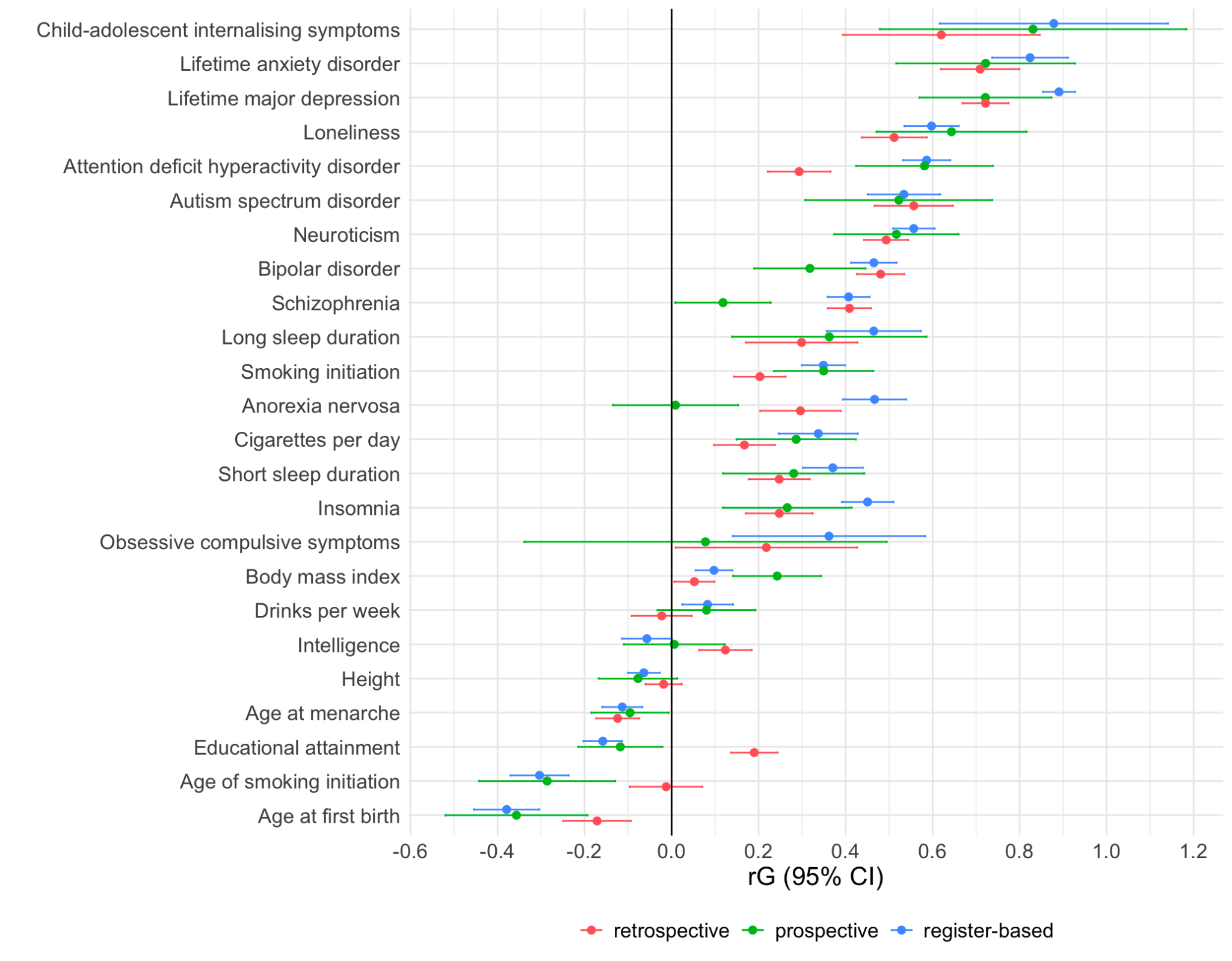

**Supplementary Figure 13. Comparison of genetic correlations between traits and adolescent-onset depression versus lifetime major depression**

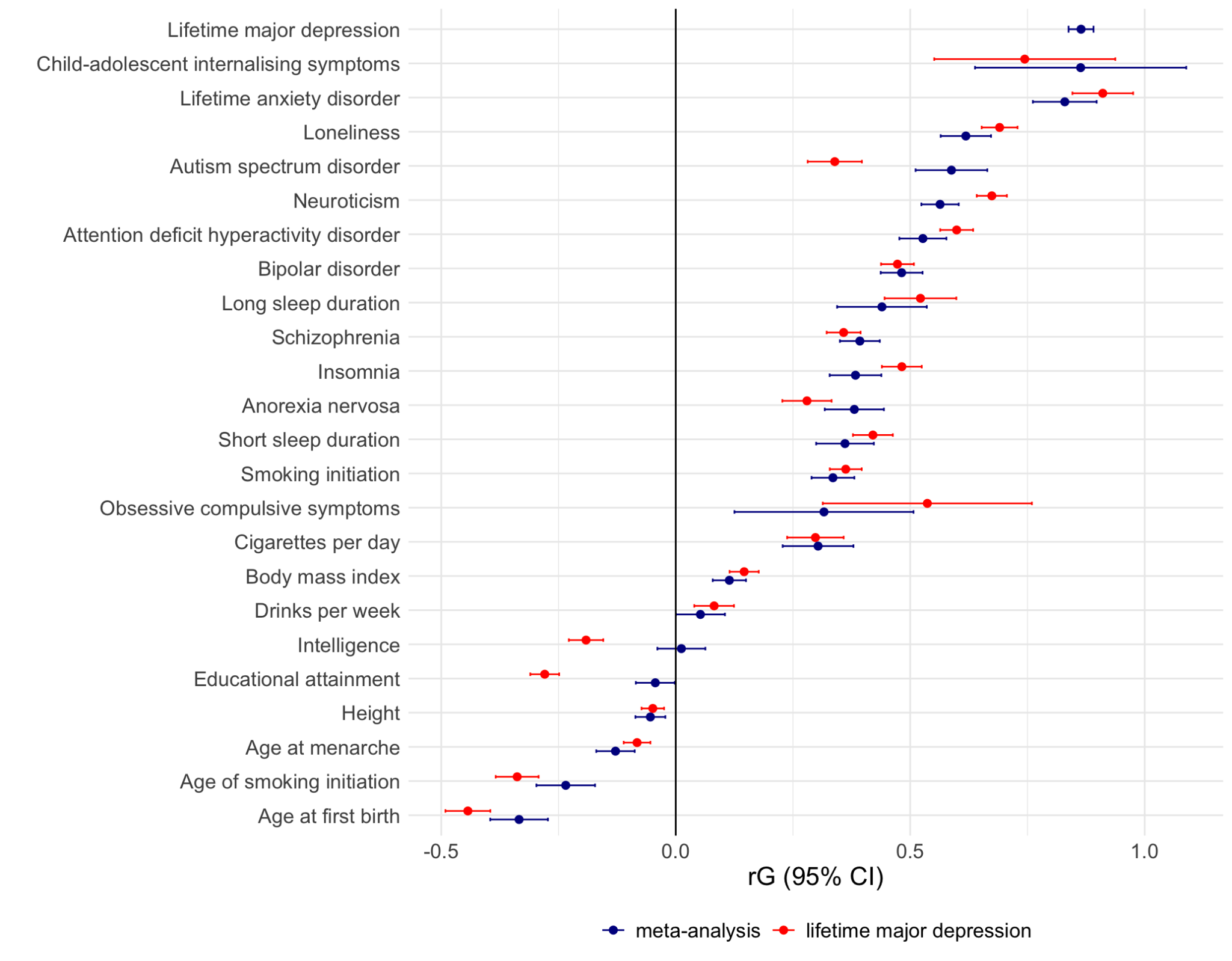

**Supplementary Figure 13. Association of PGS derived from European-only summary statistics with depression case-status in the ABCD and Add Health cohorts**

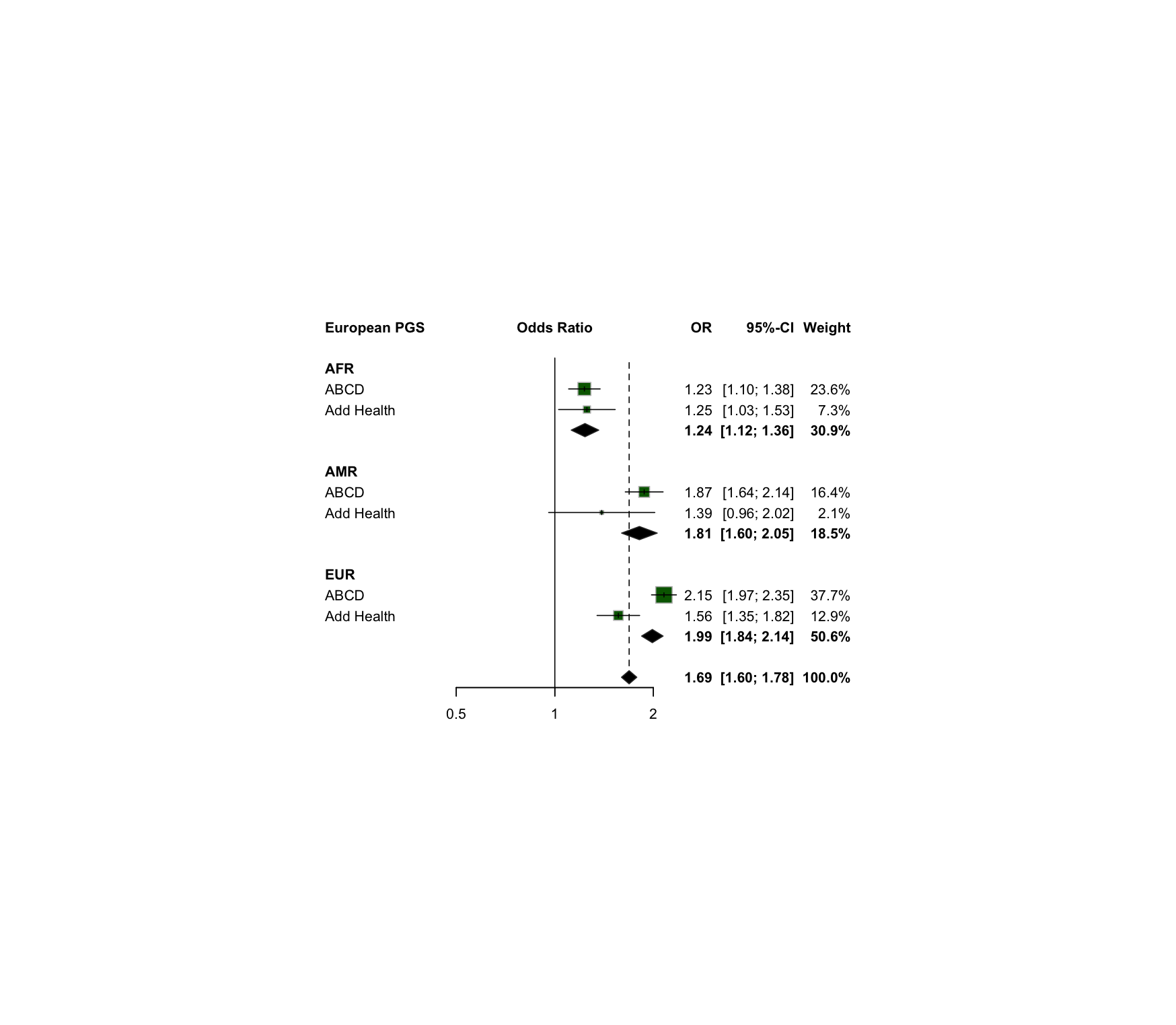

**Supplementary Figure 14. Association between European adolescent-onset depression PGS and global structural brain features in the ABCD study**

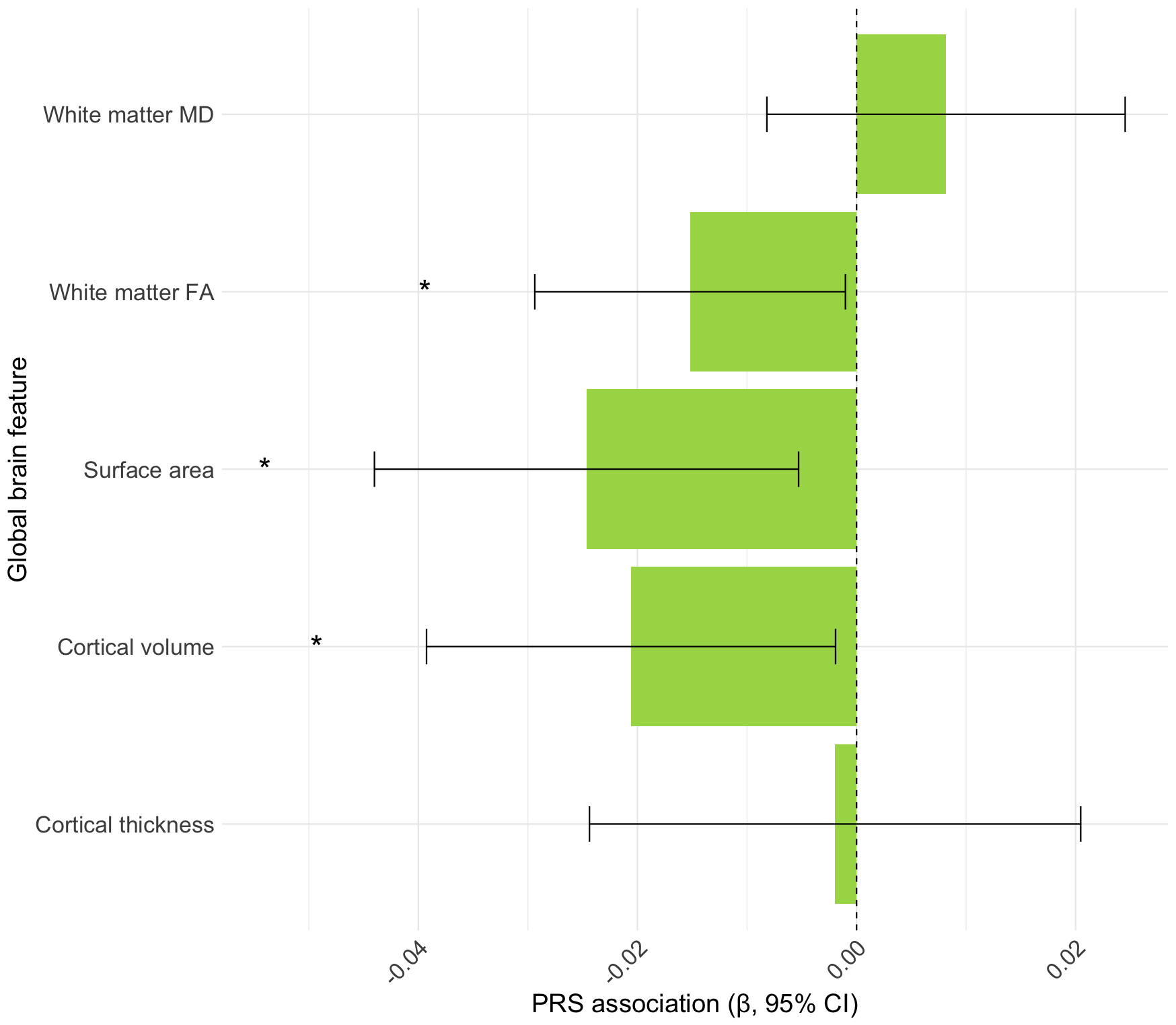

**Supplementary Figure 15. Association between European adolescent-onset depression PGS and regional structural brain features in the ABCD study**
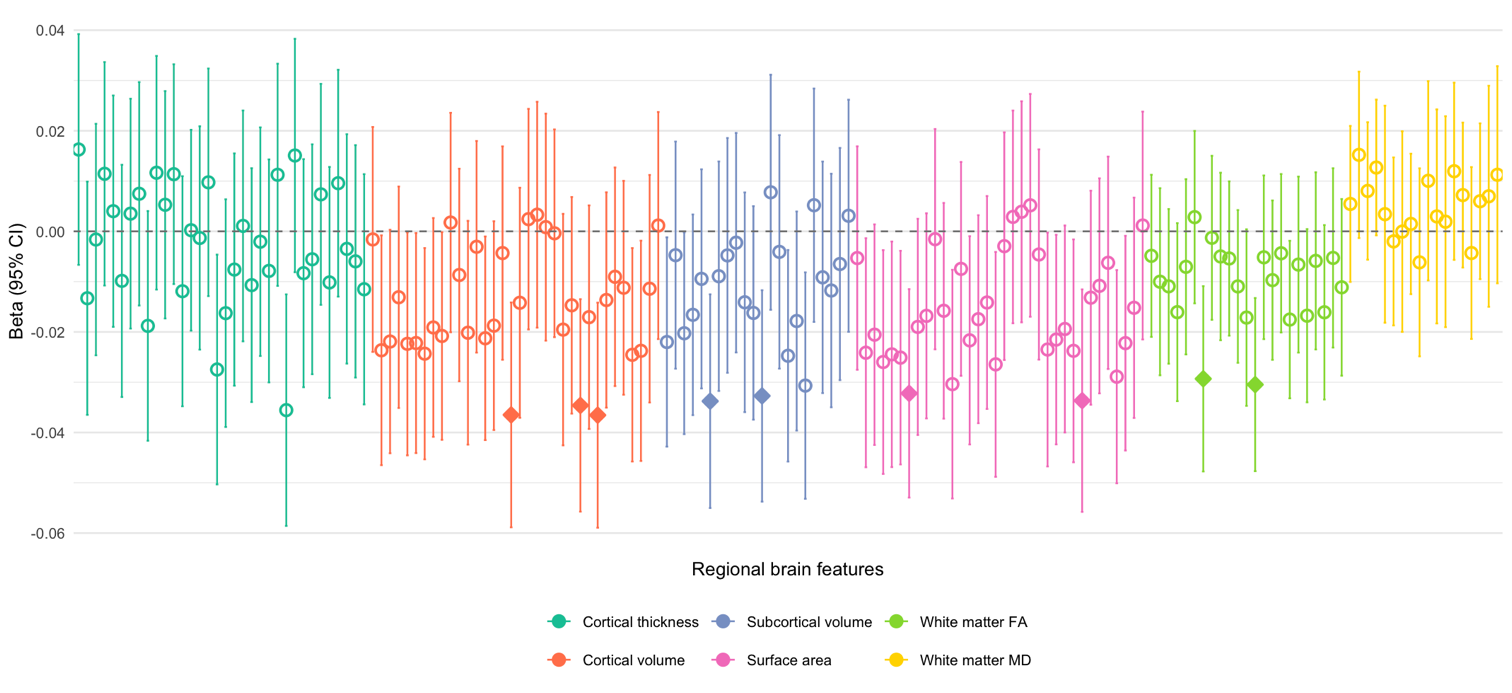

The y-axis represents the FDR corrected (q < 0.05) standardised regression coefficients with 95% confidence intervals. Features are split by imaging modalities plotted along the x-axis. Solid diamonds represent features significantly associated with PGS after FDR correction. Here significant features are reduced cortical volume of the paracentral lobule, precuneus, rostral anterior cingulate; reduced subcortical volume of the hippocampus and ventral diencephalon; reduced surface area of the fusiform gyrus and rostral anterior cingulate; and reduced white matter FA of the superior corticostriatal tract and right corticospinal tract.

**Supplementary Figure 16. Fixed effects cross-ancestry GWAS**

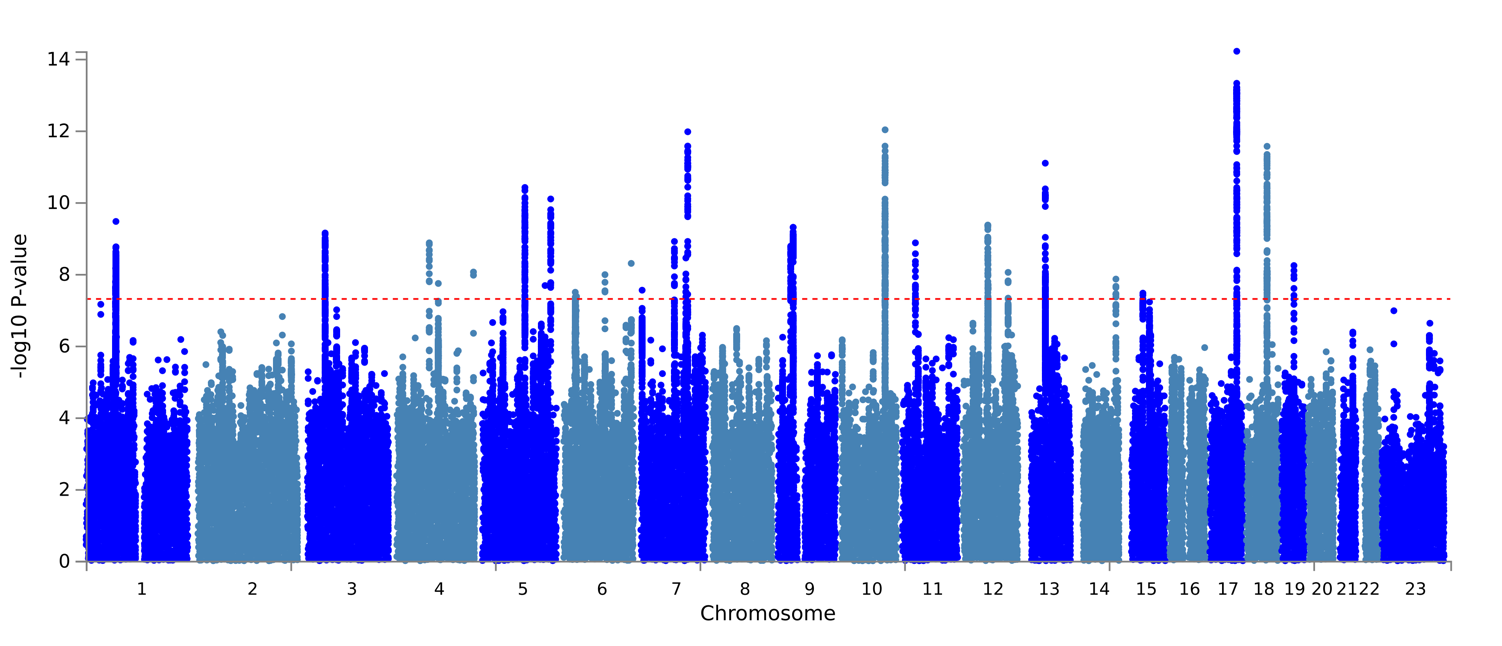

Results for the fixed effects inverse-weighted standard error meta-analysis in METAL of all ancestries used to generate cross-ancestry PGS. The meta-analysis revealed 28 genome-wide significant loci. This method does not account for ancestral heterogeneity amongst cohorts.

**Supplementary Figure 17. Derivation of depression trajectories in ALSPAC using latent class growth analysis**

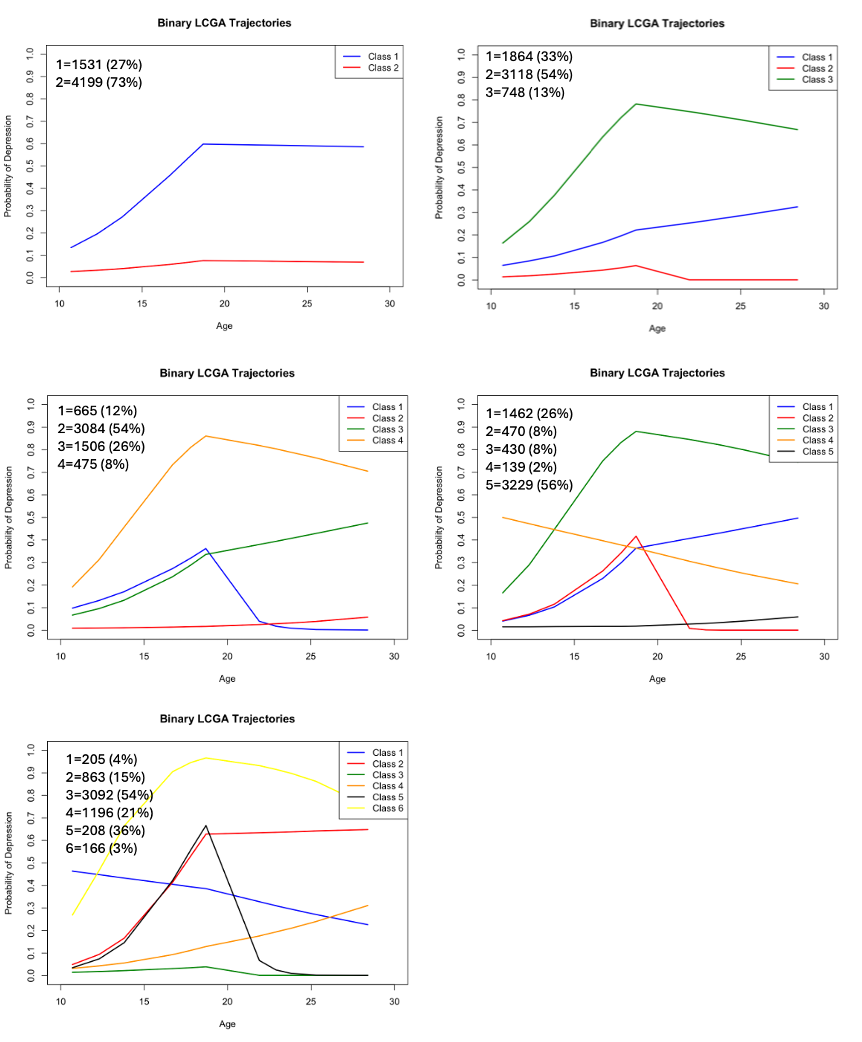

**Supplementary References**

Anckarsäter, H., Lundström, S., Kollberg, L., Kerekes, N., Palm, C., Carlström, E., Långström, N., Magnusson, P. K. E., Halldner, L., Bölte, S., Gillberg, C., Gumpert, C., Råstam, M., & Lichtenstein, P. (2011). The Child and Adolescent Twin Study in Sweden (CATSS). *Twin Research and Human Genetics*, *14*(6), 495–508. https://doi.org/10.1375/twin.14.6.495

Bosch, R., Pagerols, M., Rivas, C., Sixto, L., Bricollé, L., Español-Martín, G., Prat, R., Ramos-Quiroga, J. A., & Casas, M. (2022). Neurodevelopmental disorders among Spanish school-age children: Prevalence and sociodemographic correlates. *Psychological Medicine*, *52*(14), 3062–3072. https://doi.org/10.1017/S0033291720005115

Boyd, A., Golding, J., Macleod, J., Lawlor, D. A., Fraser, A., Henderson, J., Molloy, L., Ness, A., Ring, S., & Davey Smith, G. (2013). Cohort Profile: The ’children of the 90s’--the index offspring of the Avon Longitudinal Study of Parents and Children. *International Journal of Epidemiology*, *42*(1), 111–127. https://doi.org/10.1093/ije/dys064

Bycroft, C., Freeman, C., Petkova, D., Band, G., Elliott, L. T., Sharp, K., Motyer, A., Vukcevic, D., Delaneau, O., O’Connell, J., Cortes, A., Welsh, S., Young, A., Effingham, M., McVean, G., Leslie, S., Allen, N., Donnelly, P., & Marchini, J. (2018). The UK Biobank resource with deep phenotyping and genomic data. *Nature*, *562*(7726), 203–209. https://doi.org/10.1038/s41586-018-0579-z

Byrne, E. M., Kirk, K. M., Medland, S. E., McGrath, J. J., Colodro-Conde, L., Parker, R., Cross, S., Sullivan, L., Statham, D. J., Levinson, D. F., Licinio, J., Wray, N. R., Hickie, I. B., & Martin, N. G. (2020). Cohort profile: The Australian genetics of depression study. *BMJ Open*, *10*(5). https://doi.org/10.1136/bmjopen-2019-032580

Costello, E. J., Eaves, L., Sullivan, P., Kennedy, M., Conway, K., Adkins, D. E., Angold, A., Clark, S. L., Erkanli, A., McClay, J. L., Copeland, W., Maes, H. H., Liu, Y., Patkar, A. A., Silberg, J., & van den Oord, E. (2013). Genes, environments, and developmental research: Methods for a multi-site study of early substance abuse. *Twin Research and Human Genetics: The Official Journal of the International Society for Twin Studies*, *16*(2), 505–515. https://doi.org/10.1017/thg.2013.6

Couvy-Duchesne, B., O’Callaghan, V., Parker, R., Mills, N., Kirk, K. M., Scott, J., Vinkhuyzen, A., Hermens, D. F., Lind, P. A., Davenport, T. A., Burns, J. M., Connell, M., Zietsch, B. P., Scott, J., Wright, M. J., Medland, S. E., McGrath, J., Martin, N. G., Hickie, I. B., & Gillespie, N. A. (2018). Nineteen and Up study (19Up): Understanding pathways to mental health disorders in young Australian twins. *BMJ Open*, *8*(3), e018959. https://doi.org/10.1136/bmjopen-2017-018959

Davies, M. R., Kalsi, G., Armour, C., Jones, I. R., McIntosh, A. M., Smith, D. J., Walters, J. T. R., Bradley, J. R., Kingston, N., Ashford, S., Beange, I., Brailean, A., Cleare, A. J., Coleman, J. R. I., Curtis, C. J., Curzons, S. C. B., Davis, K. A. S., Dowey, L. R. C., Gault, V. A., … Breen, G. (2019). The Genetic Links to Anxiety and Depression (GLAD) Study: Online recruitment into the largest recontactable study of depression and anxiety. *Behaviour Research and Therapy*, *123*, 103503. https://doi.org/10.1016/j.brat.2019.103503

Davis, K. A. S., Cullen, B., Adams, M., Brailean, A., Breen, G., Coleman, J. R. I., Dregan, A., Gaspar, H. A., Hübel, C., Lee, W., McIntosh, A. M., Nolan, J., Pearsall, R., & Hotopf, M. (2019). Indicators of mental disorders in UK Biobank—A comparison of approaches. *International Journal of Methods in Psychiatric Research*, *28*(3), e1796. https://doi.org/10.1002/mpr.1796

Fernandez-Pujals, A. M., Adams, M. J., Thomson, P., McKechanie, A. G., Blackwood, D. H. R., Smith, B. H., Dominiczak, A. F., Morris, A. D., Matthews, K., Campbell, A., Linksted, P., Haley, C. S., Deary, I. J., Porteous, D. J., MacIntyre, D. J., & McIntosh, A. M. (2015). Epidemiology and Heritability of Major Depressive Disorder, Stratified by Age of Onset, Sex, and Illness Course in Generation Scotland: Scottish Family Health Study (GS:SFHS). *PloS One*, *10*(11), e0142197. https://doi.org/10.1371/journal.pone.0142197

Fraser, A., Macdonald-Wallis, C., Tilling, K., Boyd, A., Golding, J., Davey Smith, G., Henderson, J., Macleod, J., Molloy, L., Ness, A., Ring, S., Nelson, S. M., & Lawlor, D. A. (2013). Cohort Profile: The Avon Longitudinal Study of Parents and Children: ALSPAC mothers cohort. *International Journal of Epidemiology*, *42*(1), 97–110. https://doi.org/10.1093/ije/dys066

Ghatan, S., Vries, J. de, Pingault, J.-B., Jaddoe, V. W., Cecil, C., Felix, J. F., Rivadeneira, F., & Medina-Gomez, C. (2024). *Genetic Nurture: Estimating the direct genetic effects of pediatric anthropometric traits* (p. 2024.12.10.24318796). medRxiv. https://doi.org/10.1101/2024.12.10.24318796

Gillespie, N. A., Henders, A. K., Davenport, T. A., Hermens, D. F., Wright, M. J., Martin, N. G., & Hickie, I. B. (2013). The Brisbane Longitudinal Twin Study: Pathways to Cannabis Use, Abuse, and Dependence Project—Current Status, Preliminary Results, and Future Directions. *Twin Research and Human Genetics*, *16*(1), 21–33. https://doi.org/10.1017/thg.2012.111

Harris, K. M., Halpern, C. T., Whitsel, E. A., Hussey, J. M., Killeya-Jones, L. A., Tabor, J., & Dean, S. C. (2019). Cohort Profile: The National Longitudinal Study of Adolescent to Adult Health (Add Health). *International Journal of Epidemiology*, *48*(5), 1415–1415k. https://doi.org/10.1093/ije/dyz115

Harris, P. A., Taylor, R., Thielke, R., Payne, J., Gonzalez, N., & Conde, J. G. (2009). Research electronic data capture (REDCap)—A metadata-driven methodology and workflow process for providing translational research informatics support. *Journal of Biomedical Informatics*, *42*(2), 377–381. https://doi.org/10.1016/j.jbi.2008.08.010

Hartman, C. A., Richards, J. S., Vrijen, C., Oldehinkel, A. J., Oerlemans, A. M., & Kretschmer, T. (2022). Cohort Profile Update: The TRacking Adolescents’ Individual Lives Survey-The Next Generation (TRAILS NEXT). *International Journal of Epidemiology*, *51*(5), e267–e275. https://doi.org/10.1093/ije/dyac066

Hickie, I. B., Davenport, T. A., Hadzi-Pavlovic, D., Koschera, A., Naismith, S. L., Scott, E. M., & Wilhelm, K. A. (2001). Development of a simple screening tool for common mental disorders in general practice. *The Medical Journal of Australia*, *175*(S1), S10-7. https://doi.org/10.5694/j.1326-5377.2001.tb143784.x

Joshi, H. E., & Fitzsimons, E. (2016). The UK Millennium Cohort: The making of a multipurpose resource for social science and policy. *Longitudinal and Life Course Studies*, *7*(4), Article 4.

Lockhart, C., Bright, J., Ahmadzadeh, Y., Breen, G., Bristow, S., Boyd, A., Downs, J., Hotopf, M., Palaiologou, E., Rimfeld, K., Maxwell, J., Malanchini, M., McAdams, T. A., McMillan, A., Plomin, R., & Eley, T. C. (2023). Twins Early Development Study (TEDS): A genetically sensitive investigation of mental health outcomes in the mid‐twenties. *JCPP Advances*, *3*(2), e12154. https://doi.org/10.1002/jcv2.12154

Magnus, P., Birke, C., Vejrup, K., Haugan, A., Alsaker, E., Daltveit, A. K., Handal, M., Haugen, M., Høiseth, G., Knudsen, G. P., Paltiel, L., Schreuder, P., Tambs, K., Vold, L., & Stoltenberg, C. (2016). Cohort Profile Update: The Norwegian Mother and Child Cohort Study (MoBa). *International Journal of Epidemiology*, *45*(2), 382–388. https://doi.org/10.1093/ije/dyw029

Major Depressive Disorder Working Group of the Psychiatric GWAS Consortium, Ripke, S., Wray, N. R., Lewis, C. M., Hamilton, S. P., Weissman, M. M., Breen, G., Byrne, E. M., Blackwood, D. H. R., Boomsma, D. I., Cichon, S., Heath, A. C., Holsboer, F., Lucae, S., Madden, P. A. F., Martin, N. G., McGuffin, P., Muglia, P., Noethen, M. M., … Sullivan, P. F. (2013). A mega-analysis of genome-wide association studies for major depressive disorder. *Molecular Psychiatry*, *18*(4), 497–511. https://doi.org/10.1038/mp.2012.21

Medina-Gomez, C., Felix, J. F., Estrada, K., Peters, M. J., Herrera, L., Kruithof, C. J., Duijts, L., Hofman, A., van Duijn, C. M., Uitterlinden, A. G., Jaddoe, V. W. V., & Rivadeneira, F. (2015). Challenges in conducting genome-wide association studies in highly admixed multi-ethnic populations: The Generation R Study. *European Journal of Epidemiology*, *30*(4), 317–330. https://doi.org/10.1007/s10654-015-9998-4

Mitchell, B. L., Campos, A. I., Rentería, M. E., Parker, R., Sullivan, L., McAloney, K., Couvy-Duchesne, B., Medland, S. E., Gillespie, N. A., Scott, J., Zietsch, B. P., Lind, P. A., Martin, N. G., & Hickie, I. B. (2019). Twenty-Five and Up (25Up) Study: A New Wave of the Brisbane Longitudinal Twin Study. *Twin Research and Human Genetics: The Official Journal of the International Society for Twin Studies*, *22*(3), 154–163. https://doi.org/10.1017/thg.2019.27

Najman, J. M., Alati, R., Bor, W., Clavarino, A., Mamun, A., McGrath, J. J., McIntyre, D., O’Callaghan, M., Scott, J., Shuttlewood, G., Williams, G. M., & Wray, N. (2015). Cohort Profile Update: The Mater-University of Queensland Study of Pregnancy (MUSP). *International Journal of Epidemiology*, *44*(1), 78–78f. https://doi.org/10.1093/ije/dyu234

Newnham, J. P., Evans, S. F., Michael, C. A., Stanley, F. J., & Landau, L. I. (1993). Effects of frequent ultrasound during pregnancy: A randomised controlled trial. *Lancet (London, England)*, *342*(8876), 887–891. https://doi.org/10.1016/0140-6736(93)91944-h

Northstone, K., Lewcock, M., Groom, A., Boyd, A., Macleod, J., Timpson, N., & Wells, N. (2019). The Avon Longitudinal Study of Parents and Children (ALSPAC): An update on the enrolled sample of index children in 2019. *Wellcome Open Research*, *4*, 51. https://doi.org/10.12688/wellcomeopenres.15132.1

Oldehinkel, A. J., Rosmalen, J. G., Buitelaar, J. K., Hoek, H. W., Ormel, J., Raven, D., Reijneveld, S. A., Veenstra, R., Verhulst, F. C., Vollebergh, W. A., & Hartman, C. A. (2015). Cohort Profile Update: The TRacking Adolescents’ Individual Lives Survey (TRAILS). *International Journal of Epidemiology*, *44*(1), 76–76n. https://doi.org/10.1093/ije/dyu225

Olsen, C. M., Green, A. C., Neale, R. E., Webb, P. M., Cicero, R. A., Jackman, L. M., O’Brien, S. M., Perry, S. L., Ranieri, B. A., Whiteman, D. C., & QSkin Study. (2012). Cohort profile: The QSkin Sun and Health Study. *International Journal of Epidemiology*, *41*(4), 929–929i. https://doi.org/10.1093/ije/dys107

Scholtens, S., Smidt, N., Swertz, M. A., Bakker, S. J., Dotinga, A., Vonk, J. M., van Dijk, F., van Zon, S. K., Wijmenga, C., Wolffenbuttel, B. H., & Stolk, R. P. (2015). Cohort Profile: LifeLines, a three-generation cohort study and biobank. *International Journal of Epidemiology*, *44*(4), 1172–1180. https://doi.org/10.1093/ije/dyu229

Sijtsma, A., Rienks, J., van der Harst, P., Navis, G., Rosmalen, J. G. M., & Dotinga, A. (2022). Cohort Profile Update: Lifelines, a three-generation cohort study and biobank. *International Journal of Epidemiology*, *51*(5), e295–e302. https://doi.org/10.1093/ije/dyab257

Smith, B. H., Campbell, A., Linksted, P., Fitzpatrick, B., Jackson, C., Kerr, S. M., Deary, I. J., MacIntyre, D. J., Campbell, H., McGilchrist, M., Hocking, L. J., Wisely, L., Ford, I., Lindsay, R. S., Morton, R., Palmer, C. N. A., Dominiczak, A. F., Porteous, D. J., & Morris, A. D. (2013). Cohort Profile: Generation Scotland: Scottish Family Health Study (GS:SFHS). The study, its participants and their potential for genetic research on health and illness. *International Journal of Epidemiology*, *42*(3), 689–700. https://doi.org/10.1093/ije/dys084

Soh, S.-E., Chong, Y.-S., Kwek, K., Saw, S.-M., Meaney, M. J., Gluckman, P. D., Holbrook, J. D., Godfrey, K. M., & GUSTO Study Group. (2014). Insights from the Growing Up in Singapore Towards Healthy Outcomes (GUSTO) cohort study. *Annals of Nutrition & Metabolism*, *64*(3–4), 218–225. https://doi.org/10.1159/000365023

Sudlow, C., Gallacher, J., Allen, N., Beral, V., Burton, P., Danesh, J., Downey, P., Elliott, P., Green, J., Landray, M., Liu, B., Matthews, P., Ong, G., Pell, J., Silman, A., Young, A., Sprosen, T., Peakman, T., & Collins, R. (2015). UK biobank: An open access resource for identifying the causes of a wide range of complex diseases of middle and old age. *PLoS Medicine*, *12*(3), e1001779. https://doi.org/10.1371/journal.pmed.1001779

*The World Health Organization Composite International Diagnostic Interview short‐form (CIDI‐SF)—Kessler—1998—International Journal of Methods in Psychiatric Research—Wiley Online Library*. (n.d.). Retrieved 11 February 2025, from https://onlinelibrary.wiley.com/doi/abs/10.1002/mpr.47

van Eijsden, M., Vrijkotte, T. G., Gemke, R. J., & van der Wal, M. F. (2011). Cohort Profile: The Amsterdam Born Children and their Development (ABCD) Study. *International Journal of Epidemiology*, *40*(5), 1176–1186. https://doi.org/10.1093/ije/dyq128

Volkow, N. D., Koob, G. F., Croyle, R. T., Bianchi, D. W., Gordon, J. A., Koroshetz, W. J., Pérez-Stable, E. J., Riley, W. T., Bloch, M. H., Conway, K., Deeds, B. G., Dowling, G. J., Grant, S., Howlett, K. D., Matochik, J. A., Morgan, G. D., Murray, M. M., Noronha, A., Spong, C. Y., … Weiss, S. R. B. (2018). The conception of the ABCD study: From substance use to a broad NIH collaboration. *Developmental Cognitive Neuroscience*, *32*, 4–7. https://doi.org/10.1016/j.dcn.2017.10.002

Wray, N. R., Ripke, S., Mattheisen, M., Trzaskowski, M., Byrne, E. M., Abdellaoui, A., Adams, M. J., Agerbo, E., Air, T. M., Andlauer, T. M. F., Bacanu, S.-A., Bækvad-Hansen, M., Beekman, A. F. T., Bigdeli, T. B., Binder, E. B., Blackwood, D. R. H., Bryois, J., Buttenschøn, H. N., Bybjerg-Grauholm, J., … Sullivan, P. F. (2018). Genome-wide association analyses identify 44 risk variants and refine the genetic architecture of major depression. *Nature Genetics*, *50*(5), Article 5. https://doi.org/10.1038/s41588-018-0090-3

Zafarmand, M. H., Spanjer, M., Nicolaou, M., Wijnhoven, H. A. H., Schaik, B. D. C. van, Uitterlinden, A. G., Snieder, H., & Vrijkotte, T. G. M. (2020). Influence of Dietary Approaches to Stop Hypertension-Type Diet, Known Genetic Variants and Their Interplay on Blood Pressure in Early Childhood. *Hypertension*. https://doi.org/10.1161/HYPERTENSIONAHA.118.12292

Zagai, U., Lichtenstein, P., Pedersen, N. L., & Magnusson, P. K. E. (2019). The Swedish Twin Registry: Content and Management as a Research Infrastructure. *Twin Research and Human Genetics: The Official Journal of the International Society for Twin Studies*, *22*(6), 672–680. https://doi.org/10.1017/thg.2019.99
